# Estimated number of lives directly saved by COVID-19 vaccination programs in the WHO European Region, December 2020 to March 2023

**DOI:** 10.1101/2024.01.12.24301206

**Authors:** The WHO European Respiratory Surveillance Network

**Affiliations:** World Health Organization Regional Office for Europe, Copenhagen, Denmark; Österreichische Agentur für Gesundheit und Ernährungssicherheit, Vienna, Austria; Medical University, Vienna, Austria; Sciensano, Brussels, Belgium; Medical and Public Health Services, Ministry of Health, Nicosia, Cyprus; National Institute of Public Health, Prague, Czechia; Third Faculty of Medicine, Charles University, Prague, Czechia; Institute of Health Information and Statistics of the Czech Republic, Nové Město, Czechia; Statens Serum Institute, Copenhagen, Denmark; Finnish Institute for Health and Welfare, Helsinki, Finland; Hellenic National Public Health Organization, Athens, Greece; National Center for Public Health and Pharmacy, Budapest, Hungary; Centre for Health Security and Communicable Disease Control, Reykjavik, Iceland; Health Service Executive-Health Protection Surveillance Centre, Dublin, Ireland; Instituto Superiore di Sanità, Rome, Italy; National Public Health Center under the Ministry of Health, Vilnius, Lithuania; Health Directorate, Luxembourg, Luxembourg; Infectious Disease Prevention and Control Unit, Health Promotion and Disease Prevention Directorate, Pieta’, Malta; Institute of Public Health of Republic of North Macedonia, Skopje, North Macedonia; Norwegian Institute of Public Health, Oslo, Norway; Directorate of Disease Prevention and Health Promotion, Directorate-General of Health, Lisbon, Portugal; National Institute of Health Doutor Ricardo Jorge, Lisbon, Portugal; Directorate of Information and Analysis, Directorate-General of Health, Lisbon, Portugal; National Agency for Public Health, Chisinau, Republic of Moldova; "Cantacuzino" National Military Medical Institut for Research and Development, Bucharest, Romania; National Institute of Public Health, București, Romania; Regional Public Health Authority, Banská Bystrica, Slovakia; National Institute of Public Health, Ljubljana, Slovenia; Ministry of Health, Madrid, Spain; Instituto de Salud Carlos III, Madrid, Spain; UK Health Security Agency, London, United Kingdom; Public Health Scotland, Glasgow, United Kingdom

## Abstract

**Background:** By March 2023, 54 countries, areas and territories (thereafter “CAT”) reported over 2.2 million coronavirus disease 2019 (COVID-19) deaths to the World Health Organization (WHO) Regional Office for Europe (1). Here, we estimate how many lives were directly saved by vaccinating adults in the Region, from December 2020 through March 2023.

**Methods:** We estimated the number of lives directly saved by age-group, vaccine dose and circulating Variant of Concern (VOC) period, both regionally and nationally, using weekly data on COVID-19 mortality and COVID-19 vaccine uptake reported by 34 CAT, and vaccine effectiveness (VE) data from the literature. We calculated the percentage reduction in the number of expected and reported deaths.

**Findings:** We found that vaccines reduced deaths by 57% overall (CAT range: 15% to 75%), representing ∼1.4 million lives saved in those aged ≥25 years (range: 0.7 million to 2.6 million): 96% of lives saved were aged ≥60 years and 52% were aged ≥80 years; first boosters saved 51%, and 67% were saved during the Omicron period.

**Interpretation:** Over nearly 2.5 years, most lives saved by COVID-19 vaccinationwere in older adults by first booster dose and during the Omicron period, reinforcing the importance of up-to-date vaccination among these most at-risk individuals. Further modelling work should evaluate indirect effects of vaccination and public health and social measures.

**Funding:** This work was supported by a US Centers for Disease Control cooperative agreement (Grant number 6 NU511P000936-02-020), who had no role in data analysis or interpretation.

**Disclaimer:** The authors affiliated with the World Health Organization (WHO) are alone responsible for the views expressed in this publication and they do not necessarily represent the decisions or policies of the WHO.

**Research in context:** *Evidence before this study:* Since first identified in late 2019, COVID-19 has caused disproportionately high mortality rates in older adults. With the rapid development and licensing of novel COVID-19 vaccines, immunization campaigns across the WHO European Region started in late 2020 and early 2021, initially targeting the most vulnerable and exposed populations, including older adults, people with comorbidities and healthcare professionals. Several studies have estimated the number of lives saved by COVID-19 vaccination, both at national and multi-country level in the earlier stages of the pandemic. However, only one multi-country study has assessed the number of lives saved beyond the first year of the pandemic, particularly when the Omicron variant of concern (VOC) circulated, a period when vaccination coverage was high in many countries, areas and territories (CAT), but COVID-19 transmission was at its highest.

*Added value of this study:* Here we quantified the impact of COVID-19 vaccination in adults by age-group, vaccine dose and period of circulation of VOC, across diverse settings, using real world data reported by 34 CAT in the WHO European Region for the period December 2020 to April 2023. We estimated that COVID-19 vaccination programs were associated with a 57% reduction (CAT range: 15% to 75%) in the number of deaths among the ≥25 years old, representing over 1.5 million lives saved (range: 0.7 million to 2.6 million) in 34 European CAT during the first 2.5 years following vaccine introduction. The first booster savedthe most lives (721,122 / 1,408,967, (57%) of all lives saved). The ≥60 years old age group accounted for 96% of the total lives saved (1,349,617 / 1,408,967) whereas the ≥80 years old age group represented 52% of the total lives saved (728,858 / 1,408,967 lives saved) and 67% of all lives were saved during the Omicron period (942,571 / 1,408,967).

*Implications of all the available evidence:* Our results reinforce the importance of up-to-date COVID-19 vaccination, particularly among older age-groups. Communication campaigns supporting COVID-19 vaccination should stress the value of COVID-19 vaccination in saving lives to ensure vulnerable groups are up-to-date with vaccination ahead of periods of potential increased transmission.

## Introduction

From the beginning of the pandemic through March 2023, 2.2 million coronavirus disease 2019 (COVID-19) deaths were reported to the World Health Organization (WHO) Regional Office for Europe from the 54 countries, areas and territories (CAT) in the WHO European Region (1). The true number of deaths directly or indirectly linked to COVID-19 is estimated to be even greater (2).

Throughout the COVID-19 pandemic, disproportionately higher mortality rates have been observed in older age-groups. Indeed a global review of publicly available data from 2020-2022 found that persons aged ≥60 years old accountedfor over 80% of all COVID-19 fatalities (3), a pattern that has been consistently observed in other studies (4–6).

Since they were first introduced in late 2020, COVID-19 vaccines have been shown to be safe and highly effective in protecting against severe COVID-19 infection (7,8). In the European Region, since the first COVID-19 vaccines were administered (7,8), WHO has recommended that older age-groups be prioritized for COVID-19 vaccination (10). As of March 2023, 69% of people over ≥60 years in 49 CAT across the Region were reported to have received at least three doses of vaccine (7).

Previous studies have estimated the number of lives saved by COVID-19 vaccine in individual countries at various stages following the introduction of COVID-19 vaccination programs (4,5,11–15). We previously estimated that vaccination directly saved 469,186 lives among people ≥60 years old in 33 countries in the first year of the vaccination program in Europe (12). Another study estimated the total number of lives saved by vaccine during 2021 in 185 countries and territories (8). Only one study (3), has estimated the number of lives saved beyond 2021 (analysis up to late 2022), despite the continued circulation of Severe Acute Respiratory Corona Virus 2 (SARS-CoV-2), and in particular the Variant of Concern (VOC), Omicron, in the two years since. We aimed to expand on our previous work by estimating the number of lives saved by COVID-19 vaccination in adults ≥25 years of age in the European Region from the beginning of vaccine introduction through March 2023, a period of 2.5 years. We stratified our results by age-group, predominant circulating VOC and vaccination dose and considered waning protection and prior infection in our analysis. Lastly, we aimed to understand the varying impact of infections by age group.

## Methods

### Data sources

We estimated lives saves during the study period using CAT-level COVID-19 surveillance data and vaccination coverage data from week 50/2020 (December 2020) to week 12/2023 (March 2023) (hereafter referred to as “the study period”).

As part of COVID-19 routine surveillance reporting, which is jointly coordinated by the WHO Regional Office for Europe and the European Center for Disease Prevention and Control (ECDC), every week countries in the European Region provided data on COVID-19 mortality and infection, COVID-19 vaccination uptake and SARS-CoV-2 virus characterizations by lineage to The European Surveillance System (TESSy), which is curated by ECDC. On 11^th^ June 2023, we downloaded COVID-19 data from TESSy for all 54 European CAT in the WHO Region for the study period.

We conducted our analysis in two parts. For the first part, we analyzed data from the full study period (week 50/2020 to week 12/2023, hereafter, the full study period) for which a defined minimum COVID-19 vaccination data set was available. We included data for six age groups (25-49, 50-59, ≥60, 60-69, 70-79 and ≥80 years) in both analyses. To be included in the analysis, CAT needed to have reported both mortality and vaccination data for at least one of the four older age-groups: ≥60, 60-69, 70-79 and ≥80. Only countries that reported weekly data for both vaccination and mortality by age-group for ≥90% of study weeks in the full study period were included. These data were available for 34/54 CAT *^1^*. The following CAT only reported mortality and vaccination data for people aged ≥60 years for the full study period: Germany (data were collected but could not be made available); the Republic of Moldova and Ukraine. Israel and United Kingdom (Scotland) were only able to report data by the following three age groups (25-49, 50-59 and ≥60 years) (Supplementary Table 1).

As a sub-analysis, we restricted our analysis to the pre-Omicron period (week 50/2020 to 50/2021), which allowed us to include an additional five CAT *^2^*) that had consistently reported data only during the early part of the pandemic or had experienced significant changes in reporting later in the course of the pandemic.

We estimated periods of VOC-predominant circulation by CAT using virological data reported to TESSy. If data were unavailable in TESSy, we downloaded data from the Global Initiative on Sharing All Influenza Data (GISAID) obtained from CoVariants.org(last accessed: November 2023). A VOC was deemed predominant in the Region or in a country if ≥50% of sequences per week were attributed to a given variant. Regionally and at a CAT level, we defined the start of each VOC period (Alpha, Delta and Omicron) as the first week of predominance, and the end of the VOC period as the week prior to the start of the following predominant VOC period. For CATs where variant data was unavailable (Albania, Republic of Moldova and Kosovo^[*1*]^), we estimated the start and end weeks of variant periods as the median start and end week of geographically neighboring CAT.

We downloaded population data for 2021 and 2022 from the United Nations (16) for non-EU/EEA CAT and from Eurostat (17) for EU/EEA countries, except for the following CATs, where more detailed population denominators were available: Estonia (18), Italy (19), Malta (20), North Macedonia (21), Poland (22), Portugal (23), Sweden (24), Switzerland (25), the United Kingdom (England, Scotland and Wales) (26) and Kosovo^[1]^(27).

To identify COVID-19 vaccine effectiveness (VE) studies that estimated VE against mortality by age-group and by VOC, we used the COVID-19 Study Explorer from International Vaccine Access Center (IVAC, https://view-hub.org/vaccine/covid/effectiveness-studies), Johns Hopkins Bloomberg School of Public Health (last accessed on 31^st^ May 2023). We calculated an average VE against death per dose and VOC (summarized in Table 1), by calculating the average of VE estimates against death listed on the COVID-19 Study Explorer. We only included VE estimates against death from studies that: were conducted in adults from the general population; used unvaccinated individuals as the reference group; were conducted in the WHO European Region or other high-income countries, including the United States, Canada, Australia and South Korea; and estimated VE by VOC. We excluded VE estimates from studies that only included people living in long-term care facilities, people with comorbidities, immunocompromised individuals, pregnant women, children and healthcare workers.

**Table 1:**
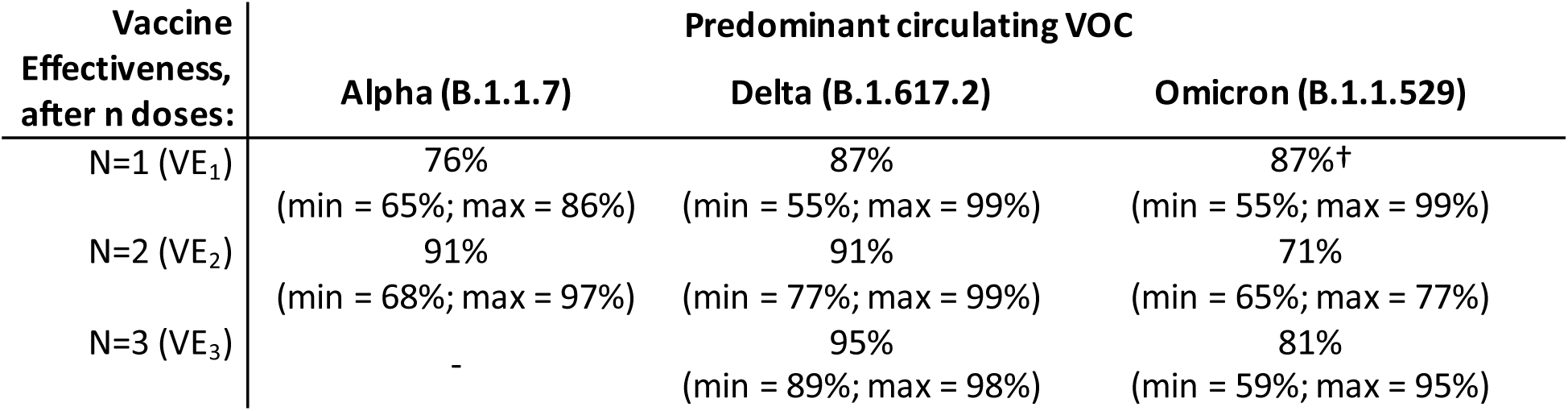
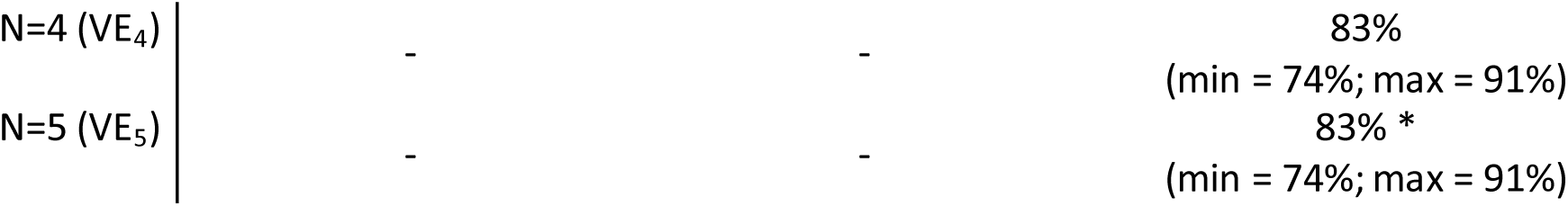
Summary of Vaccine Effectiveness (VE) values against mortality used in the analysis, according to circulating Variant of Concern and dose. Please see Supplementary Table 6 regarding the underlying literature from which the average values below were calculated. The ranges provided in brackets represent the values used for the sensitivity analyses. † Due to the absence of data relating the VE_1_ (Omicron), the values from VE_1_ (Delta) were used. *Due to the absence of data relating the VE_5_ (Omicron), the values from VE_4_ (Omicron) were used

### Data analysis

To estimate the number of lives saved as a result of COVID-19 vaccination in each CAT, we adapted methods previously developed by Machado *et al* (28), as described in Meslé *et al* (12). We used the following parameter definitions: Vaccine Effectiveness against death (**VE_d, t_**) for each respective dose, where d= dose (first dose, second dose, first booster dose, second booster dose and third booster dose) and t= time since vaccination in weeks including a time lag from vaccination to immune protection (described below); and Proportion Vaccinated (**PV_d, t_**) per week per dose (i.e. with no further doses by that week). We calculated a separate VE for each predominant VOC period (Alpha, Delta or Omicron).

Because there were no available VE data for the first vaccine dose during the Omicron period, we used the value VE_1_ Delta as a proxy. We assumed, based on previous studies, that VE declined by 0.25% every week since vaccination, regardless of dose (29). Booster doses (denoted by **VE_3_**to **VE_5_**) were administered from June 2021. The formula we used to calculate the weekly number of lives saved by VOC period, dose and age-group is given below (equations 1 – 3).

To overcome the potential of unreported data in a certain week, we considered the number of deaths in a week as the rolling average (mean) number of deaths observed in the CAT over three consecutive weeks (the relevant week, the prior week, and the following week). For the 11 CAT that had no data on uptake for second and/or third boosters, we calculated an estimated weekly coverage using data from all other reporting CAT and assumed that the time to introduction of these doses was the average time of the introduction of subsequent doses in all other MS that reported data.

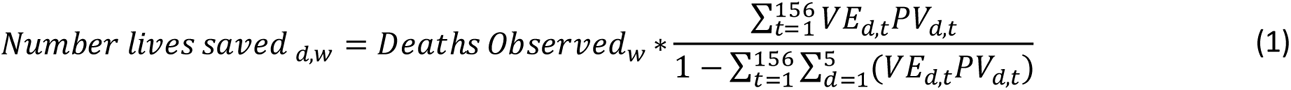

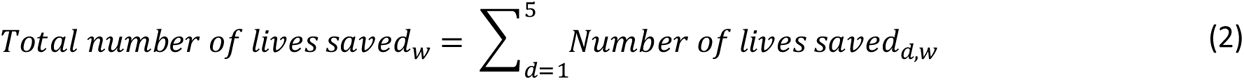

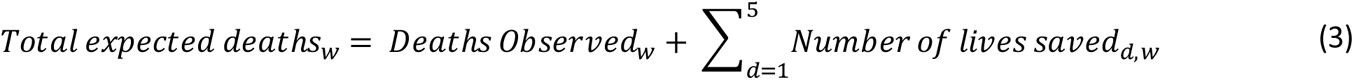

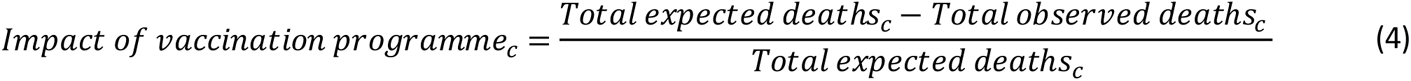

For each CAT, c, we estimated the cumulative expected COVID-19 mortality rate per 100,000 population had no vaccination occurred, as the sum of the observed deaths and of lives saved for each vaccine dose (equation 3). We estimated the impact of the vaccine program on COVID-19 mortality, by age-group and VOC, in each CAT, by calculating the percentage change (equation 4) between observed deaths and expected deaths.

We assumed the following time lags: reporting delay (one week); delay from vaccination to generation of immunoprotection (four weeks (first dose), three weeks (second dose) (35), two weeks (first booster) (30) and one week (for each additional booster) (31).

Several simplifying assumptions were made when using these data parameters. First, we assumed that CATs used the same case definitions when reporting COVID-19 mortality to TESSy. We assumed that reporting delays were similar by time and place; and that VE and waning immunity did not vary across vaccine brands and population groups. Finally, we used the same VE estimates across age groups. Lastly, we also considered the case-fatality percentages per age group using the total number of reported infections and deaths.

We conducted our analyses in R version 4.2.2 (34); the code is available on GitHub.

### Sensitivity analyses

We performed eight sensitivity analyses (SA) to quantify uncertainty around the estimated number of lives saved. In the first two sensitivity analyses, we used the maximum VE (SA1) and minimum VE (SA2) estimates identified through the results of the IVAC search. For the third and fourth SA, we increased the lag time by one week from receipt of vaccine dose to onset of immune protection (SA3) and decreased the lag time by one week (SA4). For the fifth SA (SA5), we decreased the weekly VE waning to 0.1%, and for the sixth SA (SA6), we increased the weekly percent waning to 0.7%, using values described in Wu *et al* (29). Lastly, for SA7 and SA8, we introduced an additional metric – prior infection, since this has been shown to reduce the risk of severe outcome following subsequent infection (33).

Prior infection could result in a reduction in VE if vaccination uptake differs by prior infection. To take into account prior infection, we calculated weekly VE as a weighted average of (alpha*seroprevalence*VE) + (1-seroprevalence)*VE where alpha represents the proportion decrease in VE related to prior infection in the population, and Seroprevalence represents seroprevalence estimates taken from published studies and updated over time, when later seroprevalence estimates were available (34). Both alpha (here a waning factor) and seroprevalence are proportions between zero and one. We used Alpha values of 0.8 (SA7) and 0.95 (SA8).

### Ethical considerations

Because this analysis utilized routinely reported anonymous data aggregated by broad age-groups, no ethical approval was required.

## Results

### Reported mortality

Using the variant data, we defined the following VOC periods for the Region: the Alpha period (3^rd^ January to 6^th^ June 2021, the Delta period (11^th^ July to 5^th^ December 2021) and lastly, the Omicron period (13^th^ December 2021 to 26^th^ March 2023).

Since the start of vaccination programs through March 2023, 29 CAT reported 1,050,501 COVID-19 deaths in people aged ≥25 years to TESSy; of these deaths, 43% (n=447,600) were in people ≥80 years old. In contrast, only 4% and 2% of deaths were in people aged 50-59 and 25-49 years old, respectively. Of the 34 CATs that reported data for people aged ≥60 years reported 990,881 deaths (Figure 1A,Table 2).

**Figure 1:**
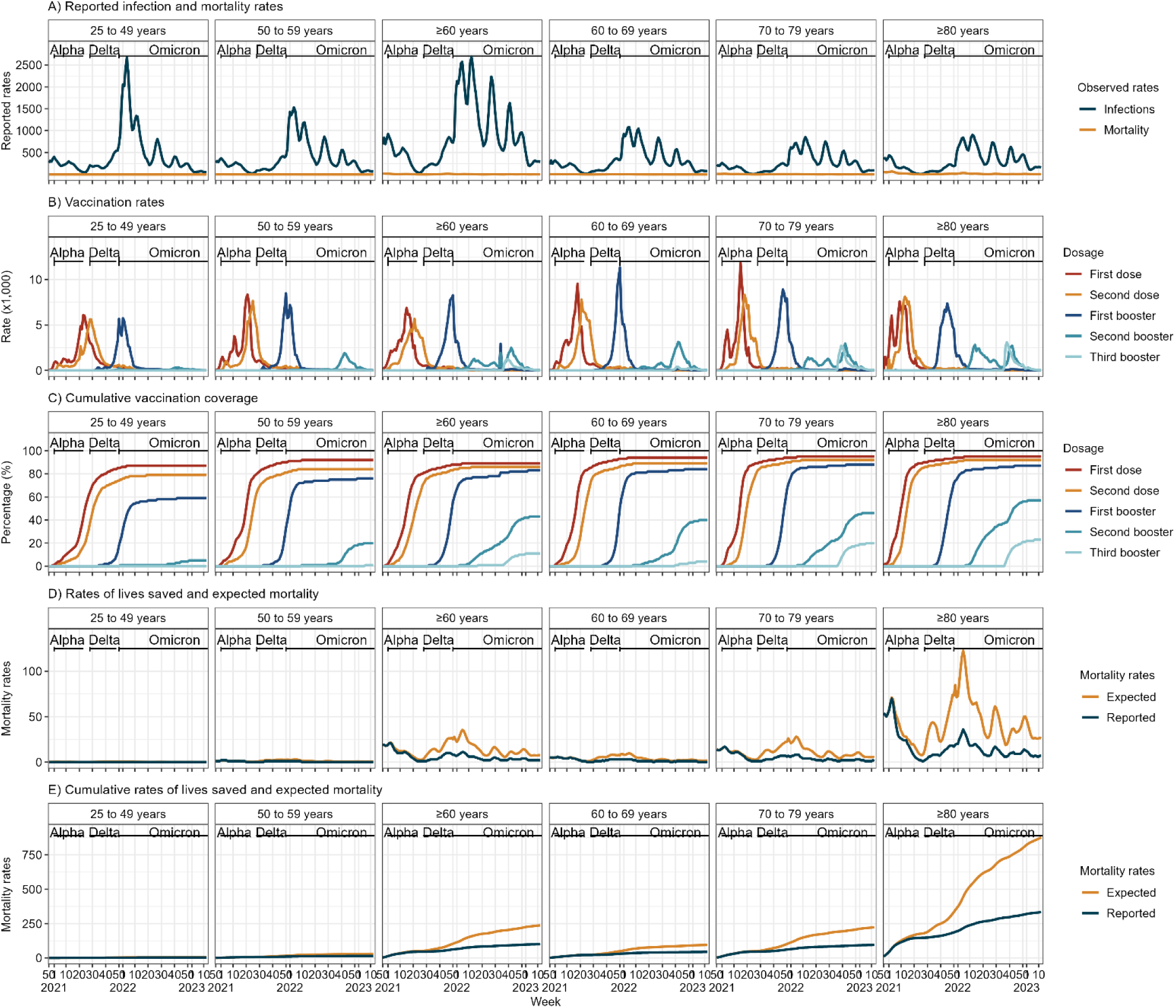
Summary of observed infection and mortality rates (A), vaccination rates (B), cumulative vaccination rates (C), rate of lives saved and expected mortality (D) and cumulative rate of lives saved and expected mortality (E), per age-group in 34 CAT in the WHO European Region in context of circulating Variants of Concern (VOC) (black horizontal lines). Note: all rates are per 100,000 general population and the ≥60 years age group includes data for 34 CAT.

**Table 2:**
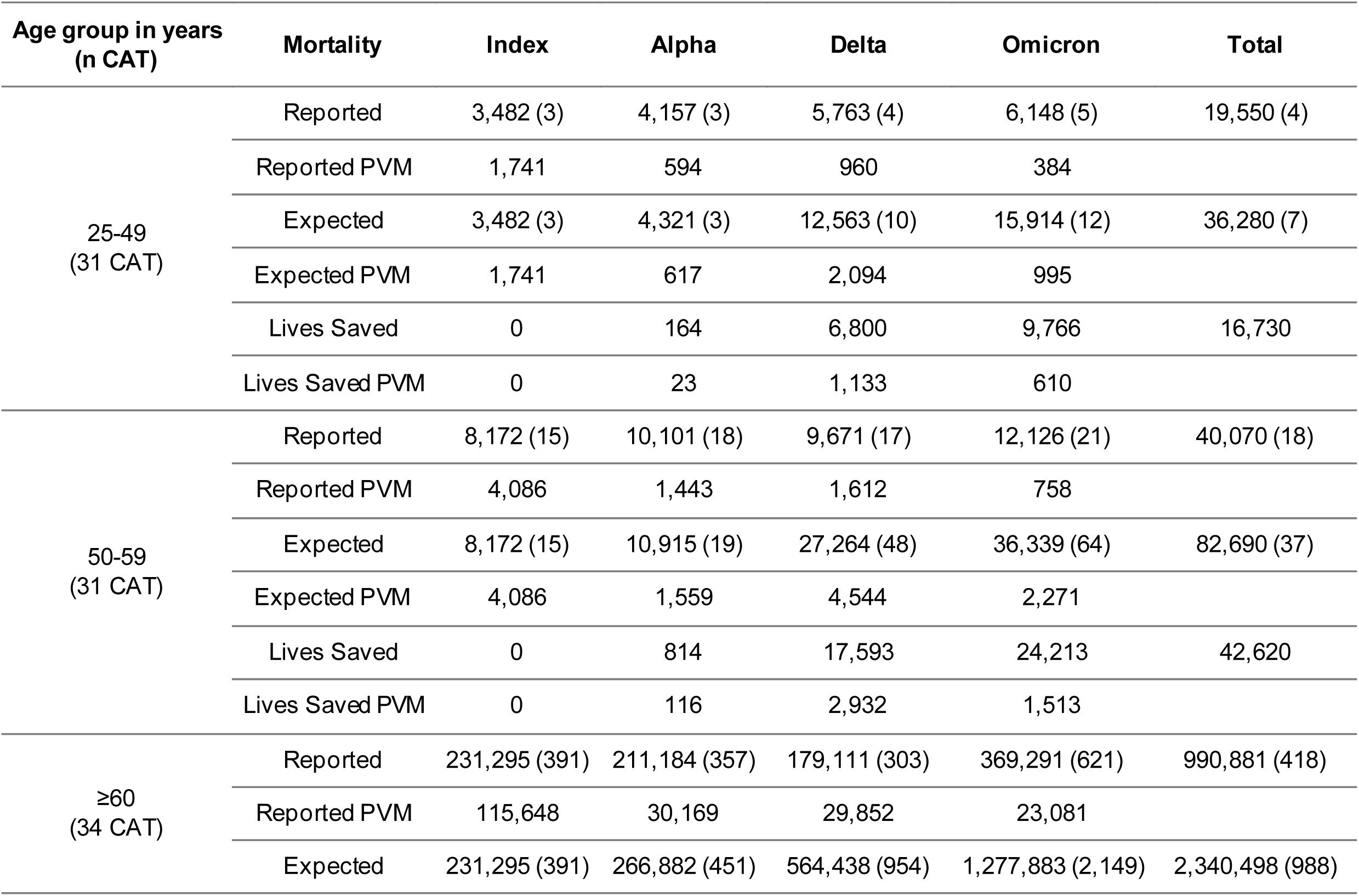
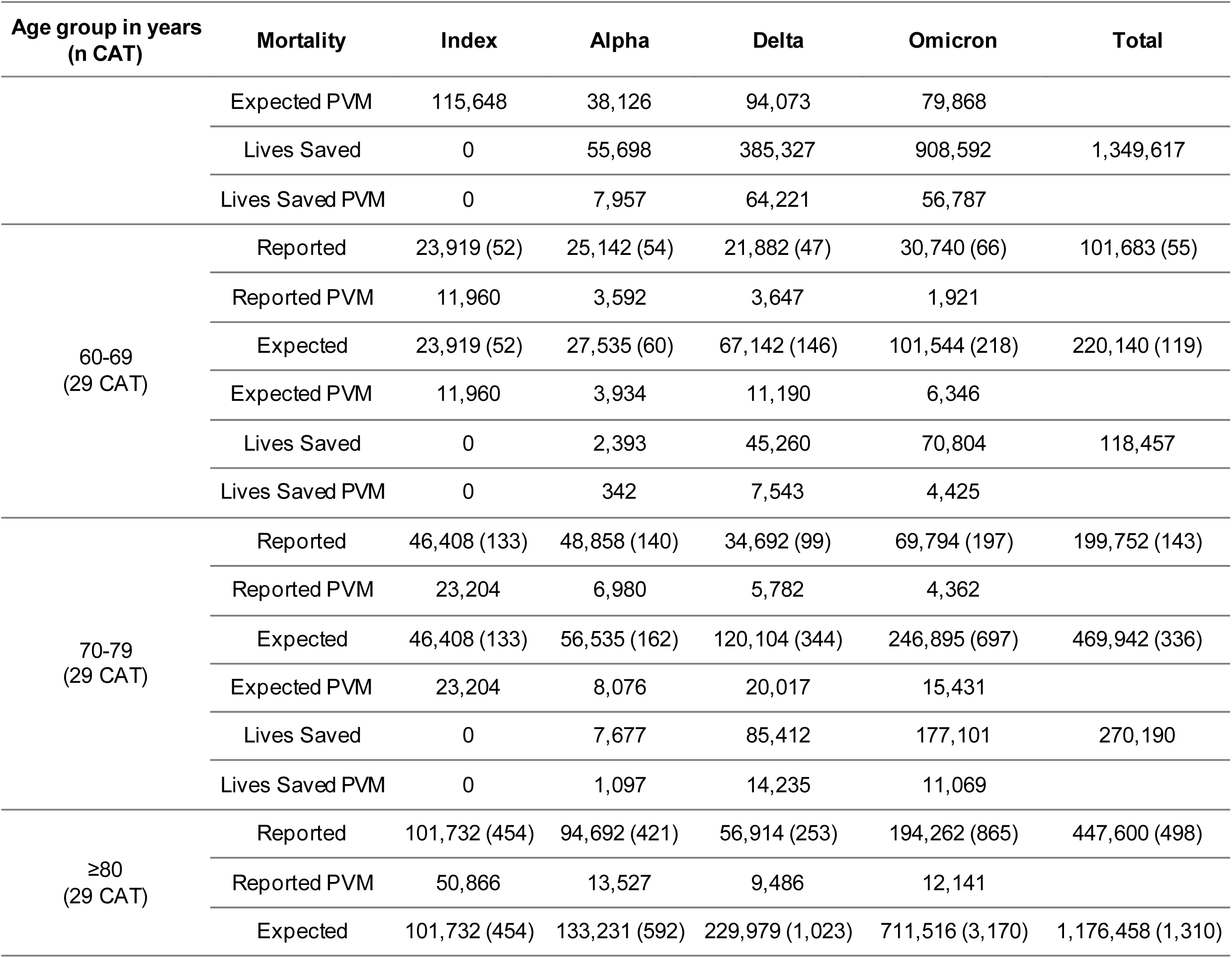
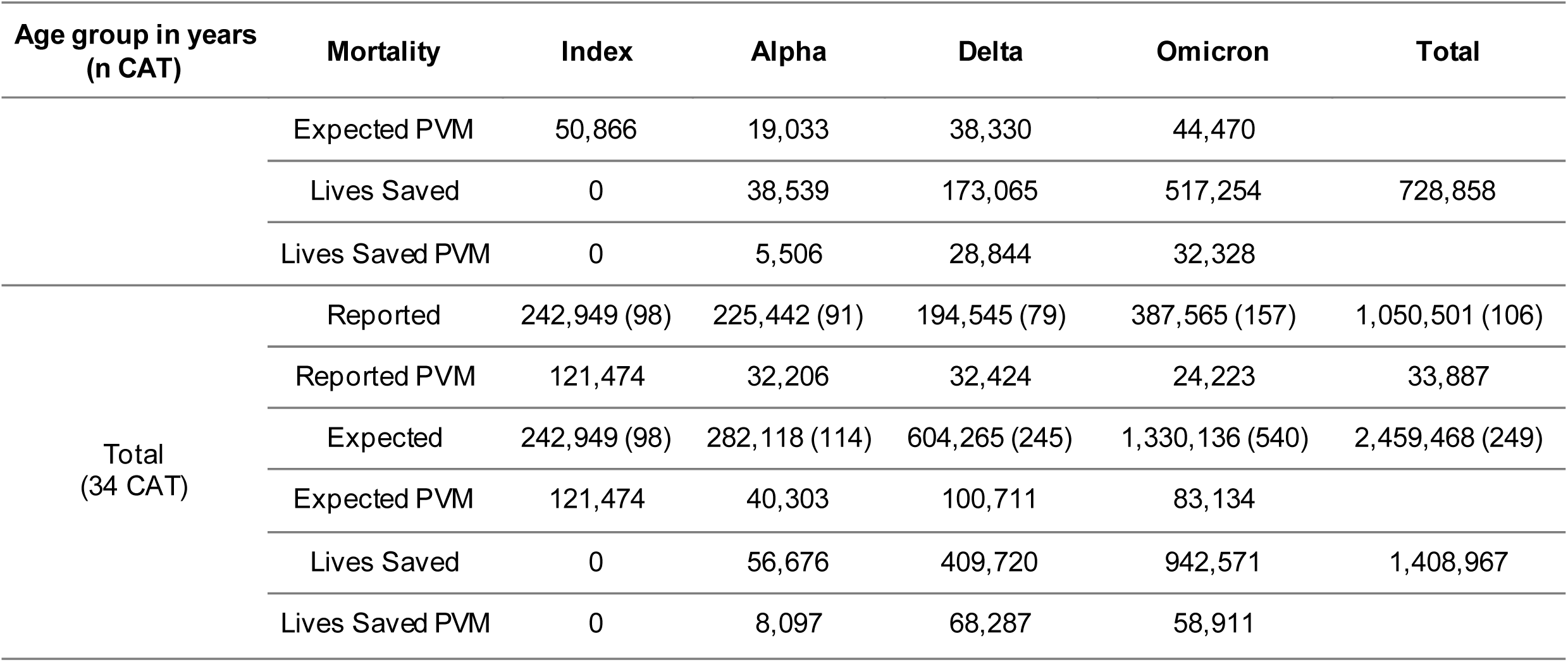
Number of reported, expected deaths and lives saved, cumulatively and Per Variant Month (PVM) with number of countries, areas and territories (CAT) included in each age group; from SARS-CoV-2 infection (rate per 100,000 general population) are shown in brackets, by variant-period and age-group, between weeks 50/2020 and 12/2023, in 34 CAT in the WHO European Region. Notes: the Index variant refers to the original circulating variant, or wild-type; the ≥60 years age group includes data for all 34 CAT; the Total is calculated as the sum of 25-49, 50-59 and ≥60 years age groups. The duration of each VOC dominance is Index: 2 months; Alpha: 7 months; Delta: 6 months and Omicron: 16 months.

The overall cumulative mortality rate for those aged ≥25 years was 109 per 100,000 population and 423 per 100,0000 for those aged ≥60 years. The cumulative mortality rate was 515 per 100,000 for people aged ≥80 years old, 147 per 100,000 population for 70-79 years old, 57 per 100,000 for 60-69 years old. For people aged 50-59 years and 25-49 years, cumulative mortality rates were 18 and 4 per 100,000 population respectively. In contrast, reported infections of any severity were highest in people 25-49 years old (9,355,861 / 153,978,991, 53%) and lowest in people aged ≥80 years old (9,355,861 / 153,978,991, 6%). Only 27% (41,410,536 / 153,978,991) of reported infections occurred among adults aged ≥60 years (Figure 1A, Table 2).

### Vaccination roll-out

Administration of the first booster doses started around week 30/2021 (July 2021). Initially, older age-groups were prioritized. In most CAT, younger individuals became eligible shortly thereafter and second booster doses became available for people in older age-groups in early 2022 (Figure 1Figure 1B, C). By the end of the analysis period (mid-March, 2023), in the 34 CAT overall, coverage in adults ≥25 years old was 87% for the primary vaccine series, 82% for the first booster, 71% for the first booster, 24% for the second booster and 5% for the third (Table 3)Table 2. Coverage for people aged ≥60 years was 89%, 86%, 83%, 43% and 11% for each respective dose and for people aged ≥80 years was 95%, 92%, 87%, 57% and 23% for each respective dose. Across all CATs included in this analysis, coverage was consistently higher in older age groups compared to younger age groups; this difference was even more pronounced for booster doses (Figure 1 and Table 2Table 2).

**Table 3:** Total Vaccine Uptake (VU) and number of lives saved by COVID-19 vaccine dose, variant and age-group, and as percentage by Variant of Concern (VOC) period of total lives saved in brackets, since the start of vaccinations in 34 countries, areas and territories (CAT) in the WHO European Region, with number of countries, areas and territories (CAT) included in each age group. Notes: the ≥60 years age group includes data for all 34 CAT; the Total is calculated as the sum of 25-49, 50-59 and ≥60 years age groups.

|  |  |  | Lives saved by variant and dose |  |  |  |
| --- | --- | --- | --- | --- | --- | --- |
| Age group in years<br>(n CAT) | Dosage | VU (%) | Alpha | Delta | Omicron | Total |
| 25-49<br>(31 CAT) | First dose | 110,256,355 (85%) | 59 (3%) | 840 (49%) | 829 (48%) | 1,728 |
|  | Second dose | 99,416,586 (77%) | 105 (1%) | 5,476 (69%) | 2,374 (30%) | 7,955 |
|  | First booster | 74,448,707 (57%) | 0 (0%) | 469 (7%) | 6,301 (93%) | 6,770 |
|  | Second booster | 6,603,769 (5%) | 0 (0%) | 15 (5%) | 262 (95%) | 277 |
|  | Third booster | 157,866 (0%) | 0 (-%) | 0 (-%) | 0 (-%) | 0 |
|  | Total | 290,883,283 | 164 (1%) | 6,800 (41%) | 9,766 (58%) | 16,730 |
| 50-59<br>(31 CAT) | First dose | 50,488,396 (89%) | 321 (10%) | 1,397 (44%) | 1,430 (45%) | 3,148 |
|  | Second dose | 46,419,517 (82%) | 493 (3%) | 14,298 (82%) | 2,542 (15%) | 17,333 |
|  | First booster | 41,658,728 (74%) | 0 (0%) | 1,898 (9%) | 18,692 (91%) | 20,590 |
|  | Second booster | 11,011,072 (19%) | 0 (0%) | 0 (0%) | 1,549 (100%) | 1,549 |
|  | Third booster | 299,132 (1%) | 0 (-%) | 0 (-%) | 0 (-%) | 0 |
|  | Total | 149,876,845 | 814 (2%) | 17,593 (41%) | 24,213 (57%) | 42,620 |
| ≥60<br>(34 CAT) | First dose | 125,250,761 (89%) | 21,973 (44%) | 10,365 (21%) | 17,316 (35%) | 49,654 |
|  | Second dose | 120,814,004 (86%) | 33,716 (10%) | 267,685 (76%) | 48,754 (14%) | 350,155 |
|  | First booster | 116,303,048 (83%) | 9 (0%) | 107,243 (15%) | 586,510 (85%) | 693,762 |
| Age group in years<br>(n CAT) | Dosage | VU (%) | Alpha | Delta | Omicron | Total |
|  | Second booster | 60,615,440 (43%) | 0 (0%) | 34 (0%) | 166,379 (100%) | 166,413 |
|  | Third booster | 16,096,607 (11%) | 0 (0%) | 0 (0%) | 89,633 (100%) | 89,633 |
|  | Total | 439,079,860 | 55,698 (4%) | 385,327 (29%) | 908,592 (67%) | 1,349,617 |
| 60-69<br>(29 CAT) | First dose | 42,317,982 (91%) | 1,487 (24%) | 2,397 (38%) | 2,348 (38%) | 6,232 |
|  | Second dose | 40,072,892 (86%) | 906 (2%) | 36,206 (87%) | 4,709 (11%) | 41,821 |
|  | First booster | 37,800,945 (81%) | 0 (0%) | 6,657 (11%) | 54,266 (89%) | 60,923 |
|  | Second booster | 17,940,191 (39%) | 0 (0%) | 0 (0%) | 9,265 (100%) | 9,265 |
|  | Third booster | 1,774,343 (4%) | 0 (0%) | 0 (0%) | 216 (100%) | 216 |
|  | Total | 139,906,353 | 2,393 (2%) | 45,260 (38%) | 70,804 (60%) | 118,457 |
| 70-79<br>(29 CAT) | First dose | 32,649,123 (93%) | 3,976 (36%) | 2,762 (25%) | 4,348 (39%) | 11,086 |
|  | Second dose | 31,633,138 (90%) | 3,701 (5%) | 60,621 (84%) | 7,440 (10%) | 71,762 |
|  | First booster | 30,445,716 (86%) | 0 (0%) | 22,028 (15%) | 123,937 (85%) | 145,965 |
|  | Second booster | 15,971,781 (45%) | 0 (0%) | 1 (0%) | 21,341 (100%) | 21,342 |
|  | Third booster | 6,852,090 (19%) | 0 (0%) | 0 (0%) | 20,035 (100%) | 20,035 |
|  | Total | 117,551,848 | 7,677 (3%) | 85,412 (32%) | 177,101 (66%) | 270,190 |
| ≥80<br>(29 CAT) | First dose | 20,919,657 (95%) | 15,479 (55%) | 3,660 (13%) | 9,042 (32%) | 28,181 |
|  | Second dose | 20,369,102 (92%) | 23,051 (14%) | 112,698 (70%) | 26,074 (16%) | 161,823 |
| Age group in years<br>(n CAT) | Dosage | VU (%) | Alpha | Delta | Omicron | Total |
|  | First booster | 19,255,453 (87%) | 9 (0%) | 56,674 (15%) | 313,385 (85%) | 370,068 |
|  | Second booster | 12,680,841 (57%) | 0 (0%) | 33 (0%) | 105,763 (100%) | 105,796 |
|  | Third booster | 5,027,196 (23%) | 0 (0%) | 0 (0%) | 62,990 (100%) | 62,990 |
|  | Total | 78,252,249 | 38,539 (5%) | 173,065 (24%) | 517,254 (71%) | 728,858 |
| Total<br>(34 CAT) | First dose | 285,995,512 (87%) | 22,353 (41%) | 12,602 (23%) | 19,575 (36%) | 54,530 |
|  | Second dose | 266,650,107 (82%) | 34,314 (9%) | 287,459 (77%) | 53,670 (14%) | 375,443 |
|  | First booster | 232,410,483 (71%) | 9 (0%) | 109,610 (15%) | 611,503 (85%) | 721,122 |
|  | Second booster | 78,230,281 (24%) | 0 (0%) | 49 (0%) | 168,190 (100%) | 168,239 |
|  | Third booster | 16,553,605 (5%) | 0 (0%) | 0 (0%) | 89,633 (100%) | 89,633 |
|  | Total | 879,839,988 | 56,676 (4%) | 409,720 (29%) | 942,571 (67%) | 1,408,967 |

### Lives saved

#### Analysis of full study period (December 2020 to March 2023)

We estimated that during the analysis period, for the 34 CAT overall, COVID-19 vaccinationreduced deaths by 57% (CAT range: 15% to 75%) in adults aged ≥25 years. This reduction represents at least 1,408,967 lives saved and a mortality risk reduction of 325 per 100,000 population (Table 2). Among all adults aged ≥60 years, vaccination reduced mortality by 58% (1,349,617 / 2,340,498 lives). The majority (728,858 / 1,408,967 lives, 52%) of the lives saved were in people aged ≥80 years, equivalent to a mortality risk reduction of 812 per 100,000 persons. We found that vaccination reduced mortality by 57% among 70-79 years old, 54% among 60-69 years old, 52% in 50-59 years old and 46% for 25-49 years old (Table 2).

Overall, the first booster saved an estimated 721,122 lives (from 1,408,967, 51%) in adults ≥25 years old. Among persons aged ≥80 years, the first booster saved 370,068 lives (from 728,858), representing a 51% reduction in expected mortality. Among people aged ≥60 years, the first booster reduced mortality by 51% (693,762 / 1,349,617) whereas in those aged 25-49 years, the second dose reduced mortality by 48% (7,955 / 16,730) (Table 3 and Figure 2).

**Figure 2:**
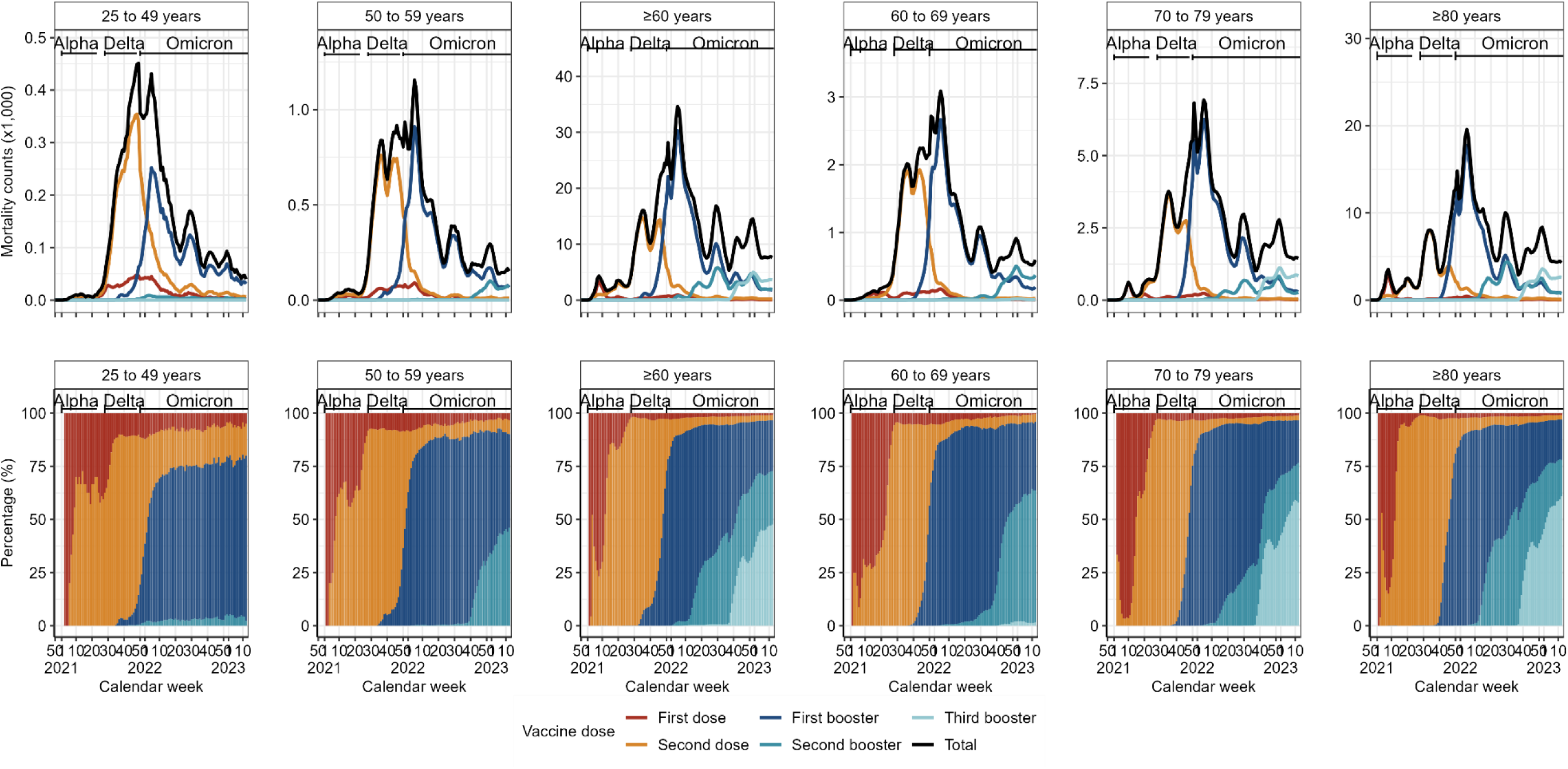
Counts (top row) and percentage (bottom row) of estimated lives saved per vaccine dose per age-groups, between weeks 50/2020 and 12/2023, in 34 countries, areas and territories (CAT) in the WHO European Region. Note: age groups 25-49 and 50-59 years include data for 31 CATs, the ≥60 years age group includes data for 34 CATs and the remaining age groups include data for 29 CATs.

In the analysis of lives saved by predominant VOC, vaccination saved the most lives (942,571 / 1,408,967, 67%) during the Omicron period which lasted 16 months (compared to six months for the Delta period), when vaccination reduced mortality by 71% (mortality risk reduction of 383 per 100,000 population). During the Omicron period, vaccination reduced mortality by 71% in people aged≥80 years (517,254 / 728,858), and by 67% for all adults ≥60 years (908,592 / 1,349,617) (Table 3).

In the analysis of lives saved by CAT, among all adults ≥25 years old, vaccination reduced mortality by the largest proportion in Israel (75%), Malta (72%), Iceland (71%), Denmark and United Kingdom (Scotland) (both 70%) (Table 4). In the CAT analysis by age-group, among people ≥80 years old, vaccination reduced mortality most in Malta (72%), followed by United Kingdom (England) (71%) and Denmark and Iceland (both 70% reduction). For adults aged 25-49 years old, vaccination reduced mortality the most in Malta (68%), Iceland(67%) and Israel (60%) (Supplementary text and Supplementary Table 3). Among all adults, vaccination reduced mortality the least in Ukraine (15% - where data were only available for people ≥60 years), Romania (20%) and Kosovo^[*1*]^ (21%). Among individuals ≥80 years old, the smallest reduction occurred in Romania (12%), Kosovo^[^*^2^*^]^ (17%) and Slovakia (27%) (Supplementary Table 3).

**Table 4:** Cumulative vaccination uptake, number of deaths reported and lives saved, reported and expected mortality rates per 100,000 population aged ≥25 years, by countries, areas and rritories (CAT), for weeks 50/2020 to 12/2023. Notes: CATs have been ordered according to the proportion of lives saved; d_1_ refers to first vaccine dose, d_2_ refers to second doses, d_3_ refers first booster, d_4_ refers to second booster and d_5_ refers to third booster; data presented below for Germany, Republic of Moldova and Ukraine only considers the ≥60 years age group.

| Vaccination uptake |  |  |  |  | Number of lives |  |  |  |  |  |  | Mortality rate per 100,000 |  |  |
| --- | --- | --- | --- | --- | --- | --- | --- | --- | --- | --- | --- | --- | --- | --- |
| Countries, areas and territories | VU<sub>2</sub> | VU<sub>3</sub> | VU<sub>4</sub> | VU<sub>5</sub> | Reported deaths | Saved after d<sub>1</sub> | Saved after d<sub>2</sub> | Saved after d<sub>3</sub> | Saved after d<sub>4</sub> | Saved after d<sub>5</sub> | Total Saved | Reported | Total Expected | % change |
| Israel<sup>1</sup> | 95 | 78 | 19 | <1% | 9,397 | 63 | 6,383 | 15,691 | 6,470 | 24 | 28,631 | 195 | 787 | 75 |
| Malta | 90 | 84 | 23 | 3 | 504 | 1 | 352 | 708 | 205 | 0 | 1,266 | 96 | 338 | 72 |
| Iceland | 79 | 88 | 35 | 10 | 235 | 0 | 6 | 441 | 121 | 2 | 570 | 70 | 240 | 71 |
| Denmark | 93 | 84 | 56 | 1 | 7,119 | 0 | 1,808 | 11,440 | 3,440 | 78 | 16,766 | 124 | 415 | 70 |
| United Kingdom (Scotland) | 94 | 84 | 50 | 13 | 10,527 | 339 | 6,474 | 11,915 | 4,081 | 1,531 | 24,340 | 262 | 869 | 70 |
| Finland | 91 | 77 | 41 | 18 | 8,508 | 170 | 2,481 | 8,302 | 6,478 | 1,768 | 19,199 | 149 | 484 | 69 |
| United Kingdom (England)<sup>1,2</sup> | 93 | 82 | 41 | 25 | 174,800 | 19,839 | 80,578 | 164,580 | 57,638 | 73,897 | 396,532 | 325 | 1,061 | 69 |
| Cyprus | 84 | 73 | 15 | 3 | 1,490 | 92 | 1,105 | 1,472 | 173 | 16 | 2,858 | 175 | 510 | 66 |
| Spain | 84 | 76 | 35 | <1% | 70,258 | 3,017 | 48,581 | 76,636 | 7,085 | 11 | 135,330 | 145 | 424 | 66 |
| Belgium | 88 | 82 | 51 | 8 | 15,921 | 981 | 7,019 | 17,208 | 2,854 | 1,141 | 29,203 | 140 | 397 | 65 |
| Ireland | 94 | 84 | 45 | 18 | 6,002 | 134 | 2,420 | 4,191 | 1,737 | 878 | 9,360 | 135 | 345 | 61 |
| Luxembourg | 77 | 72 | 23 | <1% | 754 | 18 | 248 | 763 | 146 | 0 | 1,175 | 125 | 320 | 61 |
| Austria | 82 | 74 | 30 | 1 | 17,218 | 993 | 7,075 | 14,622 | 2,135 | 34 | 24,859 | 189 | 462 | 59 |
| Italy | 88 | 88 | 19 | 2 | 131,583 | 8,918 | 34,411 | 121,224 | 22,268 | 941 | 187,762 | 206 | 500 | 59 |
| Countries, areas and territories | VU<sub>2</sub> | VU<sub>3</sub> | VU<sub>4</sub> | VU<sub>5</sub> | Reported deaths | Saved after d<sub>1</sub> | Saved after d<sub>2</sub> | Saved after d<sub>3</sub> | Saved after d<sub>4</sub> | Saved after d<sub>5</sub> | Total Saved | Reported | Total Expected | % change |
| Netherlands | 81 | 76 | 44 | 25 | 12,990 | 360 | 13,068 | 3,540 | 1,492 | 477 | 18,937 | 75 | 183 | 59 |
| Greece | 81 | 72 | 17 | 3 | 33,445 | 2,250 | 14,540 | 26,391 | 3,385 | 399 | 46,965 | 303 | 728 | 58 |
| Portugal | 84 | 85 | 48 | 7 | 21,347 | 2,218 | 5,520 | 16,759 | 3,660 | 1,627 | 29,784 | 190 | 456 | 58 |
| France | 90 | 78 | 27 | 6 | 98,828 | 4,724 | 27,329 | 72,350 | 13,832 | 1,635 | 119,870 | 149 | 330 | 55 |
| Germany<sup>1</sup> | 90 | 85 | 39 | 8 | 135,420 | 2,790 | 50,738 | 89,400 | 18,661 | 4,837 | 166,426 | 554 | 1,236 | 55 |
| Sweden<sup>1</sup> | 87 | 74 | 47 | 2 | 16,109 | 152 | 2,440 | 6,954 | 8,821 | 187 | 18,554 | 158 | 339 | 53 |
| Switzerland<sup>1,2</sup> | 78 | 81 | 13 | 2 | 8,681 | 248 | 3,154 | 5,011 | 357 | 87 | 8,857 | 99 | 201 | 51 |
| Estonia | 69 | 49 | 13 | <1% | 2,633 | 123 | 1,103 | 1,280 | 123 | 0 | 2,629 | 192 | 384 | 50 |
| Croatia<sup>1</sup> | 68 | 39 | 3 | 1 | 4,805 | 235 | 1,513 | 1,990 | 55 | 17 | 3,810 | 118 | 212 | 44 |
| Czechia | 75 | 58 | 13 | <1% | 33,292 | 1,420 | 10,032 | 14,119 | 784 | 0 | 26,355 | 314 | 562 | 44 |
| Hungary | 73 | 54 | 8 | <1% | 41,228 | 992 | 9,202 | 21,623 | 1,218 | 0 | 33,035 | 419 | 754 | 44 |
| Lithuania<sup>1</sup> | 68 | 43 | 2 | <1% | 8,195 | 547 | 4,298 | 1,715 | 1 | 0 | 6,561 | 283 | 509 | 44 |
| Slovenia<sup>1</sup> | 63 | 44 | 6 | 1 | 7,252 | 218 | 2,003 | 3,130 | 144 | 46 | 5,541 | 333 | 587 | 43 |
| Latvia<sup>1</sup> | 62 | 39 | 6 | 1 | 5,659 | 770 | 1,855 | 1,173 | 5 | 0 | 3,803 | 294 | 491 | 40 |
| North Macedonia | 60 | 15 | 1 | <1% | 7,592 | 89 | 2,757 | 492 | 0 | 0 | 3,338 | 433 | 623 | 30 |
| Slovakia | 60 | 44 | 2 | <1% | 17,768 | 207 | 3,631 | 3,643 | 0 | 0 | 7,481 | 334 | 475 | 30 |
| Countries, areas and territories | VU<sub>2</sub> | VU<sub>3</sub> | VU<sub>4</sub> | VU<sub>5</sub> | Reported deaths | Saved after d<sub>1</sub> | Saved after d<sub>2</sub> | Saved after d<sub>3</sub> | Saved after d<sub>4</sub> | Saved after d<sub>5</sub> | Total Saved | Reported | Total Expected | % change |
| Republic of Moldova<sup>1,2</sup> | 39 | 22 | 2 | <1% | 5,983 | 281 | 1,191 | 253 | 0 | 0 | 1,725 | 844 | 1,087 | 22 |
| Kosovo<sup>[1]</sup> | 65 | 11 | 0 | <1% | 1,699 | 10 | 407 | 33 | 0 | 0 | 450 | 129 | 163 | 21 |
| Romania<sup>1</sup> | 39 | 12 | 0 | <1% | 51,144 | 1,520 | 10,355 | 1,050 | 0 | 0 | 12,925 | 271 | 339 | 20 |
| Ukraine<sup>1,2</sup> | 38 | 55 | 27 | <1% | 82,115 | 811 | 11,366 | 1,023 | 870 | 0 | 14,070 | 897 | 1,051 | 15 |
| Total | 83 | 74 | 29 | 7 | 1,050,501 | 54,530 | 375,443 | 721,122 | 168,239 | 89,633 | 1,408,967 | 243 | 568 | 57 |

<sup>1</sup> Booster doses were not reported by the CAT for at least one age group and so were calculated using the average percentage change of vaccination coverage from CAT with booster 2 and 3 reporting.
<sup>2</sup> It was not possible to differentiate booster doses 2 and 3 from booster 1. The same method as above (1) was used for these CAT.
<sup>[1]</sup> All references to Kosovo in this document should be understood to be in the context of the United Nations Security Council resolution 1244 (1999).

In CATs that reached ≥90% vaccine coverage in people ≥60 years old by the early stages of the Delta period (Belgium, Denmark, Iceland, Ireland, Israel, Malta, Netherlands (Kingdom of the), United Kingdom (England, Scotland)) the proportionate reduction in mortality was the highest. In these CATs, vaccination reduced deaths by >60% in people aged ≥25 years. Conversely, in CAT where vaccination coverage was lower than 50%, such as Romania, Republic of Moldova and Kosovo[*^1^*] reduced deaths by ≤30% (Supplementary Figure 1).

#### Sub-analysis of pre-Omicron period (weeks 50/2020 to 50/2021)

In the analysis of the pre-Omicron period, which included 39 CAT (listed in Methods), we estimated that 443,041 lives were saved among people ≥25 years old (representing a 40% mortality risk reduction). For all adults ≥25 years, vaccination reduced mortality the most in Israel (75%), Iceland (71%) and Malta and Norway (both 65%), whereas vaccination reduced mortality the least in Ukraine (10% - among ≥60 years only), the Republic of Moldova and Kosovo^[*1*]^ (both 17%). In this analysis, among people aged ≥60 years old a total of 418,354 lives (94% of expected deaths) were saved (a 40% reduction) nearly half of all lives saved (201,758 lives; 46%) were among people the ≥80 years old; of these, 80% were saved during the Delta period (Supplementary Table 3).

### Sensitivity analyses

When we used the highest VE values (SA1), we estimated that vaccination saved 2,618,085 lives; when we used the lowest VE values (SA2), we estimated that vaccination saved 747,248 lives. When we used shorter lag times (VE3), we estimated 1,323,593 lives were saved, whereas the longer lag times (SA4) saved 1,530,936 lives. When we used a longer vaccination waning time (SA5), we estimated 1,554,472 lives saved whereas a shorter vaccination waning time (SA6 resulted in 1,091,492 lives saved (Figure 3). When we adjusted for a higher level of prior immunity (SA7), we found that vaccination saved 1,028,530 lives. When we assumed a lower level of prior immunity (SA8), we estimated that vaccination saved 1,294,035 lives. Detailed results can be found in Figure 3Supplementary Table 5 and Supplementary Table 6.

**Figure 3:**
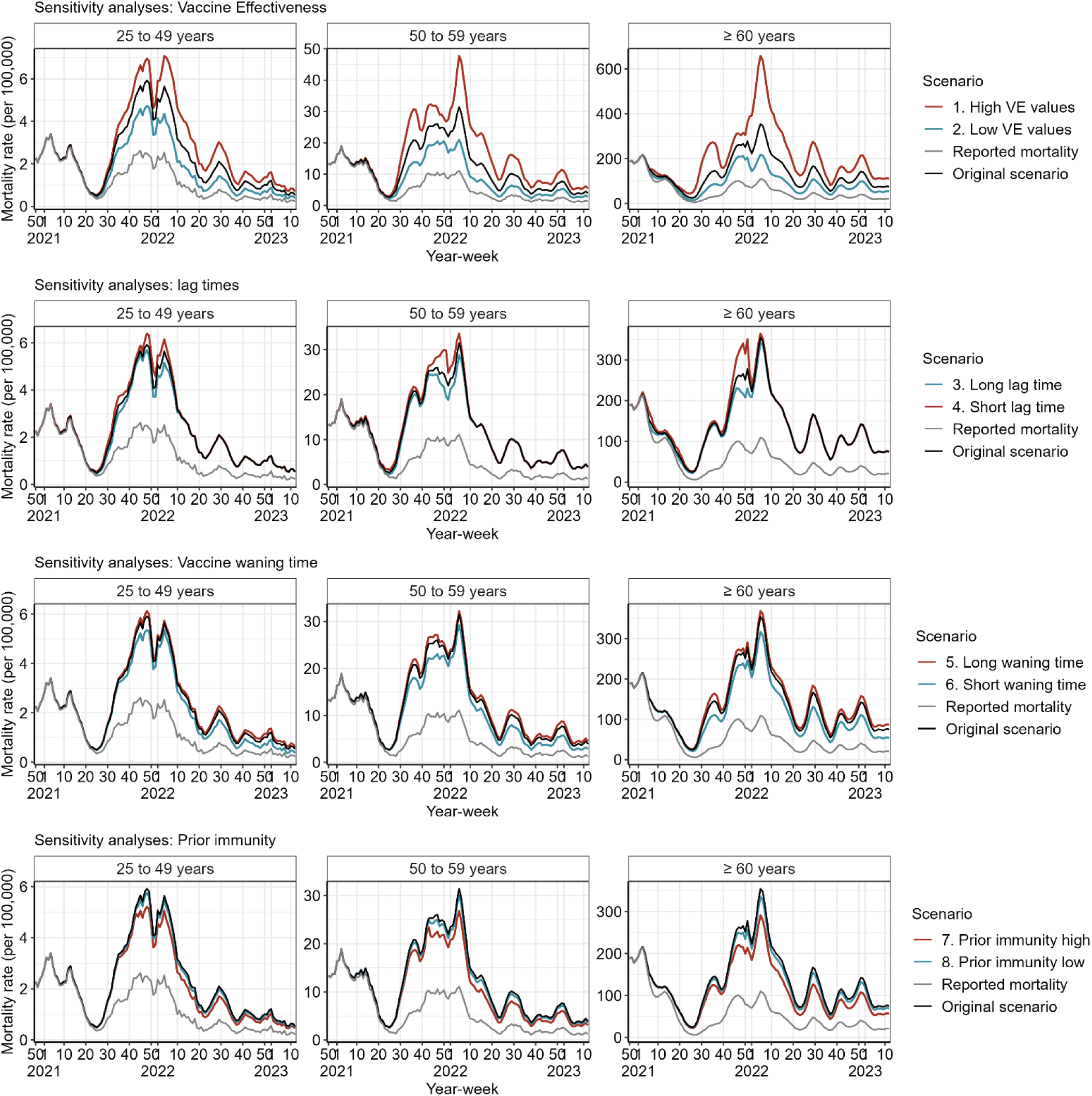
Summary of each sensitivity analysis compared to the original scenario and observed mortality, per age-group, in 34 countries, areas and territories (CAT) in the WHO European Region. Regional and country results are shown in Supplementary Tables 4 and 5. Note: the ≥60 years age group includes CAT that have only reported ≥60 years data and those reporting by finer age groups (namely, 50-59, 60-69 and ≥80 years).

## Discussion

We found that over nearly 2 ½ years of the pandemic, COVID-19 vaccination programs across 34 CAT of the WHO European Region reduced COVID-19 mortality by an estimated 57%, saving approximately 1.4 million lives. In the 34 CAT, the number of lives saved ranged from 450 to 396,532, and mortality was reduced from 15% to 75% by CAT. Our study, the first to estimate the number of lives saved from COVID-19 vaccination in the WHO European Region during a period that encompasses nearly the majority of the pandemic period, underscores the important role COVID-19 vaccines have played in reducing mortality, and adds to previous studies in Europe (12) and globally (8) that have described the profound impact vaccination programs have had on mitigating the impact of the COVID-19 pandemic.

Our study findings are consistent with those from other studies have described lives saved from vaccination during the pandemic, including our previous article (12), where we estimated that COVID-19 vaccination reduced COVID-19 mortality in Europe by 51% in the first 12 months of the pandemic. A study of the global impact of the first year of vaccination found that vaccination had decreased mortality by 63% globally (8). A modeling study of potential deaths averted in low-income and lower-middle-income countries estimated that 1.5 million lives could have been saved in 2022, when Omicron started to circulate, were vaccination to be scaled up in these countries (35).

Our study findings highlight that even during the Omicron period, when the severity of infections decreased relative to earlier periods of previous VOC circulation (10), COVID-19 vaccines still dramatically reduced mortality. We found that most lives were saved (67%) during the Omicron period, the longest VOC period, when Omicron BA.1 and BA.2 began to circulate widely despite already high vaccination coverage. Widespread infections occurred during the Omicron period likely due to a combination of lack of immunity, waning immunity against infection and a highly transmissible virus.

This added to the level of population immunity to provide hybrid immunity resulting in a reduction in infection-severity (36). Our findings likely reflect the ongoing effectiveness of COVID-19 vaccine in protecting against severe disease during this period of Omicron circulation when infection rates were extremely high even with this reduction in infection severity.

We found the highest impact of vaccine was in adults ≥60 years-old, with 96% of all COVID-19 deaths averted by vaccine in 34 CAT occurred in this age group, even though only 27% of reported infections in adults occurred in people in this age group. Similarly, adults aged ≥80 years accounted for 52% of all lives saved through vaccination, only 6% of reported SARS-CoV-2 infections occurred in this age group.

Other studies have found that COVID-19 vaccine has disproportionately saved the lives of older adults; similar proportions of lives saved in those aged ≥60 years, ranging from 70% to 90% (3,5,11,12).

In our analysis, booster doses in older age-groups played an important role in saving lives: early introduction of the first booster dose, which was recommended because of concerns about waning protection against severe disease (8), prevented 1,349,617 deaths in adults ≥60 years old. Indeed, in adults ≥60 years old, the first booster dose was responsible for the majority of lives saved, highlighting the importance of up-to-date vaccination in this high-risk age group. Our results support the recently published Strategic Advisory Group of Experts (SAGE) recommendations for COVID-19 vaccination in the context of Omicron and high population immunity (37). This message is particularly urgent at a time when COVID continues to circulate, and cause morbidity and mortality, across the European Region(the European Respiratory Virus Surveillance Summary (ERVISS, https://erviss.org/)), withrecent booster uptake concerningly low, particularly among high risk groups (7).

The strengths of this study include the availability of weekly, age stratified data from the majority of CAT within the same geographic region. The temporal granularity of these data allowed us to understand the weekly evolution of the pandemic. CAT reported data to TESSy using a common reporting protocol with standardized definitions, allowing for comparisons betweencountries, age-groups and during different phases of the pandemic. Additionally, by using the number of reported deaths for our calculations, the indirect impact of the introduction of PHSM and measures to reduce transmission and mortality were already incorporated.

As in our previous analysis (12), our work has several limitations, namely around complete adjustment for confounders, validity of the underlying data and a few aspects of our methodology. First, regarding confounders, we were not able to adjust for the effects of healthcare system capacities, sociodemographic variations such as deprivation and ethnicity, or the use of antivirals and other medication on mortality. Second, we were not able to stratify vaccine effectiveness by vaccine type, brand or age-group. We were also not able to differentiate the extent of waning immunity following vaccination disaggregated by dose. Moreover, we were not able to adjust for reporting biases - under-reporting likely occurred early in the pandemic due to lack of tests available and later due to unreported self-testing. There was likely variability in the sensitivity of surveillance by CAT which we could not account for, and we could not account for any overreporting because of potential misclassification of deaths later in the pandemic when vaccine escape became more widespread, though CAT were asked to report only deaths where deaths were not otherwise explained by COVID-19. CAT in the Eastern part of the Region were more likely to have under-reported their COVID-19 mortality counts (3); therefore, the true number of lives saved by vaccination in these CAT is likely higher than what we estimated.

Furthermore, we attempted to address the role of prior infections in our sensitivity analysis by varying VE against mortality (SA1 and SA2) and varying presumed levels of prior infection (SA7 and SA8), particular to account for widespread vaccine escape that occurred during the Omicron period. These SAs aimed to consider possible differential susceptibility linked to changes in vaccine uptake according to prior infection history and potentially less virulent VOCs (38), though we assumed that our base approach took this into account by using the observed mortality rates and VE. Finally, we assumed that data reporting was comparable among CAT both in terms of reporting delays and methodology (including whether deaths were caused by or with SARS-Cov-2 infection) and that vaccination intervals between doses were comparable. Data for the fifth and the fourth vaccination doses were not always available and had to be calculated for some CAT. Variant periods were calculated differently in our methodology (predominance determined by ≥50% of weekly sequences of a given VOC) compared to studies calculating predominance at ≥70% of weekly sequences. We calculated vaccination coverage data using number of doses as reported to WHO by CAT, and this reporting method may have varied among countries due to the diverse vaccines and dose regimens used across the Region.

In conclusion, over nearly 2.5 years, most lives saved by vaccination in Europe were in older adults (>60 years), by first booster dose and during the Omicron period, reinforcing the importance of up-to-date vaccination among these most at-risk individuals. Further modelling work should evaluate indirect effects of vaccination and public health and social measures.

## Supporting information

COVID_19_lives_saved_by_vaccination_supplementary_final

## Data Availability

All data produced in the present study are available upon reasonable request to the authors.

## Acknowledgements

We gratefully acknowledge the input and feedback from colleagues at the European Center for Disease Prevention and Control (ECDC), namely, Nathalie Nicolay, Nick Bundle, Sabrina Bacci and Edoardo Colzani and the input of national public health staff involved in surveillance activities and data submission to TESSy.

The authors would like to thank all the countries, areas and territories (CAT) for the provision of mortality and vaccinationdata, including:

- Czechia: Jiri Jarkovsky, Helena Jirincova, Helena Sebestova, Pavel Slezak, Timotej Suri, Iva Vlckova and Jan Zofka
- France: Anna Maisa
- Germany: We thank Felix Günther and Frank Sandmann (both of the Robert Koch Institute, Berlin, Germany) for helpful discussions and comments.
- Israel: Rivka Rich and Noy Pardo (Israel Ministry of Health)
- Italy: Antonino Bella, Andrea Cannone, Martina Del Manso, Massimo Fabiani, Maria Teresa Palamara, Daniele Petrone, Patrizio Pezzotti and Marco Tallon
- Luxembourg: Dritan Bejko, Bruno Consbruck, Corinna Ernst and Guy Weber
- Netherlands (Kingdom of): Rijksinstituut voor Volksgezondheid en Milieu (RIVM)
- Portugal: Maria Leonor Caleiro, Pedro Casaca and Eugénia Fernandes
- Spain: Epidemiological Surveillance and Vaccination Departments of Autonomous Communities
- Switzerland: Federal Office of Public Health, Bern
- United Kingdom (England): Colin Campbell
- United Kingdom (Scotland): Ross Phillips

## Disclaimer

The authors alone are responsible for the views expressed in this article and they do not necessarily represent the views, decisions or policies of the institutions with which they are affiliated. The code for this analysis has been made publicly available and is hosted on the GitHub website at this address: whocov/WHO_EURO_lives_saved_COVID_vaccination: Repository relating to the publication entitled: Estimated number of lives directly saved by COVID-19 vaccination programs in the WHO European Region, December 2020 to March 2023. (github.com)

## Conflict of interest

Dr. Dabrera reports the predecessor of the organization he works for, Public Health England (PHE), received an unrestricted grant from GSK to undertake a study on the outcome of patients who received parenteral zanamavir. The funder received data and interim reports from PHE but did not influence analysis and reporting of the study. Gavin Dabrera had no involvement in the GSK-funded study on parenteral zanamavir. Furthermore, the currently submitted work was part of the public health response activities to COVID-19 and had no relationship to GSK nor the study on parenteral zanamivir.

None to declare otherwise

## Footnotes

1 Austria, Belgium, Croatia, Cyprus, Czechia, Denmark, Estonia, Finland, France, Germany, Greece, Hungary, Iceland, Ireland, Israel, Italy, Latvia, Lithuania, Luxembourg, Malta, Netherlands (Kingdom of the), North Macedonia, Portugal, Republic of Moldova, Romania, Slovakia, Slovenia, Spain, Sweden, Switzerland, Ukraine, United Kingdom (England), United Kingdom (Scotland) and Kosovo (all referencesto Kosovo in this document should be understood to be in the context of the United Nations Security Councilresolution 1244 (1999)).

2 Albania, Montenegro, Norway, Poland and United Kingdom (Wales).

[1] All referencesto Kosovo in this document should be understoodto be in the context of the United Nations Security Council resolution 1244 (1999).

