## Supplementary material for "Estimated number of lives directly saved by COVID-19 vaccination programs in the WHO European Region, December 2020 to March 2023": COVID_19_lives_saved_by_vaccination_supplementary_final

### Additional text

#### Results

Reported mortality

Different circulating variants had varying impact on cumulative COVID-19 mortality rates in all age-groups, with Omicron causing the highest cumulative mortality risk (157 per 100,000) in all age-groups, and Alpha (91 per 100,00) and Delta causing the lowest (79 per 100,00). Although Omicron caused the highest number of infections, it has been the longest variant period recorded, the highest number of reported deaths per variant months were during the Delta period (**Error! Reference source not found.** A, D, Table 2). The reported cumulative mortality risk per 100,000 population in those aged ≥60 years ranged from 303 per 100,000 during the Delta period to 621 per 100,000 during the Omicron period. For those aged ≥80 years, cumulative mortality rate ranged from 253 per 100,000 (Delta) to 865 per 100,000 (Omicron) (**Error! Reference source not found.**).

Important differences in cumulative COVID-19 mortality risk over the study period were visible across the countries of the Region. Until week 12/2023, North Macedonia recorded the highest cumulative mortality risk (1,473 per 100,000) among those aged ≥60 years, followed by Hungary (1,401 per 100,000) and Slovakia (1,218 per 100,000). The countries with the lowest cumulative COVID-19 mortality rate in this age-group (≥60 years) were in the Netherlands (Kingdom of) (267 per 100,000) followed by Iceland (295 per 100,000) and Malta (372 per 100,000). The cumulative age-specific mortality rates for 60-69; 69-79 and >80 years are presented in Supplementary Table 2.

Among the 50 to 59 years age-groups, North Macedonia (282 per 100,000), Hungary (270 per 100,000) and Slovakia (194 per 100,000) recorded the highest cumulative mortality risk per 100,000 population. The countries with the lowest mortality risk in these age-groups were Iceland, Netherlands (Kingdom of the) (both 14 per 100,000) followed by Malta (17 per 100,000). Among the 25 to 49 years age-groups, North Macedonia (64 per 100,000), Hungary (52 per 100,000) and Latvia, Romania (both 36 per 100,000) recorded the highest mortality risk per 100,000 population. The countries with the lowest cumulative mortality risk in these age-groups were Belgium and Switzerland (both 3 per 100,000), the Netherlands (Kingdom of the) (2 per 100,000) and Iceland (1 per 100,000) (Supplementary Table 3).

Vaccination roll-out

When considering vaccination coverage by countries, areas and territories (CAT) the highest complete series vaccination rate administered in those aged ≥60 years was seen in Ireland, Israel, United Kingdom (England, Scotland) (all >99%), followed by Denmark (99%) and Malta (97%). Those with the lowest vaccination coverage in this age-group were Romania (40%), Republic of Moldova (39%) and Ukraine (38%). When considering the youngest age-group, those with the highest coverage of complete series were France and Israel (both 89%), Ireland and Italy (both 85%), whereas Slovakia (50%), Slovenia (47%) and Romania (37%) had the lowest uptake (Supplementary Table 3).

The vaccination programs that were rolled out fastest were in Belgium, Denmark, Iceland, Ireland, Israel, Malta, Netherlands (Kingdom of the) and United Kingdom (England, Scotland), who vaccinated at least 90% of their population aged ≥60 years and older with at least one dose within 30 weeks, the slowest were Republic of Moldova, North Macedonia, and Kosovo[1] (Supplementary Figure 1).

[1] All references to Kosovo in this document should be understood to be in the context of the United Nations Security Council resolution 1244 (1999).

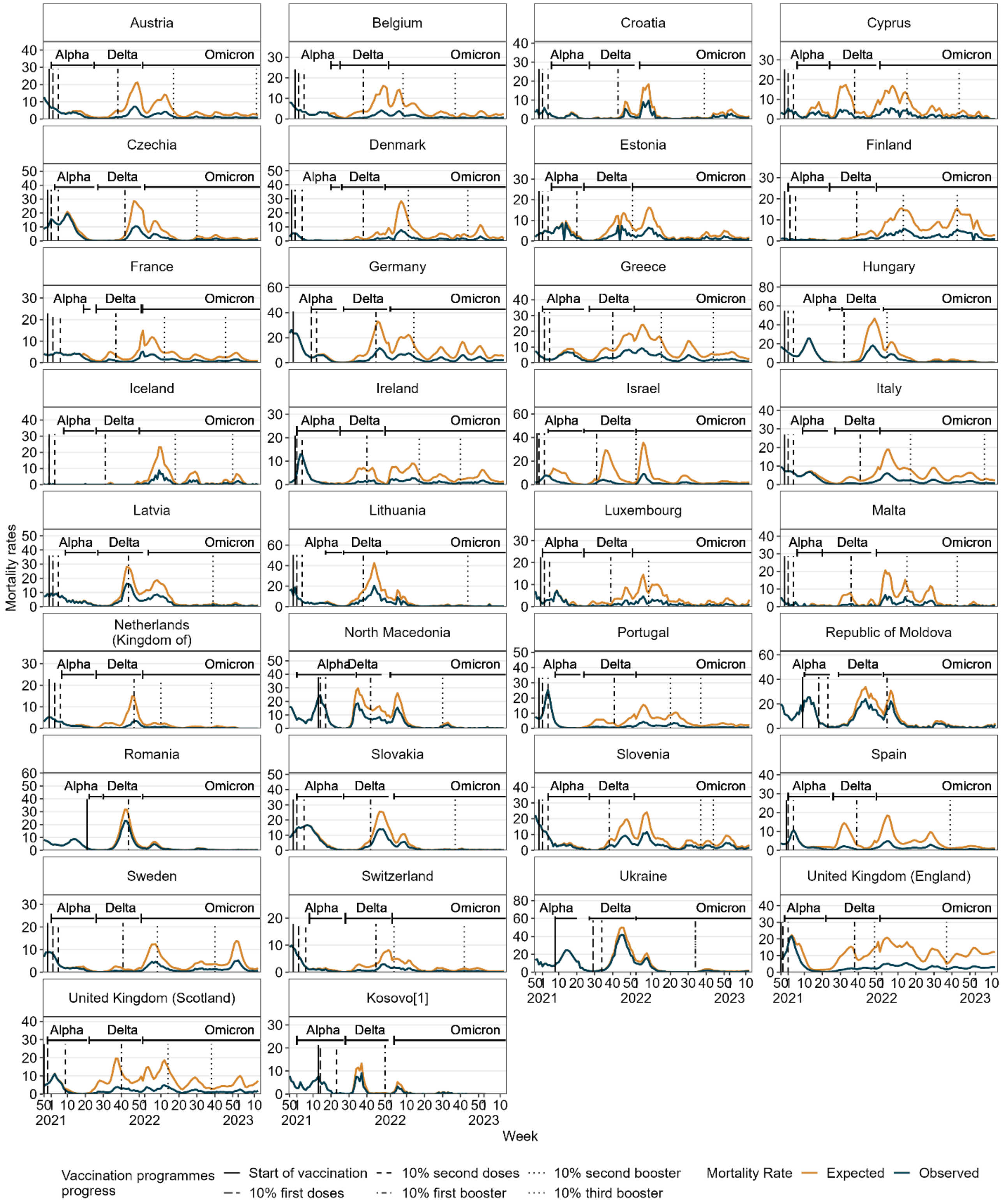


[1] All references to Kosovo in this document should be understood to be in the context of the United Nations Security Council resolution 1244 (1999).

*Supplementary Figure 1: Number of observed deaths and lives saved in population aged 25 and older, in context of circulating VOC and progress in vaccination, in 34 countries, areas and territories (CAT) in the European Region, from week 50/2020 to 12/2023.*

*Supplementary table 1: Summary table of each countries, areas and territories (CAT) in the WHO European Region with data availability and whether they were included in the analysis. Finer older refers to the presence of data relating to finer age older age groups, namely 60-69 years, 70-79 years and ≥80 years.*

| | | Data type | | | |
|---|---|---|---|---|---|
| **Countries, areas and territories** | **Age group** | **Mortality** | **Vaccination** | **Variant** | **Included in full or partial analysis?** |
| Albania | 25-49 | No | No | Yes[1] | Partial |
| | 50-59 | No | No | Yes[1] | Partial |
| | ≥60 | No | No | Yes[1] | Partial |
| | Finer older | No | No | Yes[1] | Partial |
| Armenia | 25-49 | No | Yes | Yes | No |
| | 50-59 | No | Yes | Yes | No |
| | ≥60 | No | Yes | Yes | No |
| | Finer older | No | Yes | Yes | No |
| Austria | 25-49 | Yes | Yes | Yes | Full |
| | 50-59 | Yes | Yes | Yes | Full |
| | ≥60 | Yes | Yes | Yes | Full |
| | Finer older | Yes | Yes | Yes | Full |
| Azerbaijan | 25-49 | No | No | No | No |
| | 50-59 | No | No | No | No |
| | ≥60 | No | No | No | No |
| | Finer older | No | No | No | No |
| Belarus | 25-49 | Yes | No | No | No |
| | 50-59 | Yes | No | No | No |
| | ≥60 | Yes | No | No | No |
| | Finer older | Yes | No | No | No |
| Belgium | 25-49 | Yes | Yes | Yes | Full |
| | 50-59 | Yes | Yes | Yes | Full |

| | | Data type | | | |
|---|---|---|---|---|---|
| **Countries, areas and territories** | **Age group** | **Mortality** | **Vaccination** | **Variant** | **Included in full or partial analysis?** |
| | ≥60 | Yes | Yes | Yes | Full |
| | Finer older | Yes | Yes | Yes | Full |
| Bosnia and Herzegovina[3] | 25-49 | Yes | No | Yes | No |
| | 50-59 | Yes | No | Yes | No |
| | ≥60 | Yes | Yes | Yes | No |
| | Finer older | Yes | No | Yes | No |
| Bulgaria | 25-49 | No | Yes | Yes | No |
| | 50-59 | No | Yes | Yes | No |
| | ≥60 | No | Yes | Yes | No |
| | Finer older | No | Yes | Yes | No |
| Croatia | 25-49 | Yes[6] | Yes | Yes | Full |
| | 50-59 | Yes[6] | Yes | Yes | Full |
| | ≥60 | Yes[6] | Yes | Yes | Full |
| | Finer older | Yes[6] | Yes | Yes | Full |
| Cyprus | 25-49 | Yes | Yes | Yes | Full |
| | 50-59 | Yes | Yes | Yes | Full |
| | ≥60 | Yes | Yes | Yes | Full |
| | Finer older | Yes | Yes | Yes | Full |
| Czechia | 25-49 | Yes | Yes | Yes | Full |
| | 50-59 | Yes | Yes | Yes | Full |
| | ≥60 | Yes | Yes | Yes | Full |
| | Finer older | Yes | Yes | Yes | Full |
| Denmark | 25-49 | Yes | Yes | Yes | Full |
| | 50-59 | Yes | Yes | Yes | Full |

| | | Data type | | | |
|---|---|---|---|---|---|
| **Countries, areas and territories** | **Age group** | **Mortality** | **Vaccination** | **Variant** | **Included in full or partial analysis?** |
| | ≥60 | Yes | Yes | Yes | Full |
| | Finer older | Yes | Yes | Yes | Full |
| Estonia | 25-49 | Yes | Yes | Yes | Full |
| | 50-59 | Yes | Yes | Yes | Full |
| | ≥60 | Yes | Yes | Yes | Full |
| | Finer older | Yes | Yes | Yes | Full |
| Finland | 25-49 | Yes | Yes | Yes | Full |
| | 50-59 | Yes | Yes | Yes | Full |
| | ≥60 | Yes | Yes | Yes | Full |
| | Finer older | Yes | Yes | Yes | Full |
| France | 25-49 | Yes | Yes | Yes | Full |
| | 50-59 | Yes | Yes | Yes | Full |
| | ≥60 | Yes | Yes | Yes | Full |
| | Finer older | Yes | Yes | Yes | Full |
| Georgia | 25-49 | No | No | No | No |
| | 50-59 | No | No | No | No |
| | ≥60 | No | Yes | No | No |
| | Finer older | No | No | No | No |
| Germany | 25-49 | Yes | No | Yes | No |
| | 50-59 | Yes | No | Yes | No |
| | ≥60 | Yes | Yes | Yes | Full |
| | Finer older | Yes | No | Yes | No |
| Greece | 25-49 | Yes | Yes | Yes | Full |
| | 50-59 | Yes | Yes | Yes | Full |

| Countries, areas and territories | Age group | Data type: Mortality | Data type: Vaccination | Data type: Variant | Included in full or partial analysis? |
|---|---|---|---|---|---|
| | ≥60 | Yes | Yes | Yes | Full |
| | Finer older | Yes | Yes | Yes | Full |
| Hungary | 25-49 | Yes | Yes | Yes | Full |
| | 50-59 | Yes | Yes | Yes | Full |
| | ≥60 | Yes | Yes | Yes | Full |
| | Finer older | Yes | Yes | Yes | Full |
| Iceland | 25-49 | Yes[6] | Yes | Yes | Full |
| | 50-59 | Yes[6] | Yes | Yes | Full |
| | ≥60 | Yes[6] | Yes | Yes | Full |
| | Finer older | Yes | Yes | Yes | No |
| Ireland | 25-49 | Yes | Yes | Yes | Full |
| | 50-59 | Yes | Yes | Yes | Full |
| | ≥60 | Yes | Yes | Yes | Full |
| | Finer older | Yes | Yes | Yes | Full |
| Israel | 25-49 | Yes[6] | Yes[6] | Yes[2] | Full |
| | 50-59 | Yes[6] | Yes[6] | Yes[2] | Full |
| | ≥60 | Yes[6] | Yes[6] | Yes[2] | Full |
| | Finer older | No[6] | No[6] | Yes[2] | No |
| Italy | 25-49 | Yes | Yes | Yes | Full |
| | 50-59 | Yes | Yes | Yes | Full |
| | ≥60 | Yes | Yes | Yes | Full |
| | Finer older | Yes | Yes | Yes | Full |
| Kazakhstan | 25-49 | No | No | Yes | No |
| | 50-59 | No | No | Yes | No |

| | | Data type | | | |
|---|---|---|---|---|---|
| Countries, areas and territories | Age group | Mortality | Vaccination | Variant | Included in full or partial analysis? |
| | ≥60 | No | No | Yes | No |
| | Finer older | No | No | Yes | No |
| Kyrgyzstan | 25-49 | Yes | No | No | No |
| | 50-59 | Yes | No | No | No |
| | ≥60 | Yes | Yes | No | No |
| | Finer older | Yes | No | No | No |
| Latvia | 25-49 | Yes | Yes | Yes | Full |
| | 50-59 | Yes | Yes | Yes | Full |
| | ≥60 | Yes | Yes | Yes | Full |
| | Finer older | Yes | Yes | Yes | Full |
| Lithuania | 25-49 | Yes[6] | Yes | Yes | Full |
| | 50-59 | Yes[6] | Yes | Yes | Full |
| | ≥60 | Yes[6] | Yes | Yes | Full |
| | Finer older | Yes[6] | Yes | Yes | Full |
| Luxembourg | 25-49 | Yes | Yes | Yes | Full |
| | 50-59 | Yes | Yes | Yes | Full |
| | ≥60 | Yes | Yes | Yes | Full |
| | Finer older | Yes | Yes | Yes | Full |
| Malta | 25-49 | Yes | Yes | Yes | Full |
| | 50-59 | Yes | Yes | Yes | Full |
| | ≥60 | Yes | Yes | Yes | Full |
| | Finer older | Yes | Yes | Yes | Full |
| Montenegro | 25-49 | No | No | Yes | Partial |
| | 50-59 | No | No | Yes | Partial |

| | | Data type | | | |
|---|---|---|---|---|---|
| Countries, areas and territories | Age group | Mortality | Vaccination | Variant | Included in full or partial analysis? |
| | ≥60 | No | No | Yes | Partial |
| | Finer older | No | No | Yes | Partial |
| Netherlands (Kingdom of the) | 25-49 | Yes | Yes[6] | Yes | Full |
| | 50-59 | Yes | Yes[6] | Yes | Full |
| | ≥60 | Yes | Yes[6] | Yes | Full |
| | Finer older | Yes | Yes[6] | Yes | Full |
| North Macedonia | 25-49 | Yes | Yes | Yes | Full |
| | 50-59 | Yes | Yes | Yes | Full |
| | ≥60 | Yes | Yes | Yes | Full |
| | Finer older | Yes | Yes | Yes | Full |
| Norway[4] | 25-49 | No | No | Yes | Partial |
| | 50-59 | No | No | Yes | Partial |
| | ≥60 | No | No | Yes | Partial |
| | Finer older | No | No | Yes | Partial |
| Poland | 25-49 | No | No | Yes | Partial |
| | 50-59 | No | No | Yes | Partial |
| | ≥60 | No | No | Yes | Partial |
| | Finer older | No | No | Yes | Partial |
| Portugal | 25-49 | Yes | Yes | Yes | Full |
| | 50-59 | Yes | Yes | Yes | Full |
| | ≥60 | Yes | Yes | Yes | Full |
| | Finer older | Yes | Yes | Yes | Full |
| Republic of Moldova | 25-49 | Yes | No | Yes[1] | No |
| | 50-59 | Yes | No | Yes[1] | No |

| | | Data type | | | |
|---|---|---|---|---|---|
| Countries, areas and territories | Age group | Mortality | Vaccination | Variant | Included in full or partial analysis? |
| | ≥60 | Yes | Yes | Yes[1] | Full |
| | Finer older | Yes | No | Yes[1] | No |
| Romania | 25-49 | Yes[6] | Yes | Yes | Full |
| | 50-59 | Yes[6] | Yes | Yes | Full |
| | ≥60 | Yes[6] | Yes | Yes | Full |
| | Finer older | Yes[6] | Yes | Yes | Full |
| Russian Federation | 25-49 | No | No | No | No |
| | 50-59 | No | No | No | No |
| | ≥60 | No | No | No | No |
| | Finer older | No | No | No | No |
| Serbia | 25-49 | No | Yes | No | No |
| | 50-59 | No | Yes | No | No |
| | ≥60 | No | Yes | No | No |
| | Finer older | No | Yes | No | No |
| Slovakia | 25-49 | Yes | Yes | Yes | Full |
| | 50-59 | Yes | Yes | Yes | Full |
| | ≥60 | Yes | Yes | Yes | Full |
| | Finer older | Yes | Yes | Yes | Full |
| Slovenia | 25-49 | Yes | Yes | Yes | Full |
| | 50-59 | Yes | Yes | Yes | Full |
| | ≥60 | Yes | Yes | Yes | Full |
| | Finer older | Yes | Yes | Yes | Full |
| Spain | 25-49 | Yes | Yes | Yes | Full |
| | 50-59 | Yes | Yes | Yes | Full |

| | | Data type | | | |
|---|---|---|---|---|---|
| **Countries, areas and territories** | **Age group** | **Mortality** | **Vaccination** | **Variant** | **Included in full or partial analysis?** |
| | ≥60 | Yes | Yes | Yes | Full |
| | Finer older | Yes | Yes | Yes | Full |
| Sweden | 25-49 | Yes | Yes | Yes | Full |
| | 50-59 | Yes | Yes | Yes | Full |
| | ≥60 | Yes | Yes | Yes | Full |
| | Finer older | Yes | Yes | Yes | Full |
| Switzerland | 25-49 | Yes | Yes | Yes[2] | Full |
| | 50-59 | Yes | Yes | Yes[2] | Full |
| | ≥60 | Yes | Yes | Yes[2] | Full |
| | Finer older | Yes | Yes | Yes[2] | Full |
| Tajikistan | 25-49 | No | No | No | No |
| | 50-59 | No | No | No | No |
| | ≥60 | No | No | No | No |
| | Finer older | No | No | No | No |
| Turkmenistan | 25-49 | No | No | No | No |
| | 50-59 | No | No | No | No |
| | ≥60 | No | No | No | No |
| | Finer older | No | No | No | No |
| Türkiye[5] | 25-49 | Yes | No | Yes | No |
| | 50-59 | Yes | No | Yes | No |
| | ≥60 | Yes | No | Yes | No |
| | Finer older | Yes | No | Yes | No |
| Ukraine | 25-49 | Yes | No | Yes | No |
| | 50-59 | Yes | No | Yes | No |

| | | Data type | | | |
|---|---|---|---|---|---|
| Countries, areas and territories | Age group | Mortality | Vaccination | Variant | Included in full or partial analysis? |
| | ≥60 | Yes | Yes[6] | Yes | Full |
| | Finer older | Yes | No | Yes | No |
| United Kingdom (England) | 25-49 | Yes | Yes | Yes | Full |
| | 50-59 | Yes | Yes | Yes | Full |
| | ≥60 | Yes | Yes | Yes | Full |
| | Finer older | Yes | Yes | Yes | Full |
| United Kingdom (Northern Ireland) | 25-49 | No | No | No | No |
| | 50-59 | No | No | No | No |
| | ≥60 | No | No | No | No |
| | Finer older | No | No | No | No |
| United Kingdom (Scotland) | 25-49 | Yes[6] | Yes[6] | Yes | Full |
| | 50-59 | Yes[6] | Yes[6] | Yes | Full |
| | ≥60 | Yes[6] | Yes[6] | Yes | Full |
| | Finer older | No | Yes | Yes | No |
| United Kingdom (Wales) | 25-49 | No | No | Yes | Partial |
| | 50-59 | No | No | Yes | Partial |
| | ≥60 | No | No | Yes | Partial |
| | Finer older | No | No | Yes | Partial |
| Uzbekistan | 25-49 | No | No | No | No |
| | 50-59 | No | No | No | No |
| | ≥60 | No | Yes | No | No |
| | Finer older | No | No | No | No |
| Kosovo[1] | 25-49 | Yes[6] | Yes | Yes[1] | Full |

| | | Data type | | | |
|---|---|---|---|---|---|
| **Countries, areas and territories** | **Age group** | **Mortality** | **Vaccination** | **Variant** | **Included in full or partial analysis?** |
| | 50-59 | Yes[6] | Yes | Yes[1] | Full |
| | ≥60 | Yes[6] | Yes | Yes[1] | Full |
| | Finer older | Yes[6] | Yes | Yes[1] | Full |

1 Calculated as median start and end weeks of neighboring CAT.
2 Data from GISAID.
3 Although Bosnia and Herzegovina is reporting vaccination data, it is not possible to disaggregate by dose for some weeks.
4 Due to a change in reporting system this country could not be included in the full analysis.
5 Cannot be disaggregated by age.
6 Data was completed or updated after data cut.

[1] All references to Kosovo in this document should be understood to be in the context of the United Nations Security Council resolution 1244 (1999).

*Supplementary table 2: Age group and countries, areas and territories (CAT) breakdown of vaccination uptake (VU), number of deaths observed and averted as well as observed and expected Mortality Rates (MR), up to week 12/2023 in population aged ≥25 years, for the 34 WHO European Region CAT with sufficient data.*

| | | Vaccination Uptake | | | | Deaths | | Mortality Rate | | |
|---|---|---|---|---|---|---|---|---|---|---|
| Countries, areas and territories | Age group (years) | $VU_2$ | $VU_3$ | $VU_4$ | $VU_5$ | Reported | Averted | Reported | Expected | % change |
| Austria | 25-49 | 72 | 61 | 12 | 0 | 272 | 197 | 9 | 16 | 42 |
| | 50-59 | 80 | 74 | 22 | 1 | 687 | 623 | 49 | 94 | 48 |
| | 60-69 | 84 | 79 | 37 | 1 | 1,837 | 1,829 | 169 | 337 | 50 |
| | 70-79 | 89 | 85 | 49 | 2 | 3,882 | 4,219 | 523 | 1,092 | 52 |
| | 80+ | 95 | 86 | 51 | 2 | 10,540 | 16,046 | 2,011 | 5,072 | 60 |
| | 60+ | 88 | 83 | 44 | 2 | 16,259 | 22,094 | 691 | 1,631 | 58 |
| Belgium | 25-49 | 79 | 66 | 23 | 1 | 98 | 84 | 3 | 5 | 46 |
| | 50-59 | 86 | 82 | 48 | 3 | 516 | 613 | 33 | 71 | 54 |
| | 60-69 | 92 | 89 | 65 | 5 | 1,814 | 2,633 | 130 | 320 | 59 |
| | 70-79 | 95 | 92 | 74 | 8 | 3,749 | 6,217 | 382 | 1,016 | 62 |
| | 80+ | 98 | 94 | 74 | 41 | 9,744 | 16,850 | 1,505 | 4,107 | 63 |
| | 60+ | 94 | 91 | 70 | 14 | 15,307 | 25,700 | 507 | 1,358 | 63 |
| Croatia | 25-49 | 54 | 16 | 0 | 0 | 81 | 19 | 7 | 8 | 19 |
| | 50-59 | 68 | 30 | 1 | 0 | 203 | 144 | 38 | 65 | 41 |
| | 60-69 | 76 | 47 | 3 | 0 | 716 | 604 | 126 | 233 | 46 |
| | 70-79 | 78 | 62 | 7 | 2 | 1,369 | 1,243 | 364 | 694 | 48 |
| | 80+ | 66 | 53 | 7 | 2 | 2,436 | 1,606 | 1,151 | 1,910 | 40 |
| | 60+ | 75 | 53 | 5 | 1 | 4,521 | 3,453 | 391 | 690 | 43 |
| Cyprus | 25-49 | 78 | 59 | 2 | 0 | 46 | 29 | 13 | 22 | 39 |
| | 50-59 | 82 | 74 | 6 | 0 | 75 | 94 | 69 | 155 | 56 |
| | 60-69 | 88 | 82 | 20 | 3 | 211 | 256 | 218 | 483 | 55 |
| | 70-79 | 93 | 90 | 38 | 10 | 387 | 749 | 561 | 1,647 | 66 |

| | | Vaccination Uptake | | | | Deaths | | Mortality Rate | | |
|---|---|---|---|---|---|---|---|---|---|---|
| Countries, areas and territories | Age group (years) | $VU_2$ | $VU_3$ | $VU_4$ | $VU_5$ | Reported | Averted | Reported | Expected | % change |
| | 80+ | 91 | 84 | 38 | 11 | 771 | 1,530 | 2,166 | 6,463 | 66 |
| | 60+ | 90 | 85 | 29 | 7 | 1,369 | 2,535 | 680 | 1,939 | 65 |
| Czechia | 25-49 | 64 | 37 | 2 | 0 | 623 | 220 | 17 | 23 | 26 |
| | 50-59 | 72 | 54 | 6 | 0 | 1,398 | 640 | 102 | 149 | 31 |
| | 60-69 | 79 | 69 | 16 | 0 | 4,888 | 2,339 | 388 | 574 | 32 |
| | 70-79 | 86 | 77 | 27 | 0 | 11,788 | 7,934 | 1,114 | 1,864 | 40 |
| | 80+ | 87 | 75 | 30 | 0 | 14,595 | 13,530 | 3,235 | 6,234 | 48 |
| | 60+ | 83 | 73 | 23 | 0 | 31,271 | 23,803 | 1,129 | 1,989 | 43 |
| Denmark | 25-49 | 84 | 63 | 5 | 0 | 130 | 154 | 7 | 15 | 54 |
| | 50-59 | 93 | 87 | 58 | 1 | 217 | 409 | 27 | 78 | 65 |
| | 60-69 | 97 | 94 | 79 | 1 | 656 | 1,437 | 97 | 311 | 69 |
| | 70-79 | 98 | 97 | 89 | 3 | 1,823 | 3,982 | 314 | 999 | 69 |
| | 80+ | 100 | 100 | 96 | 3 | 4,293 | 9,218 | 1,473 | 4,635 | 68 |
| | 60+ | 99 | 97 | 86 | 2 | 6,772 | 14,637 | 540 | 1,706 | 68 |
| Estonia | 25-49 | 57 | 34 | 5 | 0 | 59 | 18 | 13 | 17 | 23 |
| | 50-59 | 67 | 46 | 8 | 0 | 107 | 52 | 61 | 91 | 33 |
| | 60-69 | 72 | 54 | 15 | 1 | 330 | 191 | 196 | 310 | 37 |
| | 70-79 | 80 | 65 | 22 | 1 | 633 | 555 | 546 | 1,026 | 47 |
| | 80+ | 82 | 64 | 23 | 1 | 1,504 | 1,670 | 1,876 | 3,958 | 53 |
| | 60+ | 77 | 60 | 19 | 1 | 2,467 | 2,416 | 677 | 1,341 | 49 |
| Finland | 25-49 | 83 | 52 | 3 | 0 | 157 | 201 | 9 | 20 | 56 |
| | 50-59 | 92 | 77 | 11 | 2 | 226 | 382 | 32 | 86 | 63 |
| | 60-69 | 94 | 87 | 54 | 15 | 647 | 1,327 | 91 | 278 | 67 |
| | 70-79 | 96 | 92 | 79 | 41 | 1,895 | 4,287 | 319 | 1,040 | 69 |

| | | Vaccination Uptake | | | | Deaths | | Mortality Rate | | |
|---|---|---|---|---|---|---|---|---|---|---|
| Countries, areas and territories | Age group (years) | $VU_2$ | $VU_3$ | $VU_4$ | $VU_5$ | Reported | Averted | Reported | Expected | % change |
| | 80+ | 92 | 91 | 81 | 49 | 5,583 | 11,342 | 1,688 | 5,118 | 67 |
| | 60+ | 94 | 89 | 68 | 32 | 8,125 | 16,956 | 497 | 1,535 | 68 |
| France | 25-49 | 89 | 68 | 4 | 0 | 1,730 | 2,106 | 8 | 19 | 55 |
| | 50-59 | 91 | 80 | 13 | 1 | 3,998 | 4,935 | 45 | 100 | 55 |
| | 60-69 | 91 | 84 | 38 | 8 | 11,431 | 13,424 | 141 | 307 | 54 |
| | 70-79 | 90 | 85 | 49 | 13 | 22,785 | 27,648 | 366 | 809 | 55 |
| | 80+ | 88 | 80 | 49 | 16 | 58,884 | 63,554 | 1,441 | 2,997 | 52 |
| | 60+ | 90 | 83 | 44 | 11 | 93,100 | 104,626 | 506 | 1,074 | 53 |
| Germany | 60+ | 90 | 85 | 39 | 8 | 135,420 | 154,457 | 554 | 1,187 | 53 |
| Greece | 25-49 | 70 | 60 | 2 | 0 | 843 | 669 | 25 | 45 | 44 |
| | 50-59 | 77 | 70 | 7 | 1 | 1,939 | 1,725 | 126 | 237 | 47 |
| | 60-69 | 86 | 78 | 21 | 4 | 4,613 | 5,302 | 349 | 750 | 53 |
| | 70-79 | 91 | 84 | 33 | 8 | 7,969 | 10,636 | 802 | 1,872 | 57 |
| | 80+ | 87 | 77 | 30 | 7 | 18,081 | 25,629 | 2,398 | 5,796 | 59 |
| | 60+ | 88 | 80 | 27 | 6 | 30,663 | 41,567 | 999 | 2,353 | 58 |
| Hungary | 25-49 | 63 | 36 | 1 | 0 | 1,798 | 845 | 52 | 77 | 32 |
| | 50-59 | 70 | 50 | 3 | 0 | 3,423 | 1,930 | 270 | 422 | 36 |
| | 60-69 | 81 | 65 | 9 | 0 | 8,408 | 6,352 | 678 | 1,190 | 43 |
| | 70-79 | 84 | 74 | 17 | 1 | 12,262 | 11,039 | 1,385 | 2,632 | 47 |
| | 80+ | 75 | 64 | 15 | 1 | 15,337 | 10,857 | 3,444 | 5,882 | 41 |
| | 60+ | 81 | 68 | 13 | 0 | 36,007 | 28,248 | 1,401 | 2,500 | 44 |

| Countries, areas and territories | Age group (years) | Vaccination Uptake | | | | Deaths | | Mortality Rate | | |
|---|---|---|---|---|---|---|---|---|---|---|
| | | $VU_2$ | $VU_3$ | $VU_4$ | $VU_5$ | Reported | Averted | Reported | Expected | % change |
| Iceland | 25-49 | 57 | 81 | 4 | 0 | 1 | 3 | 1 | 3 | 75 |
| | 50-59 | 84 | 90 | 13 | 1 | 6 | 10 | 14 | 36 | 62 |
| | 60-69 | 93 | 91 | 58 | 13 | 13 | 25 | 33 | 98 | 66 |
| | 70-79 | 97 | 94 | 76 | 28 | 56 | 134 | 221 | 751 | 71 |
| | 80+ | 100 | 94 | 84 | 32 | 159 | 340 | 1,215 | 3,812 | 68 |
| | 60+ | 96 | 93 | 68 | 21 | 228 | 499 | 295 | 940 | 69 |
| Ireland | 25-49 | 85 | 65 | 12 | 0 | 143 | 153 | 8 | 17 | 52 |
| | 50-59 | 87 | 87 | 39 | 2 | 239 | 446 | 38 | 109 | 65 |
| | 60-69 | 95 | 91 | 60 | 19 | 638 | 1,070 | 127 | 339 | 63 |
| | 70-79 | 100 | 100 | 87 | 55 | 1,448 | 2,570 | 418 | 1,161 | 64 |
| | 80+ | 100 | 100 | 95 | 60 | 3,534 | 4,319 | 1,943 | 4,317 | 55 |
| | 60+ | 100 | 100 | 76 | 38 | 5,620 | 7,959 | 545 | 1,317 | 59 |
| Israel | 25-49 | 89 | 66 | 4 | 0 | 270 | 386 | 10 | 24 | 59 |
| | 50-59 | 95 | 81 | 10 | 0 | 458 | 1,004 | 58 | 184 | 69 |
| | 60+ | 100 | 100 | 57 | 1 | 8,669 | 24,857 | 667 | 2,579 | 74 |
| Italy | 25-49 | 85 | 81 | 3 | 0 | 1,801 | 1,692 | 10 | 19 | 48 |
| | 50-59 | 85 | 87 | 6 | 0 | 4,795 | 4,464 | 50 | 97 | 48 |
| | 60-69 | 85 | 91 | 21 | 1 | 12,880 | 12,023 | 170 | 329 | 48 |
| | 70-79 | 90 | 93 | 33 | 2 | 30,768 | 32,432 | 511 | 1,050 | 51 |
| | 80+ | 96 | 93 | 46 | 6 | 81,339 | 121,265 | 1,805 | 4,497 | 60 |
| | 60+ | 90 | 92 | 31 | 3 | 124,987 | 165,720 | 690 | 1,606 | 57 |
| Latvia | 25-49 | 61 | 32 | 2 | 0 | 222 | 166 | 36 | 63 | 43 |
| | 50-59 | 62 | 36 | 3 | 0 | 415 | 294 | 161 | 275 | 41 |
| | 60-69 | 65 | 42 | 6 | 1 | 936 | 709 | 379 | 666 | 43 |

| | | Vaccination Uptake | | | | Deaths | | Mortality Rate | | |
|---|---|---|---|---|---|---|---|---|---|---|
| Countries, areas and territories | Age group (years) | $VU_2$ | $VU_3$ | $VU_4$ | $VU_5$ | Reported | Averted | Reported | Expected | % change |
| | 70-79 | 67 | 48 | 11 | 3 | 1,481 | 1,094 | 892 | 1,551 | 42 |
| | 80+ | 54 | 39 | 10 | 3 | 2,605 | 1,358 | 2,298 | 3,496 | 34 |
| | 60+ | 63 | 43 | 9 | 2 | 5,022 | 3,161 | 954 | 1,554 | 39 |
| Lithuania | 25-49 | 63 | 32 | 1 | 0 | 291 | 269 | 32 | 61 | 48 |
| | 50-59 | 67 | 37 | 1 | 0 | 512 | 429 | 122 | 225 | 46 |
| | 60-69 | 72 | 47 | 2 | 0 | 1,155 | 1,069 | 304 | 586 | 48 |
| | 70-79 | 75 | 57 | 4 | 0 | 2,167 | 2,057 | 900 | 1,754 | 49 |
| | 80+ | 64 | 49 | 5 | 0 | 4,070 | 2,465 | 2,575 | 4,135 | 38 |
| | 60+ | 71 | 51 | 3 | 0 | 7,392 | 5,591 | 949 | 1,667 | 43 |
| Luxembourg | 25-49 | 68 | 59 | 3 | 0 | 13 | 0 | 5 | 5 | 0 |
| | 50-59 | 74 | 74 | 7 | 0 | 35 | 31 | 38 | 72 | 47 |
| | 60-69 | 80 | 80 | 39 | 1 | 85 | 79 | 129 | 248 | 48 |
| | 70-79 | 87 | 84 | 53 | 0 | 148 | 145 | 365 | 723 | 49 |
| | 80+ | 97 | 90 | 63 | 1 | 473 | 809 | 1,862 | 5,046 | 63 |
| | 60+ | 86 | 83 | 48 | 1 | 706 | 1,033 | 535 | 1,318 | 59 |
| Malta | 25-49 | 82 | 78 | 1 | 0 | 14 | 30 | 7 | 21 | 68 |
| | 50-59 | 86 | 89 | 4 | 0 | 10 | 18 | 17 | 48 | 64 |
| | 60-69 | 93 | 95 | 26 | 3 | 68 | 153 | 114 | 370 | 69 |
| | 70-79 | 98 | 80 | 50 | 6 | 138 | 311 | 289 | 941 | 69 |
| | 80+ | 100 | 83 | 82 | 9 | 274 | 635 | 1,276 | 4,234 | 70 |
| | 60+ | 97 | 87 | 44 | 5 | 480 | 1,099 | 372 | 1,225 | 70 |
| Netherlands | 25-49 | 63 | 50 | 9 | 0 | 137 | 73 | 2 | 4 | 35 |
| | 50-59 | 70 | 70 | 23 | 1 | 352 | 273 | 14 | 25 | 44 |
| | 60-69 | 89 | 87 | 57 | 29 | 1,091 | 1,281 | 50 | 109 | 54 |

| | | Vaccination Uptake | | | | Deaths | | Mortality Rate | | |
|---|---|---|---|---|---|---|---|---|---|---|
| **Countries, areas and territories** | **Age group (years)** | **$VU_2$** | **$VU_3$** | **$VU_4$** | **$VU_5$** | **Reported** | **Averted** | **Reported** | **Expected** | **% change** |
| | 70-79 | 97 | 96 | 80 | 59 | 3,150 | 4,211 | 190 | 445 | 57 |
| | 80+ | 100 | 100 | 86 | 65 | 8,260 | 11,417 | 968 | 2,307 | 58 |
| | 60+ | 94 | 93 | 70 | 46 | 12,501 | 16,909 | 267 | 628 | 57 |
| North Macedonia | 25-49 | 51 | 5 | 0 | 0 | 396 | 92 | 64 | 79 | 19 |
| | 50-59 | 61 | 10 | 0 | 0 | 714 | 219 | 282 | 368 | 23 |
| | 60-69 | 68 | 23 | 1 | 0 | 1,947 | 831 | 815 | 1,162 | 30 |
| | 70-79 | 69 | 29 | 3 | 0 | 2,647 | 1,373 | 1,825 | 2,771 | 34 |
| | 80+ | 51 | 18 | 2 | 0 | 1,888 | 706 | 3,366 | 4,625 | 27 |
| | 60+ | 66 | 24 | 2 | 0 | 6,482 | 2,910 | 2,712 | 3,930 | 31 |
| Portugal | 25-49 | 76 | 70 | 7 | 0 | 297 | 355 | 9 | 20 | 54 |
| | 50-59 | 78 | 88 | 43 | 0 | 611 | 802 | 41 | 94 | 57 |
| | 60-69 | 87 | 92 | 61 | 1 | 1,826 | 2,139 | 132 | 286 | 54 |
| | 70-79 | 90 | 92 | 74 | 1 | 4,467 | 5,304 | 407 | 890 | 54 |
| | 80+ | 90 | 93 | 82 | 51 | 14,146 | 18,444 | 1,953 | 4,500 | 57 |
| | 60+ | 89 | 92 | 70 | 12 | 20,439 | 25,887 | 638 | 1,445 | 56 |
| Republic of Moldova | 60+ | 39 | 22 | 2 | 0 | 5,983 | 1,671 | 844 | 1,080 | 22 |
| Romania | 25-49 | 37 | 10 | 0 | 0 | 2,283 | 618 | 36 | 46 | 21 |
| | 50-59 | 40 | 13 | 0 | 0 | 4,344 | 1,350 | 162 | 213 | 24 |
| | 60-69 | 45 | 15 | 0 | 0 | 12,296 | 3,923 | 496 | 655 | 24 |
| | 70-79 | 41 | 14 | 0 | 0 | 17,028 | 4,638 | 1,065 | 1,355 | 21 |
| | 80+ | 26 | 9 | 0 | 0 | 15,193 | 2,099 | 1,794 | 2,042 | 12 |
| | 60+ | 40 | 14 | 0 | 0 | 44,517 | 10,660 | 904 | 1,121 | 19 |

| | | Vaccination Uptake | | | | Deaths | | Mortality Rate | | |
|---|---|---|---|---|---|---|---|---|---|---|
| Countries, areas and territories | Age group (years) | $VU_2$ | $VU_3$ | $VU_4$ | $VU_5$ | Reported | Averted | Reported | Expected | % change |
| Slovakia | 25-49 | 50 | 27 | 1 | 0 | 659 | 172 | 32 | 41 | 21 |
| | 50-59 | 59 | 38 | 2 | 0 | 1,365 | 469 | 194 | 261 | 26 |
| | 60-69 | 69 | 59 | 3 | 0 | 4,073 | 1,672 | 593 | 837 | 29 |
| | 70-79 | 71 | 64 | 5 | 0 | 5,787 | 2,779 | 1,368 | 2,026 | 32 |
| | 80+ | 59 | 53 | 5 | 0 | 5,884 | 2,082 | 3,204 | 4,337 | 26 |
| | 60+ | 68 | 59 | 4 | 0 | 15,744 | 6,533 | 1,218 | 1,723 | 29 |
| Slovenia | 25-49 | 47 | 23 | 1 | 0 | 96 | 26 | 14 | 17 | 21 |
| | 50-59 | 63 | 40 | 2 | 0 | 243 | 153 | 81 | 132 | 39 |
| | 60-69 | 72 | 54 | 7 | 1 | 830 | 658 | 294 | 528 | 44 |
| | 70-79 | 78 | 64 | 13 | 4 | 1,582 | 1,426 | 838 | 1,593 | 47 |
| | 80+ | 69 | 56 | 16 | 4 | 4,501 | 2,990 | 3,829 | 6,373 | 40 |
| | 60+ | 73 | 58 | 11 | 3 | 6,913 | 5,074 | 1,175 | 2,038 | 42 |
| Spain | 25-49 | 75 | 52 | 5 | 0 | 1,160 | 1,056 | 7 | 14 | 48 |
| | 50-59 | 79 | 76 | 13 | 0 | 2,771 | 3,406 | 38 | 85 | 55 |
| | 60-69 | 86 | 89 | 47 | 0 | 7,324 | 9,579 | 130 | 299 | 57 |
| | 70-79 | 93 | 92 | 67 | 0 | 14,632 | 23,076 | 358 | 922 | 61 |
| | 80+ | 100 | 93 | 76 | 0 | 44,371 | 87,081 | 1,539 | 4,559 | 66 |
| | 60+ | 92 | 91 | 60 | 0 | 66,327 | 119,736 | 526 | 1,474 | 64 |
| Sweden | 25-49 | 81 | 53 | 12 | 0 | 208 | 117 | 6 | 9 | 36 |
| | 50-59 | 89 | 75 | 31 | 1 | 392 | 333 | 29 | 54 | 46 |
| | 60-69 | 91 | 85 | 63 | 3 | 1,123 | 1,234 | 100 | 211 | 52 |
| | 70-79 | 93 | 90 | 82 | 4 | 3,783 | 5,082 | 370 | 867 | 57 |
| | 80+ | 83 | 82 | 77 | 2 | 10,603 | 10,446 | 1,821 | 3,615 | 50 |
| | 60+ | 90 | 86 | 73 | 3 | 15,509 | 16,762 | 570 | 1,185 | 52 |

| | | Vaccination Uptake | | | | Deaths | | Mortality Rate | | |
|---|---|---|---|---|---|---|---|---|---|---|
| **Countries, areas and territories** | **Age group (years)** | **$VU_2$** | **$VU_3$** | **$VU_4$** | **$VU_5$** | **Reported** | **Averted** | **Reported** | **Expected** | **% change** |
| Switzerland | 25-49 | 69 | 47 | 2 | 0 | 91 | 46 | 3 | 5 | 34 |
| | 50-59 | 76 | 67 | 6 | 0 | 265 | 184 | 20 | 35 | 41 |
| | 60-69 | 81 | 90 | 13 | 1 | 707 | 550 | 71 | 127 | 44 |
| | 70-79 | 88 | 100 | 29 | 7 | 1,744 | 1,627 | 234 | 453 | 48 |
| | 80+ | 90 | 100 | 38 | 10 | 5,874 | 5,847 | 1,245 | 2,483 | 50 |
| | 60+ | 85 | 100 | 24 | 5 | 8,325 | 8,024 | 377 | 741 | 49 |
| Ukraine | 60+ | 38 | 55 | 27 | 0 | 82,115 | 13,670 | 897 | 1,046 | 14 |
| United Kingdom (England) | 25-49 | 83 | 61 | 6 | 0 | 5,239 | 5,573 | 28 | 58 | 52 |
| | 50-59 | 93 | 82 | 51 | 0 | 9,011 | 13,287 | 117 | 289 | 60 |
| | 60-69 | 100 | 94 | 74 | 0 | 18,739 | 36,958 | 309 | 919 | 66 |
| | 70-79 | 99 | 97 | 34 | 86 | 39,577 | 81,183 | 812 | 2,477 | 67 |
| | 80+ | 100 | 99 | 86 | 88 | 102,234 | 220,989 | 3,630 | 11,476 | 68 |
| | 60+ | 100 | 96 | 62 | 48 | 160,550 | 339,130 | 1,168 | 3,634 | 68 |
| United Kingdom (Scotland) | 25-49 | 82 | 66 | 14 | 1 | 307 | 397 | 17 | 39 | 56 |
| | 50-59 | 95 | 88 | 57 | 3 | 561 | 1,083 | 71 | 208 | 66 |
| | 60+ | 100 | 100 | 91 | 33 | 9,659 | 20,658 | 673 | 2,113 | 68 |
| Kosovo[1] | 25-49 | 59 | 5 | 0 | 0 | 85 | 8 | 14 | 15 | 9 |
| | 50-59 | 69 | 10 | 0 | 0 | 182 | 31 | 93 | 108 | 15 |
| | 60-69 | 74 | 19 | 1 | 0 | 401 | 99 | 287 | 358 | 20 |
| | 70-79 | 75 | 25 | 1 | 0 | 607 | 211 | 807 | 1,087 | 26 |
| | 80+ | 50 | 14 | 1 | 0 | 424 | 83 | 1,209 | 1,445 | 16 |
| | 60+ | 71 | 20 | 1 | 0 | 1,432 | 393 | 573 | 730 | 22 |

| | | Vaccination Uptake | | | | Deaths | | Mortality Rate | | |
|---|---|---|---|---|---|---|---|---|---|---|
| Countries, areas and territories | Age group (years) | $VU_2$ | $VU_3$ | $VU_4$ | $VU_5$ | Reported | Averted | Reported | Expected | % change |
| Total | 25-49 | 76 | 57 | 5 | 0 | 19,550 | 15,774 | 1,095 | 1,979 | 45 |
| | 50-59 | 82 | 73 | 19 | 1 | 40,070 | 39,833 | 5,064 | 10,099 | 50 |
| | 60-69 | 86 | 81 | 38 | 4 | 101,683 | 109,746 | 1,679 | 3,490 | 52 |
| | 70-79 | 89 | 86 | 45 | 19 | 199,752 | 248,162 | 4,097 | 9,188 | 55 |
| | 80+ | 91 | 86 | 57 | 22 | 447,600 | 665,207 | 15,892 | 39,511 | 60 |
| | 60+ | 85 | 82 | 43 | 11 | 1,232,727 | 1,453,741 | 875 | 1,908 | 54 |

Mortality rates are per 100,000 population.

[1] All references to Kosovo in this document should be understood to be in the context of the United Nations Security Council resolution 1244 (1999).

*Supplementary table 3: Age group and countries, areas and territories (CAT) breakdown of vaccination uptake (VU), number of deaths reported and averted as well as reported and expected Mortality Rates (MR) per Variant of Concern, up to week 12/2023 in population aged 25 years and over, for the 34 European Region CAT with sufficient data. Note: the overall total does not include the five CAT that could only be included in the pre-Omicron period, namely: Albania, Montenegro, Norway, Poland and United Kingdom (Wales).*

| | | Alpha | | | | | Delta | | | | | Omicron | | | | |
|---|---|---|---|---|---|---|---|---|---|---|---|---|---|---|---|---|
| | | Total deaths | | Mortality rates | | | Total deaths | | Mortality rates | | | Total deaths | | Mortality rates | | |
| **Countries, areas and territories** | **Age group (years)** | **Reported** | **Averted** | **Reported** | **Expected** | **% change** | **Reported** | **Averted** | **Reported** | **Expected** | **% change** | **Reported** | **Averted** | **Reported** | **Expected** | **% change** |
| Albania | 25-49 | 11 | 0 | 1 | 1 | 0 | 43 | 3 | 4 | 5 | 7 | | | | | |
| | 50-59 | 36 | 0 | 9 | 9 | 0 | 95 | 42 | 25 | 35 | 31 | | | | | |
| | 60+ | 1,029 | 36 | 84 | 87 | 3 | 1,866 | 3,510 | 153 | 441 | 65 | | | | | |
| | 60-69 | 121 | 0 | 37 | 37 | 0 | 209 | 258 | 63 | 141 | 55 | | | | | |
| | 70-79 | 164 | 12 | 84 | 90 | 7 | 263 | 823 | 135 | 558 | 76 | | | | | |
| | 80+ | 58 | 0 | 68 | 68 | 0 | 150 | 89 | 177 | 282 | 37 | | | | | |
| Austria | 25-49 | 72 | 0 | 2 | 2 | 0 | 95 | 102 | 3 | 7 | 52 | 84 | 101 | 3 | 6 | 55 |
| | 50-59 | 219 | 18 | 16 | 17 | 8 | 208 | 315 | 15 | 37 | 60 | 195 | 325 | 14 | 37 | 62 |
| | 60+ | 4,905 | 1,027 | 213 | 258 | 17 | 3,397 | 9,257 | 147 | 549 | 73 | 5,358 | 13,714 | 228 | 811 | 72 |
| | 60-69 | 591 | 46 | 56 | 61 | 7 | 473 | 855 | 45 | 127 | 64 | 546 | 1,049 | 50 | 147 | 66 |
| | 70-79 | 1,240 | 118 | 165 | 180 | 9 | 911 | 1,708 | 121 | 348 | 65 | 1,171 | 2,702 | 158 | 522 | 70 |
| | 80+ | 3,074 | 863 | 612 | 784 | 22 | 2,013 | 6,694 | 401 | 1,734 | 77 | 3,641 | 9,963 | 695 | 2,595 | 73 |

| | | Alpha | | | | | Delta | | | | | Omicron | | | | |
|---|---|---|---|---|---|---|---|---|---|---|---|---|---|---|---|---|
| | | Total deaths | | Mortality rates | | | Total deaths | | Mortality rates | | | Total deaths | | Mortality rates | | |
| Countries, areas and territories | Age group (years) | Reported | Averted | Reported | Expected | % change | Reported | Averted | Reported | Expected | % change | Reported | Averted | Reported | Expected | % change |
| Belgium | 25-49 | 6 | 0 | 0 | 0 | 0 | 31 | 68 | 1 | 3 | 69 | 21 | 26 | 1 | 1 | 55 |
| | 50-59 | 27 | 3 | 2 | 2 | 10 | 125 | 344 | 8 | 29 | 73 | 158 | 313 | 10 | 30 | 66 |
| | 60+ | 369 | 541 | 12 | 31 | 59 | 2,735 | 11,052 | 92 | 465 | 80 | 5,362 | 16,856 | 178 | 736 | 76 |
| | 60-69 | 84 | 32 | 6 | 8 | 28 | 432 | 1,283 | 32 | 125 | 75 | 613 | 1,558 | 44 | 156 | 72 |
| | 70-79 | 120 | 148 | 13 | 28 | 55 | 831 | 2,879 | 88 | 391 | 78 | 1,266 | 3,779 | 129 | 514 | 75 |
| | 80+ | 165 | 361 | 25 | 81 | 69 | 1,472 | 6,890 | 227 | 1,291 | 82 | 3,483 | 11,519 | 538 | 2,317 | 77 |
| Croatia | 25-49 | 14 | 0 | 1 | 1 | 0 | 14 | 5 | 1 | 2 | 26 | 33 | 16 | 3 | 4 | 33 |
| | 50-59 | 20 | 0 | 4 | 4 | 0 | 37 | 39 | 7 | 14 | 51 | 107 | 111 | 20 | 41 | 51 |
| | 60+ | 621 | 127 | 54 | 65 | 17 | 655 | 780 | 57 | 124 | 54 | 2,331 | 2,732 | 202 | 438 | 54 |
| | 60-69 | 113 | 12 | 20 | 22 | 10 | 117 | 166 | 21 | 50 | 59 | 354 | 463 | 62 | 144 | 57 |
| | 70-79 | 197 | 43 | 53 | 65 | 18 | 208 | 319 | 55 | 140 | 61 | 671 | 958 | 178 | 433 | 59 |
| | 80+ | 311 | 72 | 139 | 171 | 19 | 330 | 295 | 156 | 295 | 47 | 1,306 | 1,311 | 617 | 1,237 | 50 |
| Cyprus | 25-49 | 13 | 0 | 4 | 4 | 0 | 20 | 22 | 6 | 13 | 52 | 12 | 7 | 4 | 6 | 37 |
| | 50-59 | 13 | 0 | 12 | 12 | 0 | 26 | 45 | 24 | 65 | 63 | 34 | 55 | 31 | 82 | 62 |
| | 60+ | 209 | 201 | 105 | 207 | 49 | 268 | 894 | 135 | 586 | 77 | 691 | 1,634 | 343 | 1,155 | 70 |

| | | Alpha | | | | | Delta | | | | | Omicron | | | | |
|---|---|---|---|---|---|---|---|---|---|---|---|---|---|---|---|---|
| | | Total deaths | | Mortality rates | | | Total deaths | | Mortality rates | | | Total deaths | | Mortality rates | | |
| Countries, areas and territories | Age group (years) | Reported | Averted | Reported | Expected | % change | Reported | Averted | Reported | Expected | % change | Reported | Averted | Reported | Expected | % change |
| | 60-69 | 56 | 9 | 59 | 68 | 14 | 50 | 117 | 52 | 175 | 70 | 77 | 147 | 80 | 231 | 66 |
| | 70-79 | 62 | 39 | 91 | 149 | 39 | 89 | 323 | 131 | 607 | 78 | 184 | 444 | 267 | 910 | 71 |
| | 80+ | 91 | 153 | 263 | 706 | 63 | 129 | 454 | 373 | 1,687 | 78 | 430 | 1,043 | 1,208 | 4,137 | 71 |
| Czechia | 25-49 | 307 | 4 | 8 | 8 | 1 | 136 | 140 | 4 | 7 | 51 | 102 | 84 | 3 | 5 | 45 |
| | 50-59 | 740 | 17 | 55 | 56 | 2 | 286 | 417 | 21 | 52 | 59 | 214 | 240 | 16 | 33 | 53 |
| | 60+ | 14,540 | 1,868 | 524 | 592 | 11 | 5,526 | 11,763 | 199 | 623 | 68 | 5,826 | 11,822 | 210 | 637 | 67 |
| | 60-69 | 2,669 | 68 | 206 | 212 | 2 | 892 | 1,370 | 69 | 175 | 61 | 644 | 1,050 | 51 | 135 | 62 |
| | 70-79 | 5,995 | 335 | 579 | 611 | 5 | 2,049 | 4,346 | 198 | 617 | 68 | 1,862 | 3,809 | 176 | 536 | 67 |
| | 80+ | 5,876 | 1,465 | 1,321 | 1,650 | 20 | 2,585 | 6,047 | 581 | 1,940 | 70 | 3,320 | 6,963 | 736 | 2,279 | 68 |
| Denmark | 25-49 | 2 | 0 | 0 | 0 | 0 | 24 | 35 | 1 | 3 | 59 | 89 | 128 | 5 | 12 | 59 |
| | 50-59 | 2 | 0 | 0 | 0 | 0 | 31 | 110 | 4 | 18 | 78 | 148 | 333 | 18 | 60 | 69 |
| | 60+ | 17 | 50 | 1 | 4 | 75 | 558 | 1,837 | 37 | 157 | 77 | 4,711 | 14,273 | 375 | 1,513 | 75 |
| | 60-69 | 3 | 0 | 0 | 0 | 0 | 59 | 272 | 9 | 50 | 82 | 466 | 1,323 | 69 | 265 | 74 |
| | 70-79 | 8 | 9 | 1 | 3 | 53 | 148 | 513 | 26 | 115 | 78 | 1,265 | 3,878 | 218 | 885 | 75 |
| | 80+ | 6 | 41 | 2 | 17 | 87 | 351 | 1,052 | 124 | 497 | 75 | 2,980 | 9,072 | 1,022 | 4,135 | 75 |

| | | Alpha | | | | | Delta | | | | | Omicron | | | | |
|---|---|---|---|---|---|---|---|---|---|---|---|---|---|---|---|---|
| | | Total deaths | | Mortality rates | | | Total deaths | | Mortality rates | | | Total deaths | | Mortality rates | | |
| Countries, areas and territories | Age group (years) | Reported | Averted | Reported | Expected | % change | Reported | Averted | Reported | Expected | % change | Reported | Averted | Reported | Expected | % change |
| Estonia | 25-49 | 15 | 0 | 3 | 3 | 0 | 16 | 11 | 3 | 6 | 41 | 24 | 8 | 5 | 7 | 25 |
| | 50-59 | 34 | 0 | 20 | 20 | 0 | 28 | 30 | 16 | 34 | 52 | 37 | 25 | 21 | 35 | 40 |
| | 60+ | 675 | 127 | 189 | 225 | 16 | 495 | 872 | 139 | 383 | 64 | 978 | 1,556 | 268 | 696 | 61 |
| | 60-69 | 113 | 8 | 69 | 73 | 7 | 67 | 91 | 41 | 96 | 58 | 106 | 107 | 63 | 127 | 50 |
| | 70-79 | 212 | 29 | 188 | 214 | 12 | 132 | 223 | 117 | 315 | 63 | 213 | 329 | 184 | 468 | 61 |
| | 80+ | 350 | 90 | 443 | 557 | 20 | 296 | 558 | 375 | 1,081 | 65 | 659 | 1,120 | 822 | 2,219 | 63 |
| Finland | 25-49 | 8 | 0 | 0 | 0 | 0 | 22 | 35 | 1 | 3 | 61 | 122 | 179 | 7 | 17 | 59 |
| | 50-59 | 21 | 0 | 3 | 3 | 0 | 24 | 58 | 3 | 11 | 71 | 173 | 359 | 25 | 76 | 67 |
| | 60+ | 385 | 137 | 24 | 32 | 26 | 473 | 1,557 | 29 | 126 | 77 | 7,118 | 16,874 | 436 | 1,468 | 70 |
| | 60-69 | 44 | 5 | 6 | 7 | 10 | 56 | 183 | 8 | 34 | 77 | 543 | 1,266 | 77 | 255 | 70 |
| | 70-79 | 127 | 39 | 22 | 28 | 23 | 105 | 433 | 18 | 92 | 80 | 1,626 | 4,247 | 274 | 988 | 72 |
| | 80+ | 214 | 93 | 68 | 97 | 30 | 312 | 941 | 99 | 397 | 75 | 4,949 | 11,361 | 1,497 | 4,932 | 70 |
| France | 25-49 | 108 | 13 | 1 | 1 | 11 | 323 | 555 | 2 | 4 | 63 | 816 | 1,705 | 4 | 12 | 68 |
| | 50-59 | 275 | 71 | 3 | 4 | 21 | 630 | 1,381 | 7 | 23 | 69 | 1,744 | 3,871 | 20 | 63 | 69 |
| | 60+ | 3,952 | 4,301 | 22 | 45 | 52 | 8,932 | 21,943 | 49 | 170 | 71 | 40,510 | 86,030 | 220 | 688 | 68 |

| | | Alpha | | | | | Delta | | | | | Omicron | | | | |
|---|---|---|---|---|---|---|---|---|---|---|---|---|---|---|---|---|
| | | Total deaths | | Mortality rates | | | Total deaths | | Mortality rates | | | Total deaths | | Mortality rates | | |
| Countries, areas and territories | Age group (years) | Reported | Averted | Reported | Expected | % change | Reported | Averted | Reported | Expected | % change | Reported | Averted | Reported | Expected | % change |
| | 60-69 | 741 | 330 | 9 | 13 | 31 | 1,435 | 3,429 | 18 | 61 | 70 | 4,851 | 10,641 | 60 | 192 | 69 |
| | 70-79 | 1,152 | 1,234 | 19 | 40 | 52 | 2,478 | 6,985 | 41 | 158 | 74 | 9,652 | 21,425 | 155 | 499 | 69 |
| | 80+ | 2,059 | 2,737 | 50 | 116 | 57 | 5,019 | 11,529 | 121 | 400 | 70 | 26,007 | 53,964 | 637 | 1,957 | 67 |
| Germany | 60+ | 13,745 | 2,260 | 57 | 66 | 14 | 23,456 | 54,798 | 97 | 325 | 70 | 51,332 | 109,368 | 210 | 658 | 68 |
| Greece | 25-49 | 195 | 0 | 6 | 6 | 0 | 308 | 350 | 9 | 19 | 53 | 282 | 352 | 8 | 19 | 56 |
| | 50-59 | 596 | 29 | 39 | 41 | 5 | 582 | 852 | 38 | 94 | 59 | 632 | 932 | 41 | 101 | 60 |
| | 60+ | 6,957 | 2,241 | 225 | 297 | 24 | 6,860 | 11,025 | 222 | 578 | 62 | 15,180 | 31,184 | 495 | 1,510 | 67 |
| | 60-69 | 1,355 | 201 | 104 | 119 | 13 | 1,259 | 2,147 | 97 | 261 | 63 | 1,708 | 3,276 | 129 | 377 | 66 |
| | 70-79 | 2,221 | 402 | 218 | 258 | 15 | 1,875 | 3,440 | 184 | 523 | 65 | 3,402 | 7,543 | 342 | 1,101 | 69 |
| | 80+ | 3,381 | 1,638 | 436 | 647 | 33 | 3,726 | 5,438 | 480 | 1,181 | 59 | 10,070 | 20,365 | 1,335 | 4,036 | 67 |
| Hungary | 25-49 | 10 | 6 | 0 | 0 | 38 | 675 | 725 | 20 | 41 | 52 | 163 | 145 | 5 | 9 | 47 |
| | 50-59 | 10 | 8 | 1 | 1 | 44 | 1,015 | 1,453 | 80 | 195 | 59 | 478 | 558 | 38 | 82 | 54 |
| | 60+ | 64 | 133 | 2 | 8 | 68 | 8,619 | 18,271 | 335 | 1,046 | 68 | 7,720 | 11,736 | 300 | 757 | 60 |

| | | Alpha | | | | | Delta | | | | | Omicron | | | | |
|---|---|---|---|---|---|---|---|---|---|---|---|---|---|---|---|---|
| | | Total deaths | | Mortality rates | | | Total deaths | | Mortality rates | | | Total deaths | | Mortality rates | | |
| Countries, areas and territories | Age group (years) | Reported | Averted | Reported | Expected | % change | Reported | Averted | Reported | Expected | % change | Reported | Averted | Reported | Expected | % change |
| | 60-69 | 14 | 27 | 1 | 3 | 66 | 2,240 | 4,558 | 181 | 548 | 67 | 1,299 | 2,194 | 105 | 282 | 63 |
| | 70-79 | 23 | 60 | 3 | 9 | 72 | 2,917 | 7,620 | 329 | 1,190 | 72 | 2,339 | 4,181 | 264 | 736 | 64 |
| | 80+ | 27 | 46 | 6 | 16 | 63 | 3,462 | 6,093 | 777 | 2,146 | 64 | 4,082 | 5,361 | 917 | 2,120 | 57 |
| Iceland | 25-49 | 0 | 0 | 0 | 0 | - | 0 | 0 | 0 | 0 | - | 1 | 3 | 1 | 3 | 75 |
| | 50-59 | 1 | 0 | 2 | 2 | 0 | 0 | 0 | 0 | 0 | - | 4 | 10 | 9 | 32 | 71 |
| | 60+ | 0 | 0 | 0 | 0 | - | 2 | 13 | 3 | 20 | 87 | 223 | 544 | 288 | 992 | 71 |
| | 60-69 | 0 | 0 | 0 | 0 | - | 0 | 0 | 0 | 0 | - | 12 | 28 | 31 | 103 | 70 |
| | 70-79 | 0 | 0 | 0 | 0 | - | 2 | 13 | 8 | 62 | 87 | 53 | 143 | 210 | 775 | 73 |
| | 80+ | 0 | 0 | 0 | 0 | - | 0 | 0 | 0 | 0 | - | 158 | 373 | 1,207 | 4,057 | 70 |
| Ireland | 25-49 | 49 | 0 | 3 | 3 | 0 | 25 | 58 | 1 | 5 | 70 | 63 | 104 | 4 | 10 | 62 |
| | 50-59 | 92 | 0 | 15 | 15 | 0 | 38 | 208 | 6 | 40 | 85 | 101 | 283 | 16 | 61 | 74 |
| | 60+ | 2,531 | 127 | 252 | 265 | 5 | 594 | 2,343 | 59 | 293 | 80 | 2,311 | 6,237 | 224 | 829 | 73 |
| | 60-69 | 292 | 4 | 59 | 60 | 1 | 89 | 512 | 18 | 122 | 85 | 240 | 654 | 48 | 178 | 73 |
| | 70-79 | 655 | 34 | 196 | 206 | 5 | 173 | 924 | 52 | 328 | 84 | 575 | 1,874 | 166 | 708 | 77 |
| | 80+ | 1,584 | 89 | 900 | 951 | 5 | 332 | 907 | 189 | 704 | 73 | 1,496 | 3,709 | 822 | 2,861 | 71 |

| | | Alpha | | | | | Delta | | | | | Omicron | | | | |
|---|---|---|---|---|---|---|---|---|---|---|---|---|---|---|---|---|
| | | Total deaths | | Mortality rates | | | Total deaths | | Mortality rates | | | Total deaths | | Mortality rates | | |
| Countries, areas and territories | Age group (years) | Reported | Averted | Reported | Expected | % change | Reported | Averted | Reported | Expected | % change | Reported | Averted | Reported | Expected | % change |
| Israel | 25-49 | 99 | 58 | 4 | 6 | 37 | 69 | 218 | 3 | 10 | 76 | 66 | 134 | 2 | 7 | 67 |
| | 50-59 | 149 | 217 | 19 | 46 | 59 | 104 | 501 | 13 | 76 | 83 | 137 | 365 | 17 | 63 | 73 |
| | 60+ | 1,709 | 4,366 | 131 | 467 | 72 | 1,634 | 10,252 | 126 | 914 | 86 | 3,913 | 12,520 | 301 | 1,264 | 76 |
| Italy | 25-49 | 416 | 24 | 2 | 2 | 5 | 222 | 334 | 1 | 3 | 60 | 805 | 1,456 | 4 | 12 | 64 |
| | 50-59 | 1,374 | 121 | 15 | 16 | 8 | 523 | 1,097 | 6 | 17 | 68 | 1,802 | 3,581 | 19 | 56 | 67 |
| | 60+ | 26,877 | 7,194 | 150 | 190 | 21 | 8,461 | 28,102 | 47 | 204 | 77 | 54,404 | 145,853 | 300 | 1,106 | 73 |
| | 60-69 | 3,927 | 364 | 53 | 58 | 8 | 1,037 | 2,691 | 14 | 50 | 72 | 4,582 | 9,905 | 60 | 191 | 68 |
| | 70-79 | 8,406 | 877 | 140 | 155 | 9 | 2,156 | 5,968 | 36 | 136 | 73 | 11,835 | 28,361 | 197 | 668 | 71 |
| | 80+ | 14,544 | 5,953 | 325 | 458 | 29 | 5,268 | 19,443 | 118 | 552 | 79 | 37,987 | 107,587 | 843 | 3,231 | 74 |
| Latvia | 25-49 | 27 | 0 | 4 | 4 | 0 | 91 | 94 | 15 | 30 | 51 | 63 | 82 | 10 | 24 | 57 |
| | 50-59 | 67 | 1 | 26 | 26 | 1 | 154 | 170 | 60 | 126 | 52 | 88 | 139 | 34 | 88 | 61 |
| | 60+ | 631 | 59 | 120 | 131 | 9 | 1,785 | 1,545 | 339 | 633 | 46 | 1,440 | 1,713 | 274 | 599 | 54 |
| | 60-69 | 122 | 5 | 50 | 52 | 4 | 377 | 423 | 153 | 324 | 53 | 204 | 321 | 83 | 212 | 61 |
| | 70-79 | 190 | 25 | 113 | 127 | 12 | 538 | 541 | 324 | 650 | 50 | 400 | 581 | 241 | 591 | 59 |
| | 80+ | 319 | 29 | 281 | 307 | 8 | 870 | 581 | 768 | 1,280 | 40 | 836 | 811 | 738 | 1,453 | 49 |

| | | Alpha | | | | | Delta | | | | | Omicron | | | | |
|---|---|---|---|---|---|---|---|---|---|---|---|---|---|---|---|---|
| | | Total deaths | | Mortality rates | | | Total deaths | | Mortality rates | | | Total deaths | | Mortality rates | | |
| Countries, areas and territories | Age group (years) | Reported | Averted | Reported | Expected | % change | Reported | Averted | Reported | Expected | % change | Reported | Averted | Reported | Expected | % change |
| Lithuania | 25-49 | 11 | 0 | 1 | 1 | 0 | 166 | 240 | 18 | 44 | 59 | 35 | 39 | 4 | 8 | 53 |
| | 50-59 | 30 | 3 | 7 | 8 | 9 | 251 | 394 | 61 | 156 | 61 | 41 | 51 | 10 | 22 | 55 |
| | 60+ | 298 | 147 | 39 | 58 | 33 | 3,132 | 4,000 | 412 | 937 | 56 | 1,482 | 1,687 | 190 | 407 | 53 |
| | 60-69 | 54 | 14 | 15 | 19 | 21 | 495 | 886 | 137 | 384 | 64 | 164 | 215 | 43 | 100 | 57 |
| | 70-79 | 79 | 49 | 33 | 54 | 38 | 938 | 1,555 | 398 | 1,057 | 62 | 389 | 550 | 161 | 390 | 59 |
| | 80+ | 165 | 84 | 100 | 151 | 34 | 1,699 | 1,559 | 1,029 | 1,973 | 48 | 929 | 922 | 588 | 1,171 | 50 |
| Luxembourg | 25-49 | 6 | 0 | 2 | 2 | 0 | 1 | 0 | 0 | 0 | 0 | 5 | 0 | 2 | 2 | 0 |
| | 50-59 | 9 | 0 | 10 | 10 | 0 | 6 | 11 | 7 | 19 | 65 | 19 | 28 | 21 | 51 | 60 |
| | 60+ | 228 | 28 | 178 | 200 | 11 | 82 | 265 | 64 | 271 | 76 | 321 | 843 | 243 | 882 | 72 |
| | 60-69 | 29 | 0 | 46 | 46 | 0 | 14 | 23 | 22 | 58 | 62 | 35 | 66 | 53 | 153 | 65 |
| | 70-79 | 46 | 3 | 117 | 124 | 6 | 17 | 41 | 43 | 147 | 71 | 63 | 116 | 156 | 442 | 65 |
| | 80+ | 153 | 25 | 609 | 708 | 14 | 51 | 201 | 203 | 1,003 | 80 | 223 | 661 | 878 | 3,480 | 75 |
| Malta | 25-49 | 0 | 0 | 0 | 0 | - | 2 | 6 | 1 | 4 | 75 | 12 | 26 | 6 | 18 | 68 |
| | 50-59 | 0 | 0 | 0 | 0 | - | 0 | 0 | 0 | 0 | - | 10 | 19 | 17 | 49 | 66 |
| | 60+ | 19 | 17 | 15 | 28 | 47 | 50 | 255 | 39 | 237 | 84 | 334 | 943 | 259 | 990 | 74 |

| | | Alpha | | | | | Delta | | | | | Omicron | | | | |
|---|---|---|---|---|---|---|---|---|---|---|---|---|---|---|---|---|
| | | Total deaths | | Mortality rates | | | Total deaths | | Mortality rates | | | Total deaths | | Mortality rates | | |
| Countries, areas and territories | Age group (years) | Reported | Averted | Reported | Expected | % change | Reported | Averted | Reported | Expected | % change | Reported | Averted | Reported | Expected | % change |
| | 60-69 | 2 | 0 | 3 | 3 | 0 | 14 | 55 | 23 | 116 | 80 | 43 | 116 | 72 | 266 | 73 |
| | 70-79 | 6 | 0 | 13 | 13 | 0 | 16 | 94 | 34 | 230 | 85 | 93 | 258 | 195 | 735 | 74 |
| | 80+ | 11 | 17 | 51 | 130 | 61 | 20 | 106 | 93 | 587 | 84 | 198 | 569 | 922 | 3,572 | 74 |
| Montenegro | 25-49 | 1 | 0 | 0 | 0 | 0 | 44 | 18 | 20 | 29 | 29 | | | | | |
| Montenegro | 50-59 | 0 | 0 | 0 | 0 | - | 70 | 46 | 87 | 144 | 40 | | | | | |
| Montenegro | 60+ | 78 | 33 | 28 | 40 | 30 | 1,872 | 2,001 | 674 | 1,395 | 52 | | | | | |
| Montenegro | 60-69 | 5 | 1 | 6 | 8 | 17 | 179 | 185 | 233 | 473 | 51 | | | | | |
| Montenegro | 70-79 | 11 | 6 | 27 | 42 | 35 | 237 | 316 | 586 | 1,368 | 57 | | | | | |
| Montenegro | 80+ | 10 | 4 | 47 | 65 | 29 | 208 | 166 | 971 | 1,747 | 44 | | | | | |
| Netherlands | 25-49 | 31 | 0 | 1 | 1 | 0 | 46 | 53 | 1 | 2 | 54 | 29 | 22 | 1 | 1 | 43 |
| Netherlands | 50-59 | 113 | 0 | 4 | 4 | 0 | 73 | 161 | 3 | 9 | 69 | 86 | 127 | 3 | 8 | 60 |
| Netherlands | 60+ | 2,468 | 764 | 54 | 70 | 24 | 3,075 | 12,534 | 67 | 340 | 80 | 1,904 | 5,276 | 41 | 153 | 73 |
| Netherlands | 60-69 | 345 | 51 | 16 | 18 | 13 | 238 | 911 | 11 | 54 | 79 | 203 | 431 | 9 | 29 | 68 |
| Netherlands | 70-79 | 826 | 61 | 51 | 55 | 7 | 666 | 3,324 | 41 | 247 | 83 | 501 | 1,296 | 30 | 109 | 72 |

| | | Alpha | | | | | Delta | | | | | Omicron | | | | |
|---|---|---|---|---|---|---|---|---|---|---|---|---|---|---|---|---|
| | | Total deaths | | Mortality rates | | | Total deaths | | Mortality rates | | | Total deaths | | Mortality rates | | |
| Countries, areas and territories | Age group (years) | Reported | Averted | Reported | Expected | % change | Reported | Averted | Reported | Expected | % change | Reported | Averted | Reported | Expected | % change |
| | 80+ | 1,297 | 652 | 155 | 232 | 33 | 2,171 | 8,299 | 259 | 1,248 | 79 | 1,200 | 3,549 | 141 | 557 | 75 |
| North Macedonia | 25-49 | 158 | 7 | 25 | 27 | 4 | 166 | 73 | 27 | 38 | 31 | 40 | 15 | 6 | 9 | 27 |
| | 50-59 | 309 | 14 | 122 | 127 | 4 | 243 | 168 | 96 | 162 | 41 | 69 | 45 | 27 | 45 | 39 |
| | 60+ | 2,697 | 132 | 613 | 643 | 5 | 1,815 | 1,814 | 412 | 825 | 50 | 1,390 | 1,070 | 582 | 1,029 | 43 |
| | 60-69 | 872 | 39 | 365 | 381 | 4 | 570 | 571 | 239 | 477 | 50 | 297 | 259 | 124 | 233 | 47 |
| | 70-79 | 1,171 | 67 | 807 | 853 | 5 | 721 | 855 | 497 | 1,086 | 54 | 539 | 496 | 372 | 713 | 48 |
| | 80+ | 654 | 26 | 1,166 | 1,212 | 4 | 524 | 388 | 934 | 1,626 | 43 | 554 | 315 | 988 | 1,549 | 36 |
| Norway | 25-49 | 10 | 0 | 1 | 1 | 0 | 11 | 20 | 1 | 2 | 65 | | | | | |
| | 50-59 | 18 | 0 | 3 | 3 | 0 | 23 | 79 | 3 | 14 | 77 | | | | | |
| | 60+ | 672 | 243 | 26 | 36 | 27 | 1,638 | 4,809 | 64 | 253 | 75 | | | | | |
| | 60-69 | 44 | 0 | 7 | 7 | 0 | 55 | 192 | 9 | 42 | 78 | | | | | |
| | 70-79 | 71 | 6 | 16 | 17 | 8 | 153 | 474 | 34 | 139 | 76 | | | | | |
| | 80+ | 109 | 75 | 46 | 78 | 41 | 338 | 937 | 143 | 539 | 73 | | | | | |
| Poland | 25-49 | 209 | 2 | 1 | 2 | 1 | 92 | 72 | 1 | 1 | 44 | | | | | |

| | | Alpha | | | | | Delta | | | | | Omicron | | | | |
|---|---|---|---|---|---|---|---|---|---|---|---|---|---|---|---|---|
| | | Total deaths | | Mortality rates | | | Total deaths | | Mortality rates | | | Total deaths | | Mortality rates | | |
| Countries, areas and territories | Age group (years) | Reported | Averted | Reported | Expected | % change | Reported | Averted | Reported | Expected | % change | Reported | Averted | Reported | Expected | % change |
| | 50-59 | 348 | 26 | 8 | 8 | 7 | 89 | 94 | 2 | 4 | 51 | | | | | |
| | 60+ | 16,032 | 3,195 | 82 | 99 | 17 | 4,920 | 6,453 | 25 | 58 | 57 | | | | | |
| | 60-69 | 1,422 | 108 | 28 | 30 | 7 | 300 | 435 | 6 | 15 | 59 | | | | | |
| | 70-79 | 1,800 | 262 | 59 | 67 | 13 | 542 | 792 | 18 | 44 | 59 | | | | | |
| | 80+ | 2,122 | 695 | 131 | 174 | 25 | 798 | 924 | 49 | 106 | 54 | | | | | |
| Portugal | 25-49 | 15 | 0 | 0 | 0 | 0 | 43 | 117 | 1 | 5 | 73 | 106 | 274 | 3 | 11 | 72 |
| | 50-59 | 45 | 0 | 3 | 3 | 0 | 87 | 310 | 6 | 27 | 78 | 203 | 574 | 13 | 52 | 74 |
| | 60+ | 955 | 131 | 30 | 35 | 12 | 1,835 | 7,745 | 58 | 304 | 81 | 7,092 | 20,633 | 221 | 865 | 74 |
| | 60-69 | 128 | 1 | 9 | 9 | 1 | 214 | 793 | 16 | 74 | 79 | 515 | 1,548 | 37 | 149 | 75 |
| | 70-79 | 279 | 13 | 26 | 27 | 4 | 487 | 1,855 | 46 | 220 | 79 | 1,386 | 3,958 | 126 | 487 | 74 |
| | 80+ | 548 | 117 | 76 | 93 | 18 | 1,134 | 5,097 | 158 | 869 | 82 | 5,191 | 15,127 | 717 | 2,806 | 74 |
| Republic of Moldova | 60+ | 1,322 | 10 | 187 | 188 | 1 | 2,060 | 990 | 291 | 430 | 32 | 1,467 | 725 | 207 | 309 | 33 |
| Rom | 25-49 | 15 | 0 | 0 | 0 | 0 | 1,252 | 520 | 19 | 27 | 29 | 197 | 110 | 3 | 5 | 36 |

| Countries, areas and territories | Age group (years) | Alpha | | | | | Delta | | | | | Omicron | | | | |
|---|---|---|---|---|---|---|---|---|---|---|---|---|---|---|---|---|
| | | Total deaths | | Mortality rates | | | Total deaths | | Mortality rates | | | Total deaths | | Mortality rates | | |
| | | Reported | Averted | Reported | Expected | % change | Reported | Averted | Reported | Expected | % change | Reported | Averted | Reported | Expected | % change |
| | 50-59 | 37 | 6 | 1 | 2 | 14 | 2,261 | 1,138 | 87 | 131 | 33 | 409 | 240 | 15 | 24 | 37 |
| | 60+ | 363 | 50 | 7 | 8 | 12 | 20,890 | 8,351 | 423 | 593 | 29 | 6,188 | 2,510 | 126 | 177 | 29 |
| | 60-69 | 89 | 16 | 4 | 4 | 15 | 6,035 | 3,245 | 247 | 380 | 35 | 1,259 | 758 | 51 | 81 | 38 |
| | 70-79 | 141 | 25 | 9 | 11 | 15 | 8,094 | 3,689 | 517 | 752 | 31 | 2,130 | 1,035 | 133 | 198 | 33 |
| | 80+ | 133 | 9 | 14 | 15 | 6 | 6,761 | 1,417 | 731 | 884 | 17 | 2,799 | 717 | 331 | 415 | 20 |
| Slovakia | 25-49 | 336 | 0 | 16 | 16 | 0 | 223 | 148 | 11 | 18 | 40 | 63 | 29 | 3 | 5 | 32 |
| | 50-59 | 721 | 24 | 103 | 106 | 3 | 428 | 378 | 61 | 115 | 47 | 111 | 86 | 16 | 28 | 44 |
| | 60+ | 7,615 | 398 | 592 | 623 | 5 | 4,644 | 4,462 | 359 | 704 | 49 | 1,955 | 1,956 | 151 | 302 | 50 |
| | 60-69 | 2,164 | 79 | 314 | 326 | 4 | 1,220 | 1,260 | 178 | 361 | 51 | 363 | 409 | 53 | 112 | 53 |
| | 70-79 | 2,900 | 164 | 711 | 751 | 5 | 1,688 | 1,959 | 399 | 862 | 54 | 656 | 780 | 155 | 340 | 54 |
| | 80+ | 2,551 | 155 | 1,349 | 1,431 | 6 | 1,736 | 1,243 | 945 | 1,622 | 42 | 936 | 767 | 510 | 927 | 45 |
| Slovenia | 25-49 | 19 | 0 | 3 | 3 | 0 | 24 | 12 | 3 | 5 | 33 | 40 | 16 | 6 | 8 | 29 |
| | 50-59 | 43 | 0 | 14 | 14 | 0 | 63 | 71 | 21 | 44 | 53 | 96 | 88 | 32 | 61 | 48 |
| | 60+ | 996 | 199 | 172 | 206 | 17 | 1,235 | 1,640 | 213 | 496 | 57 | 3,001 | 3,515 | 510 | 1,108 | 54 |
| | 60-69 | 159 | 9 | 57 | 60 | 5 | 185 | 255 | 66 | 157 | 58 | 348 | 435 | 123 | 278 | 56 |

| | | Alpha | | | | | Delta | | | | | Omicron | | | | |
|---|---|---|---|---|---|---|---|---|---|---|---|---|---|---|---|---|
| | | Total deaths | | Mortality rates | | | Total deaths | | Mortality rates | | | Total deaths | | Mortality rates | | |
| Countries, areas and territories | Age group (years) | Reported | Averted | Reported | Expected | % change | Reported | Averted | Reported | Expected | % change | Reported | Averted | Reported | Expected | % change |
| | 70-79 | 267 | 27 | 145 | 160 | 9 | 320 | 540 | 174 | 469 | 63 | 643 | 947 | 340 | 842 | 60 |
| | 80+ | 570 | 163 | 493 | 634 | 22 | 730 | 845 | 631 | 1,362 | 54 | 2,010 | 2,133 | 1,710 | 3,525 | 51 |
| Spain | 25-49 | 374 | 6 | 2 | 2 | 2 | 259 | 343 | 2 | 4 | 57 | 461 | 770 | 3 | 8 | 63 |
| | 50-59 | 1,177 | 40 | 17 | 17 | 3 | 404 | 1,474 | 6 | 26 | 78 | 1,039 | 2,134 | 14 | 44 | 67 |
| | 60+ | 25,616 | 4,236 | 207 | 241 | 14 | 7,067 | 40,211 | 57 | 382 | 85 | 28,897 | 86,116 | 229 | 911 | 75 |
| | 60-69 | 3,131 | 299 | 57 | 62 | 9 | 915 | 3,301 | 17 | 77 | 78 | 2,779 | 6,722 | 49 | 168 | 71 |
| | 70-79 | 6,436 | 631 | 161 | 177 | 9 | 1,404 | 7,796 | 35 | 230 | 85 | 5,801 | 16,650 | 142 | 549 | 74 |
| | 80+ | 16,049 | 3,306 | 557 | 671 | 17 | 4,748 | 29,114 | 165 | 1,174 | 86 | 20,317 | 62,744 | 705 | 2,881 | 76 |
| Sweden | 25-49 | 72 | 0 | 2 | 2 | 0 | 20 | 17 | 1 | 1 | 46 | 98 | 113 | 3 | 6 | 54 |
| | 50-59 | 157 | 2 | 12 | 12 | 1 | 21 | 55 | 2 | 6 | 72 | 184 | 303 | 14 | 36 | 62 |
| | 60+ | 4,308 | 578 | 160 | 182 | 12 | 574 | 1,513 | 21 | 78 | 72 | 8,226 | 15,973 | 302 | 889 | 66 |
| | 60-69 | 440 | 41 | 40 | 43 | 9 | 53 | 166 | 5 | 20 | 76 | 518 | 1,121 | 46 | 147 | 68 |
| | 70-79 | 1,161 | 138 | 114 | 127 | 11 | 156 | 514 | 15 | 66 | 77 | 1,952 | 4,893 | 191 | 670 | 71 |
| | 80+ | 2,707 | 399 | 484 | 555 | 13 | 365 | 833 | 65 | 214 | 70 | 5,756 | 9,959 | 988 | 2,699 | 63 |

| | | Alpha | | | | | Delta | | | | | Omicron | | | | |
|---|---|---|---|---|---|---|---|---|---|---|---|---|---|---|---|---|
| | | Total deaths | | Mortality rates | | | Total deaths | | Mortality rates | | | Total deaths | | Mortality rates | | |
| Countries, areas and territories | Age group (years) | Reported | Averted | Reported | Expected | % change | Reported | Averted | Reported | Expected | % change | Reported | Averted | Reported | Expected | % change |
| Switzerland | 25-49 | 10 | 0 | 0 | 0 | 0 | 30 | 30 | 1 | 2 | 50 | 29 | 17 | 1 | 2 | 37 |
| | 50-59 | 48 | 0 | 4 | 4 | 0 | 70 | 103 | 5 | 13 | 60 | 71 | 87 | 5 | 12 | 55 |
| | 60+ | 764 | 369 | 35 | 51 | 33 | 1,612 | 3,772 | 73 | 244 | 70 | 1,901 | 4,479 | 86 | 289 | 70 |
| | 60-69 | 117 | 12 | 12 | 13 | 9 | 169 | 300 | 17 | 47 | 64 | 168 | 266 | 17 | 44 | 61 |
| | 70-79 | 220 | 71 | 30 | 39 | 24 | 332 | 744 | 45 | 145 | 69 | 420 | 926 | 56 | 181 | 69 |
| | 80+ | 427 | 286 | 90 | 151 | 40 | 1,111 | 2,728 | 235 | 813 | 71 | 1,313 | 3,287 | 278 | 975 | 71 |
| Ukraine | 60+ | 22,793 | 145 | 249 | 251 | 1 | 36,584 | 8,330 | 400 | 491 | 19 | 13,299 | 5,595 | 145 | 206 | 30 |
| United Kingdom (England) | 25-49 | 1,677 | 46 | 9 | 9 | 3 | 1,301 | 2,264 | 7 | 19 | 64 | 2,158 | 3,609 | 12 | 31 | 63 |
| | 50-59 | 3,514 | 234 | 45 | 49 | 6 | 1,733 | 5,654 | 22 | 96 | 77 | 3,506 | 8,385 | 45 | 154 | 71 |
| | 60+ | 58,904 | 23,427 | 428 | 599 | 28 | 17,769 | 95,635 | 129 | 825 | 84 | 77,789 | 257,278 | 566 | 2,437 | 77 |
| | 60-69 | 7,295 | 721 | 120 | 132 | 9 | 3,074 | 15,330 | 51 | 304 | 83 | 7,769 | 24,442 | 128 | 532 | 76 |
| | 70-79 | 14,401 | 3,036 | 295 | 358 | 17 | 5,099 | 26,052 | 105 | 639 | 84 | 18,644 | 60,879 | 382 | 1,631 | 77 |
| | 80+ | 37,208 | 19,670 | 1,321 | 2,019 | 35 | 9,596 | 54,253 | 341 | 2,267 | 85 | 51,376 | 171,957 | 1,824 | 7,930 | 77 |

| | | Alpha | | | | | Delta | | | | | Omicron | | | | |
|---|---|---|---|---|---|---|---|---|---|---|---|---|---|---|---|---|
| | | Total deaths | | Mortality rates | | | Total deaths | | Mortality rates | | | Total deaths | | Mortality rates | | |
| Countries, areas and territories | Age group (years) | Reported | Averted | Reported | Expected | % change | Reported | Averted | Reported | Expected | % change | Reported | Averted | Reported | Expected | % change |
| United Kingdom (Scotland) | 25-49 | 61 | 0 | 3 | 3 | 0 | 119 | 220 | 7 | 19 | 65 | 123 | 193 | 7 | 18 | 61 |
| | 50-59 | 176 | 6 | 22 | 23 | 3 | 162 | 638 | 20 | 101 | 80 | 208 | 530 | 26 | 93 | 72 |
| | 60+ | 2,923 | 308 | 204 | 225 | 10 | 1,889 | 7,220 | 132 | 635 | 79 | 4,484 | 15,225 | 313 | 1,374 | 77 |
| United Kingdom (Wales) | 25-49 | 52 | 0 | 6 | 6 | 0 | 29 | 86 | 3 | 12 | 75 | | | | | |
| | 50-59 | 130 | 1 | 30 | 30 | 1 | 82 | 320 | 19 | 92 | 80 | | | | | |
| | 60+ | 6,372 | 717 | 368 | 409 | 10 | 2,586 | 9,156 | 149 | 678 | 78 | | | | | |
| | 60-69 | 291 | 19 | 77 | 82 | 6 | 161 | 790 | 43 | 251 | 83 | | | | | |
| | 70-79 | 589 | 63 | 187 | 207 | 10 | 266 | 1,091 | 84 | 431 | 80 | | | | | |
| | 80+ | 1,244 | 157 | 720 | 811 | 11 | 435 | 1,171 | 252 | 929 | 73 | | | | | |
| Kosovo[1] | 25-49 | 31 | 0 | 5 | 5 | 0 | 40 | 5 | 6 | 7 | 11 | 6 | 3 | 1 | 1 | 33 |
| | 50-59 | 82 | 0 | 43 | 43 | 0 | 58 | 18 | 29 | 39 | 24 | 22 | 16 | 11 | 19 | 42 |
| | 60+ | 728 | 0 | 301 | 301 | 0 | 348 | 286 | 139 | 254 | 45 | 153 | 122 | 61 | 110 | 44 |
| | 60-69 | 193 | 0 | 145 | 145 | 0 | 103 | 67 | 74 | 122 | 39 | 34 | 34 | 24 | 49 | 50 |
| | 70-79 | 317 | 0 | 431 | 431 | 0 | 142 | 159 | 189 | 400 | 53 | 63 | 63 | 84 | 168 | 50 |

| | | Alpha | | | | | Delta | | | | | Omicron | | | | |
|---|---|---|---|---|---|---|---|---|---|---|---|---|---|---|---|---|
| | | Total deaths | | Mortality rates | | | Total deaths | | Mortality rates | | | Total deaths | | Mortality rates | | |
| Countries, areas and territories | Age group (years) | Reported | Averted | Reported | Expected | % change | Reported | Averted | Reported | Expected | % change | Reported | Averted | Reported | Expected | % change |
| | 80+ | 218 | 0 | 633 | 633 | 0 | 103 | 60 | 294 | 465 | 37 | 56 | 25 | 160 | 231 | 31 |

Mortality rates are per 100,000 population.

[1] All references to Kosovo in this document should be understood to be in the context of the United Nations Security Council resolution 1244 (1999).

*Supplementary table 4: Cumulative Vaccination Uptake (VU), as well as reported mortality counts, lives saved per vaccine dose and mortality rates per 100,000 population, by countries, areas and territories (CAT), included in the analysis. Note: $d_1$ refers to first dose, $d_2$, refers to complete series, $d_3$ refers to first additional booster, $d_4$ refers to second additional booster and $d_5$ refers to third additional booster; the ≥60 years age group includes data for all 34 CAT.*

| | | Vaccination uptake | | | | Number of lives | | | | | | | Mortality rate per 100,000 | | |
|---|---|---|---|---|---|---|---|---|---|---|---|---|---|---|---|
| **Countries, areas and territories** | **Age group (years)** | **$VU_2$** | **$VU_3$** | **$VU_4$** | **$VU_5$** | **Reported deaths** | **Saved after $d_1$** | **Saved after $d_2$** | **Saved after $d_3$** | **Saved after $d_4$** | **Saved after $d_5$** | **Total averted** | **Reported** | **Total Expected** | **% change** |
| Austria | 25-49 | 72 | 61 | 12 | <1% | 272 | 17 | 87 | 93 | 0 | 0 | 197 | 9 | 16 | 44 |
| | 50-59 | 80 | 74 | 22 | 1 | 687 | 52 | 249 | 310 | 12 | 0 | 623 | 49 | 94 | 48 |
| | 60+ | 88 | 83 | 44 | 2 | 16,259 | 855 | 6,359 | 12,937 | 1,911 | 32 | 22,094 | 691 | 1,631 | 58 |
| | 60-69 | 84 | 79 | 37 | 1 | 1,837 | 97 | 686 | 955 | 91 | 0 | 1,829 | 169 | 337 | 50 |
| | 70-79 | 89 | 85 | 49 | 2 | 3,882 | 189 | 1,235 | 2,415 | 377 | 3 | 4,219 | 523 | 1,092 | 52 |
| | 80+ | 95 | 86 | 51 | 2 | 10,540 | 569 | 4,438 | 9,567 | 1,443 | 29 | 16,046 | 2,011 | 5,072 | 60 |
| Belgium | 25-49 | 79 | 66 | 23 | 1 | 98 | 3 | 62 | 19 | 0 | 0 | 84 | 3 | 5 | 40 |
| | 50-59 | 86 | 82 | 48 | 3 | 516 | 38 | 293 | 257 | 25 | 0 | 613 | 33 | 71 | 54 |
| | 60+ | 94 | 91 | 70 | 14 | 15,307 | 853 | 6,172 | 15,157 | 2,504 | 1,014 | 25,700 | 508 | 1,360 | 63 |
| | 60-69 | 92 | 89 | 65 | 5 | 1,814 | 95 | 1,063 | 1,214 | 242 | 19 | 2,633 | 130 | 320 | 59 |
| | 70-79 | 95 | 92 | 74 | 8 | 3,749 | 209 | 1,866 | 3,456 | 624 | 62 | 6,217 | 382 | 1,016 | 62 |
| | 80+ | 98 | 94 | 74 | 41 | 9,744 | 549 | 3,243 | 10,487 | 1,638 | 933 | 16,850 | 1,514 | 4,131 | 63 |
| Croatia | 25-49 | 54 | 16 | 0 | <1% | 81 | 3 | 13 | 3 | 0 | 0 | 19 | 7 | 8 | 12 |

| Countries, areas and territories | Age group (years) | Vaccination uptake | | | | Number of lives | | | | | | | Mortality rate per 100,000 | | |
|---|---|---|---|---|---|---|---|---|---|---|---|---|---|---|---|
| | | $VU_2$ | $VU_3$ | $VU_4$ | $VU_5$ | Reported deaths | Saved after $d_1$ | Saved after $d_2$ | Saved after $d_3$ | Saved after $d_4$ | Saved after $d_5$ | Total averted | Reported | Total Expected | % change |
| | 50-59 | 68 | 30 | 1 | <1% | 203 | 13 | 88 | 43 | 0 | 0 | 144 | 38 | 65 | 42 |
| | 60+ | 75 | 53 | 5 | 1 | 4,521 | 209 | 1,342 | 1,836 | 52 | 14 | 3,453 | 391 | 690 | 43 |
| | 60-69 | 76 | 47 | 3 | <1% | 716 | 41 | 295 | 268 | 0 | 0 | 604 | 126 | 233 | 46 |
| | 70-79 | 78 | 62 | 7 | 2 | 1,369 | 67 | 479 | 676 | 18 | 3 | 1,243 | 364 | 694 | 48 |
| | 80+ | 66 | 53 | 7 | 2 | 2,436 | 101 | 568 | 892 | 34 | 11 | 1,606 | 1,151 | 1,910 | 40 |
| Cyprus | 25-49 | 78 | 59 | 2 | <1% | 46 | 0 | 23 | 6 | 0 | 0 | 29 | 13 | 22 | 41 |
| | 50-59 | 82 | 74 | 6 | <1% | 75 | 0 | 51 | 43 | 0 | 0 | 94 | 69 | 155 | 55 |
| | 60+ | 90 | 85 | 29 | 7 | 1,369 | 84 | 985 | 1,294 | 158 | 14 | 2,535 | 680 | 1,939 | 65 |
| | 60-69 | 88 | 82 | 20 | 3 | 211 | 4 | 124 | 128 | 0 | 0 | 256 | 218 | 483 | 55 |
| | 70-79 | 93 | 90 | 38 | 10 | 387 | 20 | 312 | 378 | 39 | 0 | 749 | 561 | 1,647 | 66 |
| | 80+ | 91 | 84 | 38 | 11 | 771 | 60 | 549 | 788 | 119 | 14 | 1,530 | 2,166 | 6,463 | 66 |
| Czechia | 25-49 | 64 | 37 | 2 | <1% | 623 | 11 | 162 | 47 | 0 | 0 | 220 | 17 | 23 | 26 |
| | 50-59 | 72 | 54 | 6 | <1% | 1,398 | 34 | 418 | 188 | 0 | 0 | 640 | 102 | 149 | 32 |
| | 60+ | 83 | 73 | 23 | <1% | 31,271 | 1,293 | 8,936 | 12,854 | 720 | 0 | 23,803 | 1,129 | 1,989 | 43 |
| | 60-69 | 79 | 69 | 16 | <1% | 4,888 | 146 | 1,186 | 978 | 29 | 0 | 2,339 | 388 | 574 | 32 |

| | | Vaccination uptake | | | | Number of lives | | | | | | | Mortality rate per 100,000 | | |
|---|---|---|---|---|---|---|---|---|---|---|---|---|---|---|---|
| Countries, areas and territories | Age group (years) | $VU_2$ | $VU_3$ | $VU_4$ | $VU_5$ | Reported deaths | Saved after $d_1$ | Saved after $d_2$ | Saved after $d_3$ | Saved after $d_4$ | Saved after $d_5$ | Total averted | Reported | Total Expected | % change |
| | 70-79 | 86 | 77 | 27 | <1% | 11,788 | 441 | 2,971 | 4,295 | 227 | 0 | 7,934 | 1,114 | 1,864 | 40 |
| | 80+ | 87 | 75 | 30 | <1% | 14,595 | 706 | 4,779 | 7,581 | 464 | 0 | 13,530 | 3,235 | 6,234 | 48 |
| Denmark | 25-49 | 84 | 63 | 5 | <1% | 130 | 0 | 62 | 92 | 0 | 0 | 154 | 7 | 15 | 53 |
| | 50-59 | 93 | 87 | 58 | 1 | 217 | 0 | 116 | 255 | 38 | 0 | 409 | 27 | 78 | 65 |
| | 60+ | 99 | 97 | 86 | 2 | 6,772 | 0 | 1,518 | 10,045 | 3,004 | 70 | 14,637 | 438 | 1,384 | 68 |
| | 60-69 | 97 | 94 | 79 | 1 | 656 | 0 | 268 | 923 | 246 | 0 | 1,437 | 97 | 311 | 69 |
| | 70-79 | 98 | 97 | 89 | 3 | 1,823 | 0 | 493 | 2,680 | 785 | 24 | 3,982 | 314 | 999 | 69 |
| | 80+ | 100 | 100 | 96 | 3 | 4,293 | 0 | 757 | 6,442 | 1,973 | 46 | 9,218 | 1,473 | 4,635 | 68 |
| Estonia | 25-49 | 57 | 34 | 5 | <1% | 59 | 0 | 14 | 4 | 0 | 0 | 18 | 13 | 17 | 24 |
| | 50-59 | 67 | 46 | 8 | <1% | 107 | 0 | 31 | 21 | 0 | 0 | 52 | 61 | 91 | 33 |
| | 60+ | 77 | 60 | 19 | 1 | 2,467 | 120 | 1,009 | 1,171 | 116 | 0 | 2,416 | 677 | 1,341 | 50 |
| | 60-69 | 72 | 54 | 15 | 1 | 330 | 5 | 97 | 89 | 0 | 0 | 191 | 196 | 310 | 37 |
| | 70-79 | 80 | 65 | 22 | 1 | 633 | 23 | 245 | 264 | 23 | 0 | 555 | 546 | 1,026 | 47 |
| | 80+ | 82 | 64 | 23 | 1 | 1,504 | 92 | 667 | 818 | 93 | 0 | 1,670 | 1,876 | 3,958 | 53 |
| Finland | 25-49 | 83 | 52 | 3 | <1% | 157 | 4 | 100 | 97 | 0 | 0 | 201 | 9 | 20 | 55 |

| | | Vaccination uptake | | | | Number of lives | | | | | | | Mortality rate per 100,000 | | |
|---|---|---|---|---|---|---|---|---|---|---|---|---|---|---|---|
| Countries, areas and territories | Age group (years) | $VU_2$ | $VU_3$ | $VU_4$ | $VU_5$ | Reported deaths | Saved after $d_1$ | Saved after $d_2$ | Saved after $d_3$ | Saved after $d_4$ | Saved after $d_5$ | Total averted | Reported | Total Expected | % change |
| | 50-59 | 92 | 77 | 11 | 2 | 226 | 3 | 117 | 255 | 7 | 0 | 382 | 32 | 86 | 63 |
| | 60+ | 94 | 89 | 68 | 32 | 8,125 | 155 | 2,100 | 7,258 | 5,851 | 1,592 | 16,956 | 497 | 1,535 | 68 |
| | 60-69 | 94 | 87 | 54 | 15 | 647 | 12 | 285 | 786 | 212 | 32 | 1,327 | 91 | 278 | 67 |
| | 70-79 | 96 | 92 | 79 | 41 | 1,895 | 45 | 608 | 2,180 | 1,098 | 356 | 4,287 | 319 | 1,040 | 69 |
| | 80+ | 92 | 91 | 81 | 49 | 5,583 | 98 | 1,207 | 4,292 | 4,541 | 1,204 | 11,342 | 1,688 | 5,118 | 67 |
| France | 25-49 | 89 | 68 | 4 | <1% | 1,730 | 171 | 740 | 1,192 | 3 | 0 | 2,106 | 8 | 19 | 58 |
| | 50-59 | 91 | 80 | 13 | 1 | 3,998 | 304 | 1,316 | 3,199 | 116 | 0 | 4,935 | 45 | 100 | 55 |
| | 60+ | 90 | 83 | 44 | 11 | 93,100 | 4,008 | 24,119 | 62,470 | 12,524 | 1,505 | 104,626 | 506 | 1,074 | 53 |
| | 60-69 | 91 | 84 | 38 | 8 | 11,431 | 690 | 3,239 | 8,453 | 973 | 69 | 13,424 | 141 | 307 | 54 |
| | 70-79 | 90 | 85 | 49 | 13 | 22,785 | 1,036 | 6,935 | 16,537 | 2,854 | 286 | 27,648 | 366 | 809 | 55 |
| | 80+ | 88 | 80 | 49 | 16 | 58,884 | 2,282 | 13,945 | 37,480 | 8,697 | 1,150 | 63,554 | 1,441 | 2,997 | 52 |
| Germany | 60+ | 90 | 85 | 39 | 8 | 135,420 | 2,632 | 47,891 | 82,445 | 17,067 | 4,422 | 154,457 | 554 | 1,187 | 53 |
| Greece | 25-49 | 70 | 60 | 2 | <1% | 843 | 95 | 354 | 220 | 0 | 0 | 669 | 25 | 45 | 44 |
| | 50-59 | 77 | 70 | 7 | 1 | 1,939 | 130 | 843 | 752 | 0 | 0 | 1,725 | 126 | 237 | 47 |
| | 60+ | 88 | 80 | 27 | 6 | 30,663 | 1,878 | 12,764 | 23,473 | 3,092 | 360 | 41,567 | 999 | 2,353 | 58 |

| Countries, areas and territories | Age group (years) | Vaccination uptake | | | | Number of lives | | | | | | | Mortality rate per 100,000 | | |
|---|---|---|---|---|---|---|---|---|---|---|---|---|---|---|---|
| | | VU2 | VU3 | VU4 | VU5 | Reported deaths | Saved after d1 | Saved after d2 | Saved after d3 | Saved after d4 | Saved after d5 | Total averted | Reported | Total Expected | % change |
| | 60-69 | 86 | 78 | 21 | 4 | 4,613 | 330 | 2,157 | 2,665 | 136 | 14 | 5,302 | 349 | 750 | 53 |
| | 70-79 | 91 | 84 | 33 | 8 | 7,969 | 264 | 3,408 | 6,174 | 706 | 84 | 10,636 | 802 | 1,872 | 57 |
| | 80+ | 87 | 77 | 30 | 7 | 18,081 | 1,284 | 7,199 | 14,634 | 2,250 | 262 | 25,629 | 2,398 | 5,796 | 59 |
| Hungary | 25-49 | 63 | 36 | 1 | <1% | 1,798 | 66 | 525 | 254 | 0 | 0 | 845 | 52 | 77 | 32 |
| | 50-59 | 70 | 50 | 3 | <1% | 3,423 | 112 | 924 | 894 | 0 | 0 | 1,930 | 270 | 422 | 36 |
| | 60+ | 81 | 68 | 13 | <1% | 36,007 | 761 | 7,237 | 19,113 | 1,137 | 0 | 28,248 | 1,401 | 2,500 | 44 |
| | 60-69 | 81 | 65 | 9 | <1% | 8,408 | 242 | 2,110 | 3,893 | 107 | 0 | 6,352 | 678 | 1,190 | 43 |
| | 70-79 | 84 | 74 | 17 | 1 | 12,262 | 244 | 2,563 | 7,782 | 450 | 0 | 11,039 | 1,385 | 2,632 | 47 |
| | 80+ | 75 | 64 | 15 | 1 | 15,337 | 275 | 2,564 | 7,438 | 580 | 0 | 10,857 | 3,444 | 5,882 | 41 |
| Iceland | 25-49 | 57 | 81 | 4 | <1% | 1 | 0 | 0 | 3 | 0 | 0 | 3 | 1 | 3 | 67 |
| | 50-59 | 84 | 90 | 13 | 1 | 6 | 0 | 0 | 10 | 0 | 0 | 10 | 14 | 36 | 61 |
| | 60+ | 96 | 93 | 68 | 21 | 228 | 0 | 6 | 385 | 107 | 1 | 499 | 295 | 940 | 69 |
| | 60-69 | 93 | 91 | 58 | 13 | 13 | 0 | 0 | 23 | 2 | 0 | 25 | 33 | 98 | 66 |
| | 70-79 | 97 | 94 | 76 | 28 | 56 | 0 | 6 | 101 | 27 | 0 | 134 | 221 | 751 | 71 |
| | 80+ | 100 | 94 | 84 | 32 | 159 | 0 | 0 | 261 | 78 | 1 | 340 | 1,215 | 3,812 | 68 |

| | | Vaccination uptake | | | | Number of lives | | | | | | | Mortality rate per 100,000 | | |
|---|---|---|---|---|---|---|---|---|---|---|---|---|---|---|---|
| Countries, areas and territories | Age group (years) | $VU_2$ | $VU_3$ | $VU_4$ | $VU_5$ | Reported deaths | Saved after $d_1$ | Saved after $d_2$ | Saved after $d_3$ | Saved after $d_4$ | Saved after $d_5$ | Total averted | Reported | Total Expected | % change |
| Ireland | 25-49 | 85 | 65 | 12 | <1% | 143 | 2 | 81 | 70 | 0 | 0 | 153 | 8 | 17 | 53 |
| | 50-59 | 87 | 87 | 39 | 2 | 239 | 49 | 174 | 202 | 21 | 0 | 446 | 38 | 109 | 65 |
| | 60+ | 100 | 100 | 76 | 38 | 5,620 | 62 | 2,036 | 3,537 | 1,542 | 782 | 7,959 | 545 | 1,317 | 59 |
| | 60-69 | 95 | 91 | 60 | 19 | 638 | 23 | 494 | 407 | 119 | 27 | 1,070 | 127 | 339 | 63 |
| | 70-79 | 100 | 100 | 87 | 55 | 1,448 | 24 | 869 | 1,013 | 455 | 209 | 2,570 | 418 | 1,161 | 64 |
| | 80+ | 100 | 100 | 95 | 60 | 3,534 | 15 | 673 | 2,117 | 968 | 546 | 4,319 | 1,943 | 4,317 | 55 |
| Israel | 25-49 | 89 | 66 | 4 | <1% | 270 | 19 | 174 | 193 | 0 | 0 | 386 | 10 | 24 | 58 |
| | 50-59 | 95 | 81 | 10 | <1% | 458 | 41 | 374 | 579 | 10 | 0 | 1,004 | 58 | 184 | 68 |
| | 60+ | 100 | 100 | 57 | 1 | 8,669 | 0 | 5,591 | 13,437 | 5,809 | 20 | 24,857 | 667 | 2,579 | 74 |
| Italy | 25-49 | 85 | 81 | 3 | <1% | 1,801 | 148 | 486 | 1,058 | 0 | 0 | 1,692 | 10 | 19 | 47 |
| | 50-59 | 85 | 87 | 6 | <1% | 4,795 | 432 | 1,118 | 2,881 | 33 | 0 | 4,464 | 50 | 97 | 48 |
| | 60+ | 90 | 92 | 31 | 3 | 124,987 | 7,676 | 31,039 | 106,230 | 19,933 | 842 | 165,720 | 690 | 1,606 | 57 |
| | 60-69 | 85 | 91 | 21 | 1 | 12,880 | 1,081 | 2,533 | 7,915 | 479 | 15 | 12,023 | 170 | 329 | 48 |
| | 70-79 | 90 | 93 | 33 | 2 | 30,768 | 2,227 | 5,617 | 22,140 | 2,384 | 64 | 32,432 | 511 | 1,050 | 51 |
| | 80+ | 96 | 93 | 46 | 6 | 81,339 | 4,368 | 22,889 | 76,175 | 17,070 | 763 | 121,265 | 1,805 | 4,497 | 60 |

| Countries, areas and territories | Age group (years) | Vaccination uptake | | | | Number of lives | | | | | | | Mortality rate per 100,000 | | |
|---|---|---|---|---|---|---|---|---|---|---|---|---|---|---|---|
| | | $VU_2$ | $VU_3$ | $VU_4$ | $VU_5$ | Reported deaths | Saved after $d_1$ | Saved after $d_2$ | Saved after $d_3$ | Saved after $d_4$ | Saved after $d_5$ | Total averted | Reported | Total Expected | % change |
| Latvia | 25-49 | 61 | 32 | 2 | <1% | 222 | 46 | 93 | 27 | 0 | 0 | 166 | 36 | 63 | 43 |
| | 50-59 | 62 | 36 | 3 | <1% | 415 | 79 | 159 | 56 | 0 | 0 | 294 | 161 | 275 | 41 |
| | 60+ | 63 | 43 | 9 | 2 | 5,022 | 601 | 1,533 | 1,022 | 5 | 0 | 3,161 | 954 | 1,554 | 39 |
| | 60-69 | 65 | 42 | 6 | 1 | 936 | 146 | 393 | 170 | 0 | 0 | 709 | 379 | 666 | 43 |
| | 70-79 | 67 | 48 | 11 | 3 | 1,481 | 180 | 553 | 361 | 0 | 0 | 1,094 | 892 | 1,551 | 42 |
| | 80+ | 54 | 39 | 10 | 3 | 2,605 | 275 | 587 | 491 | 5 | 0 | 1,358 | 2,298 | 3,496 | 34 |
| Lithuania | 25-49 | 63 | 32 | 1 | <1% | 291 | 56 | 194 | 19 | 0 | 0 | 269 | 32 | 61 | 48 |
| | 50-59 | 67 | 37 | 1 | <1% | 512 | 72 | 325 | 32 | 0 | 0 | 429 | 122 | 225 | 46 |
| | 60+ | 71 | 51 | 3 | <1% | 7,392 | 391 | 3,620 | 1,580 | 0 | 0 | 5,591 | 949 | 1,667 | 43 |
| | 60-69 | 72 | 47 | 2 | <1% | 1,155 | 110 | 786 | 173 | 0 | 0 | 1,069 | 304 | 586 | 48 |
| | 70-79 | 75 | 57 | 4 | <1% | 2,167 | 127 | 1,394 | 536 | 0 | 0 | 2,057 | 900 | 1,754 | 49 |
| | 80+ | 64 | 49 | 5 | <1% | 4,070 | 154 | 1,440 | 871 | 0 | 0 | 2,465 | 2,575 | 4,135 | 38 |
| Luxembourg | 25-49 | 68 | 59 | 3 | <1% | 13 | 0 | 0 | 0 | 0 | 0 | 0 | 5 | 5 | 0 |
| | 50-59 | 74 | 74 | 7 | <1% | 35 | 0 | 10 | 21 | 0 | 0 | 31 | 38 | 72 | 47 |
| | 60+ | 86 | 83 | 48 | 1 | 706 | 14 | 222 | 665 | 132 | 0 | 1,033 | 535 | 1,318 | 59 |

| Countries, areas and territories | Age group (years) | Vaccination uptake VU$_2$ | VU$_3$ | VU$_4$ | VU$_5$ | Number of lives: Reported deaths | Saved after d$_1$ | Saved after d$_2$ | Saved after d$_3$ | Saved after d$_4$ | Saved after d$_5$ | Total averted | Mortality rate per 100,000: Reported | Total Expected | % change |
|---|---|---|---|---|---|---|---|---|---|---|---|---|---|---|---|
| | 60-69 | 80 | 80 | 39 | 1 | 85 | 0 | 21 | 50 | 8 | 0 | 79 | 129 | 248 | 48 |
| | 70-79 | 87 | 84 | 53 | <1% | 148 | 3 | 35 | 94 | 13 | 0 | 145 | 365 | 723 | 50 |
| | 80+ | 97 | 90 | 63 | 1 | 473 | 11 | 166 | 521 | 111 | 0 | 809 | 1,862 | 5,046 | 63 |
| Malta | 25-49 | 82 | 78 | 1 | <1% | 14 | 0 | 6 | 24 | 0 | 0 | 30 | 7 | 21 | 67 |
| | 50-59 | 86 | 89 | 4 | <1% | 10 | 0 | 0 | 18 | 0 | 0 | 18 | 17 | 48 | 65 |
| | 60+ | 97 | 87 | 44 | 5 | 480 | 0 | 314 | 599 | 186 | 0 | 1,099 | 372 | 1,225 | 70 |
| | 60-69 | 93 | 95 | 26 | 3 | 68 | 0 | 49 | 103 | 1 | 0 | 153 | 114 | 370 | 69 |
| | 70-79 | 98 | 80 | 50 | 6 | 138 | 0 | 92 | 181 | 38 | 0 | 311 | 289 | 941 | 69 |
| | 80+ | 100 | 83 | 82 | 9 | 274 | 0 | 173 | 315 | 147 | 0 | 635 | 1,276 | 4,234 | 70 |
| Netherlands (Kingdom of) | 25-49 | 63 | 50 | 9 | <1% | 137 | 5 | 50 | 18 | 0 | 0 | 73 | 2 | 4 | 50 |
| | 50-59 | 70 | 70 | 23 | 1 | 352 | 51 | 131 | 91 | 0 | 0 | 273 | 14 | 25 | 44 |
| | 60+ | 94 | 93 | 70 | 46 | 12,501 | 286 | 11,752 | 3,122 | 1,327 | 422 | 16,909 | 267 | 628 | 57 |
| | 60-69 | 89 | 87 | 57 | 29 | 1,091 | 97 | 830 | 285 | 59 | 10 | 1,281 | 50 | 109 | 54 |
| | 70-79 | 97 | 96 | 80 | 59 | 3,150 | 99 | 2,991 | 734 | 297 | 90 | 4,211 | 190 | 445 | 57 |
| | 80+ | 100 | 100 | 86 | 65 | 8,260 | 90 | 7,931 | 2,103 | 971 | 322 | 11,417 | 968 | 2,307 | 58 |

| Countries, areas and territories | Age group (years) | Vaccination uptake | | | | Number of lives | | | | | | | Mortality rate per 100,000 | | |
|---|---|---|---|---|---|---|---|---|---|---|---|---|---|---|---|
| | | $VU_2$ | $VU_3$ | $VU_4$ | $VU_5$ | Reported deaths | Saved after $d_1$ | Saved after $d_2$ | Saved after $d_3$ | Saved after $d_4$ | Saved after $d_5$ | Total averted | Reported | Total Expected | % change |
| North Macedonia | 25-49 | 51 | 5 | 0 | <1% | 396 | 0 | 92 | 0 | 0 | 0 | 92 | 64 | 79 | 19 |
| | 50-59 | 61 | 10 | 0 | <1% | 714 | 0 | 212 | 7 | 0 | 0 | 219 | 282 | 368 | 23 |
| | 60+ | 66 | 24 | 2 | <1% | 6,482 | 87 | 2,364 | 459 | 0 | 0 | 2,910 | 1,473 | 2,134 | 31 |
| | 60-69 | 68 | 23 | 1 | <1% | 1,947 | 6 | 712 | 113 | 0 | 0 | 831 | 815 | 1,162 | 30 |
| | 70-79 | 69 | 29 | 3 | <1% | 2,647 | 36 | 1,090 | 247 | 0 | 0 | 1,373 | 1,825 | 2,771 | 34 |
| | 80+ | 51 | 18 | 2 | <1% | 1,888 | 45 | 562 | 99 | 0 | 0 | 706 | 3,366 | 4,625 | 27 |
| Portugal | 25-49 | 76 | 70 | 7 | <1% | 297 | 92 | 123 | 140 | 0 | 0 | 355 | 9 | 20 | 55 |
| | 50-59 | 78 | 88 | 43 | <1% | 611 | 162 | 225 | 388 | 27 | 0 | 802 | 41 | 94 | 56 |
| | 60+ | 89 | 92 | 70 | 12 | 20,439 | 1,773 | 4,870 | 14,560 | 3,241 | 1,443 | 25,887 | 638 | 1,445 | 56 |
| | 60-69 | 87 | 92 | 61 | 1 | 1,826 | 247 | 607 | 1,173 | 112 | 0 | 2,139 | 132 | 286 | 54 |
| | 70-79 | 90 | 92 | 74 | 1 | 4,467 | 341 | 1,272 | 3,249 | 441 | 1 | 5,304 | 407 | 890 | 54 |
| | 80+ | 90 | 93 | 82 | 51 | 14,146 | 1,185 | 2,991 | 10,138 | 2,688 | 1,442 | 18,444 | 1,953 | 4,500 | 57 |
| Republic of Moldova | 60+ | 39 | 22 | 2 | <1% | 5,983 | 272 | 1,156 | 243 | 0 | 0 | 1,671 | 844 | 1,080 | 22 |
| Romania | 25-49 | 37 | 10 | 0 | <1% | 2,283 | 144 | 454 | 20 | 0 | 0 | 618 | 36 | 46 | 22 |
| | 50-59 | 40 | 13 | 0 | <1% | 4,344 | 239 | 1,042 | 69 | 0 | 0 | 1,350 | 162 | 213 | 24 |

| | | Vaccination uptake | | | | Number of lives | | | | | | | Mortality rate per 100,000 | | |
|---|---|---|---|---|---|---|---|---|---|---|---|---|---|---|---|
| Countries, areas and territories | Age group (years) | $VU_2$ | $VU_3$ | $VU_4$ | $VU_5$ | Reported deaths | Saved after $d_1$ | Saved after $d_2$ | Saved after $d_3$ | Saved after $d_4$ | Saved after $d_5$ | Total averted | Reported | Total Expected | % change |
| | 60+ | 40 | 14 | 0 | <1% | 44,517 | 1,092 | 8,644 | 924 | 0 | 0 | 10,660 | 904 | 1,121 | 19 |
| | 60-69 | 45 | 15 | 0 | <1% | 12,296 | 442 | 3,203 | 278 | 0 | 0 | 3,923 | 496 | 655 | 24 |
| | 70-79 | 41 | 14 | 0 | <1% | 17,028 | 432 | 3,807 | 399 | 0 | 0 | 4,638 | 1,065 | 1,355 | 21 |
| | 80+ | 26 | 9 | 0 | <1% | 15,193 | 218 | 1,634 | 247 | 0 | 0 | 2,099 | 1,794 | 2,042 | 12 |
| Slovakia | 25-49 | 50 | 27 | 1 | <1% | 659 | 12 | 135 | 25 | 0 | 0 | 172 | 32 | 41 | 22 |
| | 50-59 | 59 | 38 | 2 | <1% | 1,365 | 35 | 339 | 95 | 0 | 0 | 469 | 194 | 261 | 26 |
| | 60+ | 68 | 59 | 4 | <1% | 15,744 | 151 | 3,034 | 3,348 | 0 | 0 | 6,533 | 1,218 | 1,723 | 29 |
| | 60-69 | 69 | 59 | 3 | <1% | 4,073 | 74 | 945 | 653 | 0 | 0 | 1,672 | 593 | 837 | 29 |
| | 70-79 | 71 | 64 | 5 | <1% | 5,787 | 37 | 1,251 | 1,491 | 0 | 0 | 2,779 | 1,368 | 2,026 | 32 |
| | 80+ | 59 | 53 | 5 | <1% | 5,884 | 40 | 838 | 1,204 | 0 | 0 | 2,082 | 3,204 | 4,337 | 26 |
| Slovenia | 25-49 | 47 | 23 | 1 | <1% | 96 | 4 | 17 | 5 | 0 | 0 | 26 | 14 | 17 | 18 |
| | 50-59 | 63 | 40 | 2 | <1% | 243 | 18 | 78 | 57 | 0 | 0 | 153 | 81 | 132 | 39 |
| | 60+ | 73 | 58 | 11 | 3 | 6,913 | 182 | 1,822 | 2,898 | 128 | 44 | 5,074 | 1,175 | 2,038 | 42 |
| | 60-69 | 72 | 54 | 7 | 1 | 830 | 45 | 278 | 330 | 5 | 0 | 658 | 294 | 528 | 44 |
| | 70-79 | 78 | 64 | 13 | 4 | 1,582 | 57 | 512 | 820 | 29 | 8 | 1,426 | 838 | 1,593 | 47 |

| Countries, areas and territories | Age group (years) | Vaccination uptake | | | | Number of lives | | | | | | | Mortality rate per 100,000 | | |
|---|---|---|---|---|---|---|---|---|---|---|---|---|---|---|---|
| | | $VU_2$ | $VU_3$ | $VU_4$ | $VU_5$ | Reported deaths | Saved after $d_1$ | Saved after $d_2$ | Saved after $d_3$ | Saved after $d_4$ | Saved after $d_5$ | Total averted | Reported | Total Expected | % change |
| | 80+ | 69 | 56 | 16 | 4 | 4,501 | 80 | 1,032 | 1,748 | 94 | 36 | 2,990 | 3,829 | 6,373 | 40 |
| Spain | 25-49 | 75 | 52 | 5 | <1% | 1,160 | 196 | 564 | 296 | 0 | 0 | 1,056 | 7 | 14 | 50 |
| | 50-59 | 79 | 76 | 13 | <1% | 2,771 | 548 | 1,488 | 1,353 | 17 | 0 | 3,406 | 38 | 85 | 55 |
| | 60+ | 92 | 91 | 60 | <1% | 66,327 | 2,073 | 43,711 | 67,669 | 6,274 | 9 | 119,736 | 526 | 1,474 | 64 |
| | 60-69 | 86 | 89 | 47 | <1% | 7,324 | 1,056 | 3,041 | 5,234 | 248 | 0 | 9,579 | 130 | 299 | 57 |
| | 70-79 | 93 | 92 | 67 | <1% | 14,632 | 1,008 | 7,690 | 13,364 | 1,014 | 0 | 23,076 | 358 | 922 | 61 |
| | 80+ | 100 | 93 | 76 | <1% | 44,371 | 9 | 32,980 | 49,071 | 5,012 | 9 | 87,081 | 1,539 | 4,559 | 66 |
| Sweden | 25-49 | 81 | 53 | 12 | <1% | 208 | 0 | 67 | 49 | 1 | 0 | 117 | 6 | 9 | 33 |
| | 50-59 | 89 | 75 | 31 | 1 | 392 | 2 | 108 | 187 | 36 | 0 | 333 | 29 | 54 | 46 |
| | 60+ | 90 | 86 | 73 | 3 | 15,509 | 148 | 2,153 | 6,224 | 8,064 | 173 | 16,762 | 570 | 1,185 | 52 |
| | 60-69 | 91 | 85 | 63 | 3 | 1,123 | 27 | 263 | 593 | 340 | 11 | 1,234 | 100 | 211 | 53 |
| | 70-79 | 93 | 90 | 82 | 4 | 3,783 | 83 | 668 | 1,980 | 2,273 | 78 | 5,082 | 370 | 867 | 57 |
| | 80+ | 83 | 82 | 77 | 2 | 10,603 | 38 | 1,222 | 3,651 | 5,451 | 84 | 10,446 | 1,821 | 3,615 | 50 |
| Switzerland | 25-49 | 69 | 47 | 2 | <1% | 91 | 0 | 32 | 14 | 0 | 0 | 46 | 3 | 5 | 40 |
| | 50-59 | 76 | 67 | 6 | <1% | 265 | 7 | 108 | 69 | 0 | 0 | 184 | 20 | 35 | 43 |

| Countries, areas and territories | Age group (years) | Vaccination uptake VU$_2$ | VU$_3$ | VU$_4$ | VU$_5$ | Number of lives Reported deaths | Saved after $d_1$ | Saved after $d_2$ | Saved after $d_3$ | Saved after $d_4$ | Saved after $d_5$ | Total averted | Mortality rate per 100,000 Reported | Total Expected | % change |
|---|---|---|---|---|---|---|---|---|---|---|---|---|---|---|---|
| | 60+ | 85 | 100 | 24 | 5 | 8,325 | 216 | 2,874 | 4,572 | 281 | 81 | 8,024 | 377 | 741 | 49 |
| | 60-69 | 81 | 90 | 13 | 1 | 707 | 19 | 275 | 255 | 1 | 0 | 550 | 71 | 127 | 44 |
| | 70-79 | 88 | 100 | 29 | 7 | 1,744 | 33 | 582 | 937 | 65 | 10 | 1,627 | 234 | 453 | 48 |
| | 80+ | 90 | 100 | 38 | 10 | 5,874 | 164 | 2,017 | 3,380 | 215 | 71 | 5,847 | 1,245 | 2,483 | 50 |
| Ukraine | 60+ | 38 | 55 | 27 | <1% | 82,115 | 795 | 11,074 | 984 | 817 | 0 | 13,670 | 897 | 1,046 | 14 |
| United Kingdom (England) | 25-49 | 83 | 61 | 6 | <1% | 5,239 | 512 | 2,666 | 2,145 | 250 | 0 | 5,573 | 28 | 58 | 52 |
| | 50-59 | 93 | 82 | 51 | <1% | 9,011 | 528 | 5,573 | 6,180 | 1,006 | 0 | 13,287 | 117 | 289 | 60 |
| | 60+ | 100 | 96 | 62 | 48 | 160,550 | 18,441 | 67,687 | 138,466 | 49,662 | 64,874 | 339,130 | 1,168 | 3,634 | 68 |
| | 60-69 | 100 | 94 | 74 | <1% | 18,739 | 899 | 13,585 | 17,615 | 4,859 | 0 | 36,958 | 309 | 919 | 66 |
| | 70-79 | 99 | 97 | 34 | 86 | 39,577 | 3,297 | 18,225 | 38,342 | 4,949 | 16,370 | 81,183 | 812 | 2,477 | 67 |
| | 80+ | 100 | 99 | 86 | 88 | 102,234 | 14,245 | 35,877 | 82,509 | 39,854 | 48,504 | 220,989 | 3,630 | 11,476 | 68 |
| United Kingdom (Scotland) | 25-49 | 82 | 66 | 14 | 1 | 307 | 34 | 214 | 149 | 0 | 0 | 397 | 17 | 39 | 56 |
| | 50-59 | 95 | 88 | 57 | 3 | 561 | 27 | 577 | 428 | 51 | 0 | 1,083 | 71 | 208 | 66 |
| | 60+ | 100 | 100 | 91 | 33 | 9,659 | 272 | 5,401 | 10,105 | 3,540 | 1,340 | 20,658 | 673 | 2,113 | 68 |

| | | Vaccination uptake | | | | Number of lives | | | | | | | Mortality rate per 100,000 | | |
|---|---|---|---|---|---|---|---|---|---|---|---|---|---|---|---|
| **Countries, areas and territories** | **Age group (years)** | **$VU_2$** | **$VU_3$** | **$VU_4$** | **$VU_5$** | **Reported deaths** | **Saved after $d_1$** | **Saved after $d_2$** | **Saved after $d_3$** | **Saved after $d_4$** | **Saved after $d_5$** | **Total averted** | **Reported** | **Total Expected** | **% change** |
| Kosovo[1] | 25-49 | 59 | 5 | 0 | <1% | 85 | 0 | 8 | 0 | 0 | 0 | 8 | 14 | 15 | 7 |
| | 50-59 | 69 | 10 | 0 | <1% | 182 | 0 | 31 | 0 | 0 | 0 | 31 | 93 | 108 | 14 |
| | 60+ | 71 | 20 | 1 | <1% | 1,432 | 10 | 356 | 27 | 0 | 0 | 393 | 573 | 730 | 22 |
| | 60-69 | 74 | 19 | 1 | <1% | 401 | 0 | 93 | 6 | 0 | 0 | 99 | 287 | 358 | 20 |
| | 70-79 | 75 | 25 | 1 | <1% | 607 | 5 | 190 | 16 | 0 | 0 | 211 | 807 | 1,087 | 26 |
| | 80+ | 50 | 14 | 1 | <1% | 424 | 5 | 73 | 5 | 0 | 0 | 83 | 1,209 | 1,445 | 16 |
| Total | - | 83 | 74 | 29 | 7 | 1,050,501 | 52,006 | 355,807 | 656,331 | 150,837 | 79,054 | 1,294,035 | 243 | 541 | 55 |

[1] All references to Kosovo in this document should be understood to be in the context of the United Nations Security Council resolution 1244 (1999).

*Supplementary table 5: Regional summary of sensitivity analyses by age and variant, for all 34 countries, areas and territories (CAT) included in the analysis between weeks 50/2020 and 12/2023.*

| | | Index | | | Alpha | | | Delta | | | Omicron | | | Total | | |
|---|---|---|---|---|---|---|---|---|---|---|---|---|---|---|---|---|
| **Age group** | **Scenario** | **Total lives saved** | **Mortality rate expected** | **Percentage change** | **Total lives saved** | **Mortality rate expected** | **Percentage change** | **Total lives saved** | **Mortality rate expected** | **Percentage change** | **Total lives saved** | **Mortality rate expected** | **Percentage change** | **Total lives saved** | **Mortality rate expected** | **Percentage change** |
| 25-49 years | 1 | 0 | 0 | 0 | 190 | 11 | 4 | 8,825 | 494 | 60 | 15,655 | 877 | 72 | 24,670 | 1,382 | 56 |
| | 2 | 0 | 0 | 0 | 105 | 6 | 2 | 4,390 | 246 | 43 | 5,181 | 290 | 46 | 9,676 | 542 | 33 |
| | 3 | 0 | 0 | 0 | 115 | 6 | 3 | 6,077 | 340 | 51 | 9,264 | 519 | 60 | 15,456 | 866 | 44 |
| | 4 | 0 | 0 | 0 | 221 | 12 | 5 | 7,764 | 435 | 57 | 10,273 | 576 | 63 | 18,258 | 1,023 | 48 |
| | 5 | 0 | 0 | 0 | 166 | 9 | 4 | 7,104 | 398 | 55 | 10,732 | 601 | 64 | 18,002 | 1,009 | 48 |
| | 6 | 0 | 0 | 0 | 161 | 9 | 4 | 5,973 | 335 | 51 | 7,599 | 426 | 55 | 13,733 | 769 | 41 |
| | 7 | 0 | 0 | 0 | 160 | 9 | 4 | 5,844 | 327 | 50 | 7,271 | 407 | 54 | 13,275 | 744 | 40 |
| | 8 | 0 | 0 | 0 | 164 | 9 | 4 | 6,552 | 367 | 53 | 9,058 | 507 | 60 | 15,774 | 884 | 45 |
| 50-59 years | 1 | 0 | 0 | 0 | 1,000 | 126 | 9 | 25,928 | 3,277 | 73 | 44,333 | 5,603 | 79 | 71,261 | 9,007 | 64 |
| | 2 | 0 | 0 | 0 | 537 | 68 | 5 | 10,451 | 1,321 | 52 | 11,785 | 1,490 | 49 | 22,773 | 2,878 | 36 |
| | 3 | 0 | 0 | 0 | 609 | 77 | 6 | 15,829 | 2,001 | 62 | 23,100 | 2,920 | 66 | 39,538 | 4,997 | 50 |
| | 4 | 0 | 0 | 0 | 1,126 | 142 | 10 | 20,087 | 2,539 | 68 | 25,357 | 3,205 | 68 | 46,570 | 5,886 | 54 |

| | | Index | | | Alpha | | | Delta | | | Omicron | | | Total | | |
|---|---|---|---|---|---|---|---|---|---|---|---|---|---|---|---|---|
| Age group | Scenario | Total lives saved | Mortality rate expected | Percentage change | Total lives saved | Mortality rate expected | Percentage change | Total lives saved | Mortality rate expected | Percentage change | Total lives saved | Mortality rate expected | Percentage change | Total lives saved | Mortality rate expected | Percentage change |
| | 5 | 0 | 0 | 0 | 830 | 105 | 8 | 18,800 | 2,376 | 66 | 26,864 | 3,395 | 69 | 46,494 | 5,876 | 54 |
| | 6 | 0 | 0 | 0 | 780 | 99 | 7 | 14,582 | 1,843 | 60 | 18,598 | 2,351 | 61 | 33,960 | 4,292 | 46 |
| | 7 | 0 | 0 | 0 | 782 | 99 | 7 | 14,672 | 1,854 | 60 | 17,475 | 2,209 | 59 | 32,929 | 4,162 | 45 |
| | 8 | 0 | 0 | 0 | 807 | 102 | 7 | 16,780 | 2,121 | 63 | 22,246 | 2,812 | 65 | 39,833 | 5,034 | 50 |
| 60+ years | 1 | 0 | 0 | 0 | 76,172 | 554 | 27 | 653,418 | 4,752 | 78 | 1,792,564 | 13,038 | 83 | 2,522,154 | 18,344 | 72 |
| | 2 | 0 | 0 | 0 | 32,986 | 240 | 14 | 226,148 | 1,645 | 56 | 455,665 | 3,314 | 55 | 714,799 | 5,199 | 42 |
| | 3 | 0 | 0 | 0 | 39,106 | 284 | 16 | 336,123 | 2,445 | 65 | 893,370 | 6,498 | 71 | 1,268,599 | 9,227 | 56 |
| | 4 | 0 | 0 | 0 | 79,814 | 581 | 27 | 464,228 | 3,376 | 72 | 922,066 | 6,706 | 71 | 1,466,108 | 10,663 | 60 |
| | 5 | 0 | 0 | 0 | 56,712 | 412 | 21 | 425,810 | 3,097 | 70 | 1,007,454 | 7,327 | 73 | 1,489,976 | 10,837 | 60 |
| | 6 | 0 | 0 | 0 | 52,913 | 385 | 20 | 299,125 | 2,176 | 63 | 691,761 | 5,031 | 65 | 1,043,799 | 7,592 | 51 |
| | 7 | 0 | 0 | 0 | 52,191 | 380 | 20 | 301,449 | 2,193 | 63 | 628,686 | 4,573 | 63 | 982,326 | 7,145 | 50 |
| | 8 | 0 | 0 | 0 | 54,795 | 399 | 21 | 360,041 | 2,619 | 67 | 823,592 | 5,990 | 69 | 1,238,428 | 9,007 | 56 |
| Total | 1 | 0 | 0 | 0 | 77,362 | 474 | 26 | 688,171 | 4,215 | 78 | 1,852,552 | 11,348 | 83 | 2,618,085 | 16,037 | 71 |
| | 2 | 0 | 0 | 0 | 33,628 | 206 | 13 | 240,989 | 1,476 | 55 | 472,631 | 2,895 | 55 | 747,248 | 4,577 | 42 |

| | | Index | | | Alpha | | | Delta | | | Omicron | | | Total | | |
|---|---|---|---|---|---|---|---|---|---|---|---|---|---|---|---|---|
| Age group | Scenario | Total lives saved | Mortality rate expected | Percentage change | Total lives saved | Mortality rate expected | Percentage change | Total lives saved | Mortality rate expected | Percentage change | Total lives saved | Mortality rate expected | Percentage change | Total lives saved | Mortality rate expected | Percentage change |
| | 3 | 0 | 0 | 0 | 39,830 | 244 | 15 | 358,029 | 2,193 | 65 | 925,734 | 5,671 | 70 | 1,323,593 | 8,108 | 56 |
| | 4 | 0 | 0 | 0 | 81,161 | 497 | 26 | 492,079 | 3,014 | 72 | 957,696 | 5,866 | 71 | 1,530,936 | 9,378 | 59 |
| | 5 | 0 | 0 | 0 | 57,708 | 353 | 20 | 451,714 | 2,767 | 70 | 1,045,050 | 6,401 | 73 | 1,554,472 | 9,522 | 60 |
| | 6 | 0 | 0 | 0 | 53,854 | 330 | 19 | 319,680 | 1,958 | 62 | 717,958 | 4,398 | 65 | 1,091,492 | 6,686 | 51 |
| | 7 | 0 | 0 | 0 | 53,133 | 325 | 19 | 321,965 | 1,972 | 62 | 653,432 | 4,003 | 63 | 1,028,530 | 6,300 | 49 |
| | 8 | 0 | 0 | 0 | 55,766 | 342 | 20 | 383,373 | 2,348 | 66 | 854,896 | 5,237 | 69 | 1,294,035 | 7,927 | 55 |

Scenario descriptions: 1: High VE values; 2: Low VE values; 3: Long lag time; 4: Short lag time; 5: Long vaccine waning time; 6: Short vaccine waning time; 7: High prior immunity; 8: Low prior immunity

*Supplementary table 6: Results of each sensitivity analyses, showing the total number of deaths averted for each country or area, expected mortality rate per 100,000 population and the % expected deaths averted by vaccination for the population aged 25 years and older, in 34 WHO European Region countries, areas and territories (CAT), for weeks 50/2020 to 12/2023.*

| | | Deaths | | Mortality rate | | |
|---|---|---|---|---|---|---|
| **Countries, areas and territories** | **Sensitivity analysis** | **Reported** | **Averted** | **Reported** | **Expected** | **Percentage change** |
| Austria | 1 | 17,218 | 41,505 | 249 | 849 | 71 |
| | 2 | 17,218 | 13,632 | 249 | 446 | 44 |
| | 3 | 17,218 | 22,089 | 249 | 568 | 56 |
| | 4 | 17,218 | 29,341 | 249 | 673 | 63 |
| | 5 | 17,218 | 27,201 | 249 | 642 | 61 |
| | 6 | 17,218 | 19,607 | 249 | 533 | 53 |
| | 7 | 17,218 | 18,342 | 249 | 514 | 52 |
| | 8 | 17,218 | 22,914 | 249 | 580 | 57 |
| Belgium | 1 | 15,921 | 58,638 | 179 | 837 | 79 |
| | 2 | 15,921 | 14,691 | 179 | 343 | 48 |
| | 3 | 15,921 | 26,167 | 179 | 472 | 62 |
| | 4 | 15,921 | 34,163 | 179 | 562 | 68 |
| | 5 | 15,921 | 32,247 | 179 | 540 | 67 |
| | 6 | 15,921 | 22,438 | 179 | 430 | 58 |
| | 7 | 15,921 | 20,259 | 179 | 406 | 56 |
| | 8 | 15,921 | 26,397 | 179 | 475 | 62 |
| Croatia | 1 | 4,805 | 5,163 | 139 | 288 | 52 |
| | 2 | 4,805 | 2,417 | 139 | 209 | 33 |
| | 3 | 4,805 | 3,586 | 139 | 242 | 43 |
| | 4 | 4,805 | 4,063 | 139 | 256 | 46 |
| | 5 | 4,805 | 4,112 | 139 | 258 | 46 |
| | 6 | 4,805 | 3,119 | 139 | 229 | 39 |

| | | Deaths | | Mortality rate | | |
|---|---|---|---|---|---|---|
| Countries, areas and territories | Sensitivity analysis | Reported | Averted | Reported | Expected | Percentage change |
| | 7 | 4,805 | 3,084 | 139 | 228 | 39 |
| | 8 | 4,805 | 3,616 | 139 | 243 | 43 |
| Cyprus | 1 | 1,490 | 4,964 | 251 | 1,086 | 77 |
| | 2 | 1,490 | 1,425 | 251 | 491 | 49 |
| | 3 | 1,490 | 2,687 | 251 | 703 | 64 |
| | 4 | 1,490 | 3,046 | 251 | 763 | 67 |
| | 5 | 1,490 | 3,150 | 251 | 781 | 68 |
| | 6 | 1,490 | 2,200 | 251 | 621 | 60 |
| | 7 | 1,490 | 2,178 | 251 | 617 | 59 |
| | 8 | 1,490 | 2,658 | 251 | 698 | 64 |
| Czechia | 1 | 33,292 | 38,297 | 400 | 860 | 53 |
| | 2 | 33,292 | 15,827 | 400 | 590 | 32 |
| | 3 | 33,292 | 22,814 | 400 | 674 | 41 |
| | 4 | 33,292 | 31,575 | 400 | 779 | 49 |
| | 5 | 33,292 | 28,326 | 400 | 740 | 46 |
| | 6 | 33,292 | 21,657 | 400 | 660 | 39 |
| | 7 | 33,292 | 20,408 | 400 | 645 | 38 |
| | 8 | 33,292 | 24,663 | 400 | 696 | 43 |
| Denmark | 1 | 7,119 | 36,958 | 156 | 964 | 84 |
| | 2 | 7,119 | 7,948 | 156 | 330 | 53 |
| | 3 | 7,119 | 16,364 | 156 | 514 | 70 |
| | 4 | 7,119 | 17,381 | 156 | 536 | 71 |
| | 5 | 7,119 | 18,377 | 156 | 558 | 72 |
| | 6 | 7,119 | 13,109 | 156 | 443 | 65 |

| | | Deaths | | Mortality rate | | |
|---|---|---|---|---|---|---|
| **Countries, areas and territories** | **Sensitivity analysis** | **Reported** | **Averted** | **Reported** | **Expected** | **Percentage change** |
| | 7 | 7,119 | 11,590 | 156 | 409 | 62 |
| | 8 | 7,119 | 15,200 | 156 | 488 | 68 |
| Estonia | 1 | 2,633 | 3,810 | 184 | 450 | 59 |
| | 2 | 2,633 | 1,571 | 184 | 294 | 37 |
| | 3 | 2,633 | 2,444 | 184 | 355 | 48 |
| | 4 | 2,633 | 2,887 | 184 | 386 | 52 |
| | 5 | 2,633 | 2,871 | 184 | 385 | 52 |
| | 6 | 2,633 | 2,119 | 184 | 332 | 45 |
| | 7 | 2,633 | 2,101 | 184 | 331 | 44 |
| | 8 | 2,633 | 2,486 | 184 | 358 | 49 |
| Finland | 1 | 8,508 | 32,344 | 176 | 844 | 79 |
| | 2 | 8,508 | 10,935 | 176 | 402 | 56 |
| | 3 | 8,508 | 18,739 | 176 | 563 | 69 |
| | 4 | 8,508 | 19,774 | 176 | 585 | 70 |
| | 5 | 8,508 | 20,774 | 176 | 605 | 71 |
| | 6 | 8,508 | 15,380 | 176 | 494 | 64 |
| | 7 | 8,508 | 13,602 | 176 | 457 | 61 |
| | 8 | 8,508 | 17,539 | 176 | 538 | 67 |
| France | 1 | 98,828 | 196,944 | 182 | 544 | 67 |
| | 2 | 98,828 | 64,285 | 182 | 300 | 39 |
| | 3 | 98,828 | 112,586 | 182 | 389 | 53 |
| | 4 | 98,828 | 128,343 | 182 | 418 | 56 |
| | 5 | 98,828 | 129,788 | 182 | 420 | 57 |
| | 6 | 98,828 | 96,457 | 182 | 359 | 49 |

| | | Deaths | | Mortality rate | | |
|---|---|---|---|---|---|---|
| Countries, areas and territories | Sensitivity analysis | Reported | Averted | Reported | Expected | Percentage change |
| | 7 | 98,828 | 91,441 | 182 | 350 | 48 |
| | 8 | 98,828 | 111,667 | 182 | 387 | 53 |
| Germany | 1 | 270,840 | 525,526 | 375 | 1,102 | 66 |
| | 2 | 270,840 | 188,096 | 375 | 635 | 41 |
| | 3 | 270,840 | 311,702 | 375 | 806 | 53 |
| | 4 | 270,840 | 365,162 | 375 | 880 | 57 |
| | 5 | 270,840 | 364,092 | 375 | 879 | 57 |
| | 6 | 270,840 | 261,388 | 375 | 737 | 49 |
| | 7 | 270,840 | 250,158 | 375 | 721 | 48 |
| | 8 | 270,840 | 308,914 | 375 | 802 | 53 |
| Greece | 1 | 33,445 | 72,709 | 362 | 1,150 | 69 |
| | 2 | 33,445 | 26,636 | 362 | 651 | 44 |
| | 3 | 33,445 | 43,825 | 362 | 837 | 57 |
| | 4 | 33,445 | 51,012 | 362 | 915 | 60 |
| | 5 | 33,445 | 50,807 | 362 | 912 | 60 |
| | 6 | 33,445 | 38,031 | 362 | 774 | 53 |
| | 7 | 33,445 | 36,424 | 362 | 757 | 52 |
| | 8 | 33,445 | 43,961 | 362 | 838 | 57 |
| Hungary | 1 | 41,228 | 43,302 | 532 | 1,090 | 51 |
| | 2 | 41,228 | 22,153 | 532 | 817 | 35 |
| | 3 | 41,228 | 30,282 | 532 | 922 | 42 |
| | 4 | 41,228 | 36,171 | 532 | 998 | 47 |
| | 5 | 41,228 | 35,249 | 532 | 986 | 46 |
| | 6 | 41,228 | 27,368 | 532 | 885 | 40 |

| | | Deaths | | Mortality rate | | |
|---|---|---|---|---|---|---|
| **Countries, areas and territories** | **Sensitivity analysis** | **Reported** | **Averted** | **Reported** | **Expected** | **Percentage change** |
| | 7 | 41,228 | 25,862 | 532 | 865 | 38 |
| | 8 | 41,228 | 31,023 | 532 | 932 | 43 |
| Iceland | 1 | 235 | 1,079 | 104 | 583 | 82 |
| | 2 | 235 | 244 | 104 | 212 | 51 |
| | 3 | 235 | 568 | 104 | 356 | 71 |
| | 4 | 235 | 568 | 104 | 356 | 71 |
| | 5 | 235 | 625 | 104 | 381 | 73 |
| | 6 | 235 | 412 | 104 | 287 | 64 |
| | 7 | 235 | 373 | 104 | 270 | 61 |
| | 8 | 235 | 512 | 104 | 331 | 69 |
| Ireland | 1 | 6,002 | 18,309 | 200 | 808 | 75 |
| | 2 | 6,002 | 4,776 | 200 | 358 | 44 |
| | 3 | 6,002 | 9,139 | 200 | 503 | 60 |
| | 4 | 6,002 | 9,704 | 200 | 522 | 62 |
| | 5 | 6,002 | 10,330 | 200 | 543 | 63 |
| | 6 | 6,002 | 7,182 | 200 | 438 | 54 |
| | 7 | 6,002 | 6,714 | 200 | 423 | 53 |
| | 8 | 6,002 | 8,558 | 200 | 484 | 59 |
| Israel | 1 | 18,066 | 99,123 | 1,390 | 9,014 | 85 |
| | 2 | 18,066 | 28,440 | 1,390 | 3,577 | 61 |
| | 3 | 18,066 | 49,076 | 1,390 | 5,164 | 73 |
| | 4 | 18,066 | 64,970 | 1,390 | 6,387 | 78 |
| | 5 | 18,066 | 62,578 | 1,390 | 6,203 | 78 |
| | 6 | 18,066 | 42,241 | 1,390 | 4,639 | 70 |

| | | Deaths | | Mortality rate | | |
|---|---|---|---|---|---|---|
| Countries, areas and territories | Sensitivity analysis | Reported | Averted | Reported | Expected | Percentage change |
| | 7 | 18,066 | 40,714 | 1,390 | 4,521 | 69 |
| | 8 | 18,066 | 51,104 | 1,390 | 5,320 | 74 |
| Italy | 1 | 131,583 | 359,171 | 245 | 913 | 73 |
| | 2 | 131,583 | 90,785 | 245 | 414 | 41 |
| | 3 | 131,583 | 175,512 | 245 | 571 | 57 |
| | 4 | 131,583 | 204,376 | 245 | 625 | 61 |
| | 5 | 131,583 | 208,509 | 245 | 633 | 61 |
| | 6 | 131,583 | 143,423 | 245 | 512 | 52 |
| | 7 | 131,583 | 134,870 | 245 | 496 | 51 |
| | 8 | 131,583 | 171,876 | 245 | 565 | 57 |
| Latvia | 1 | 5,659 | 5,028 | 359 | 678 | 47 |
| | 2 | 5,659 | 2,356 | 359 | 508 | 29 |
| | 3 | 5,659 | 3,482 | 359 | 580 | 38 |
| | 4 | 5,659 | 4,193 | 359 | 625 | 43 |
| | 5 | 5,659 | 3,982 | 359 | 611 | 41 |
| | 6 | 5,659 | 3,299 | 359 | 568 | 37 |
| | 7 | 5,659 | 3,147 | 359 | 558 | 36 |
| | 8 | 5,659 | 3,621 | 359 | 588 | 39 |
| Lithuania | 1 | 8,195 | 8,447 | 357 | 726 | 51 |
| | 2 | 8,195 | 4,292 | 357 | 545 | 34 |
| | 3 | 8,195 | 6,036 | 357 | 621 | 43 |
| | 4 | 8,195 | 7,169 | 357 | 670 | 47 |
| | 5 | 8,195 | 6,938 | 357 | 660 | 46 |
| | 6 | 8,195 | 5,564 | 357 | 600 | 40 |

| | | Deaths | | Mortality rate | | |
|---|---|---|---|---|---|---|
| **Countries, areas and territories** | **Sensitivity analysis** | **Reported** | **Averted** | **Reported** | **Expected** | **Percentage change** |
| | 7 | 8,195 | 5,587 | 357 | 601 | 41 |
| | 8 | 8,195 | 6,289 | 357 | 632 | 44 |
| Luxembourg | 1 | 754 | 2,253 | 196 | 780 | 75 |
| | 2 | 754 | 547 | 196 | 337 | 42 |
| | 3 | 754 | 1,097 | 196 | 480 | 59 |
| | 4 | 754 | 1,290 | 196 | 530 | 63 |
| | 5 | 754 | 1,299 | 196 | 533 | 63 |
| | 6 | 754 | 868 | 196 | 421 | 53 |
| | 7 | 754 | 794 | 196 | 402 | 51 |
| | 8 | 754 | 1,064 | 196 | 472 | 58 |
| Malta | 1 | 504 | 2,589 | 391 | 2,399 | 84 |
| | 2 | 504 | 595 | 391 | 852 | 54 |
| | 3 | 504 | 1,233 | 391 | 1,347 | 71 |
| | 4 | 504 | 1,301 | 391 | 1,400 | 72 |
| | 5 | 504 | 1,436 | 391 | 1,505 | 74 |
| | 6 | 504 | 904 | 391 | 1,092 | 64 |
| | 7 | 504 | 878 | 391 | 1,072 | 64 |
| | 8 | 504 | 1,147 | 391 | 1,281 | 69 |
| North Macedonia | 1 | 7,592 | 4,012 | 1,725 | 2,637 | 35 |
| | 2 | 7,592 | 2,428 | 1,725 | 2,277 | 24 |
| | 3 | 7,592 | 3,108 | 1,725 | 2,431 | 29 |
| | 4 | 7,592 | 3,596 | 1,725 | 2,542 | 32 |
| | 5 | 7,592 | 3,486 | 1,725 | 2,517 | 31 |
| | 6 | 7,592 | 2,944 | 1,725 | 2,394 | 28 |

| | | Deaths | | Mortality rate | | |
|---|---|---|---|---|---|---|
| **Countries, areas and territories** | **Sensitivity analysis** | **Reported** | **Averted** | **Reported** | **Expected** | **Percentage change** |
| | 7 | 7,592 | 2,910 | 1,725 | 2,386 | 28 |
| | 8 | 7,592 | 3,221 | 1,725 | 2,457 | 30 |
| Netherlands | 1 | 12,990 | 39,614 | 94 | 381 | 75 |
| | 2 | 12,990 | 9,264 | 94 | 161 | 42 |
| | 3 | 12,990 | 18,122 | 94 | 226 | 58 |
| | 4 | 12,990 | 20,958 | 94 | 246 | 62 |
| | 5 | 12,990 | 22,246 | 94 | 256 | 63 |
| | 6 | 12,990 | 13,212 | 94 | 190 | 51 |
| | 7 | 12,990 | 13,529 | 94 | 192 | 51 |
| | 8 | 12,990 | 17,255 | 94 | 219 | 57 |
| Portugal | 1 | 21,347 | 60,323 | 336 | 1,285 | 74 |
| | 2 | 21,347 | 14,082 | 336 | 558 | 40 |
| | 3 | 21,347 | 28,165 | 336 | 779 | 57 |
| | 4 | 21,347 | 31,819 | 336 | 837 | 60 |
| | 5 | 21,347 | 32,867 | 336 | 853 | 61 |
| | 6 | 21,347 | 22,834 | 336 | 695 | 52 |
| | 7 | 21,347 | 20,880 | 336 | 665 | 49 |
| | 8 | 21,347 | 27,044 | 336 | 762 | 56 |
| Republic of Moldova | 1 | 11,966 | 4,026 | 1,688 | 2,256 | 25 |
| | 2 | 11,966 | 2,552 | 1,688 | 2,048 | 18 |
| | 3 | 11,966 | 3,280 | 1,688 | 2,151 | 22 |
| | 4 | 11,966 | 3,620 | 1,688 | 2,199 | 23 |
| | 5 | 11,966 | 3,552 | 1,688 | 2,189 | 23 |
| | 6 | 11,966 | 3,090 | 1,688 | 2,124 | 21 |

| | | Deaths | | Mortality rate | | |
|---|---|---|---|---|---|---|
| Countries, areas and territories | Sensitivity analysis | Reported | Averted | Reported | Expected | Percentage change |
| | 7 | 11,966 | 3,056 | 1,688 | 2,119 | 20 |
| | 8 | 11,966 | 3,342 | 1,688 | 2,160 | 22 |
| Romania | 1 | 51,144 | 14,883 | 519 | 670 | 23 |
| | 2 | 51,144 | 9,809 | 519 | 618 | 16 |
| | 3 | 51,144 | 12,430 | 519 | 645 | 20 |
| | 4 | 51,144 | 13,601 | 519 | 657 | 21 |
| | 5 | 51,144 | 13,427 | 519 | 655 | 21 |
| | 6 | 51,144 | 11,499 | 519 | 636 | 18 |
| | 7 | 51,144 | 11,727 | 519 | 638 | 19 |
| | 8 | 51,144 | 12,628 | 519 | 647 | 20 |
| Slovakia | 1 | 17,768 | 9,092 | 462 | 699 | 34 |
| | 2 | 17,768 | 5,377 | 462 | 602 | 23 |
| | 3 | 17,768 | 6,603 | 462 | 634 | 27 |
| | 4 | 17,768 | 8,711 | 462 | 689 | 33 |
| | 5 | 17,768 | 7,772 | 462 | 664 | 30 |
| | 6 | 17,768 | 6,641 | 462 | 635 | 27 |
| | 7 | 17,768 | 6,364 | 462 | 628 | 26 |
| | 8 | 17,768 | 7,174 | 462 | 649 | 29 |
| Slovenia | 1 | 7,252 | 7,411 | 417 | 844 | 51 |
| | 2 | 7,252 | 3,510 | 417 | 619 | 33 |
| | 3 | 7,252 | 5,209 | 417 | 717 | 42 |
| | 4 | 7,252 | 5,945 | 417 | 760 | 45 |
| | 5 | 7,252 | 5,946 | 417 | 760 | 45 |
| | 6 | 7,252 | 4,551 | 417 | 679 | 39 |

| | | Deaths | | Mortality rate | | |
|---|---|---|---|---|---|---|
| Countries, areas and territories | Sensitivity analysis | Reported | Averted | Reported | Expected | Percentage change |
| | 7 | 7,252 | 4,502 | 417 | 677 | 38 |
| | 8 | 7,252 | 5,253 | 417 | 720 | 42 |
| Spain | 1 | 70,258 | 313,314 | 189 | 1,031 | 82 |
| | 2 | 70,258 | 59,381 | 189 | 349 | 46 |
| | 3 | 70,258 | 130,653 | 189 | 540 | 65 |
| | 4 | 70,258 | 141,751 | 189 | 570 | 67 |
| | 5 | 70,258 | 153,809 | 189 | 602 | 69 |
| | 6 | 70,258 | 98,189 | 189 | 453 | 58 |
| | 7 | 70,258 | 98,366 | 189 | 453 | 58 |
| | 8 | 70,258 | 124,198 | 189 | 523 | 64 |
| Sweden | 1 | 16,109 | 28,194 | 200 | 549 | 64 |
| | 2 | 16,109 | 11,534 | 200 | 343 | 42 |
| | 3 | 16,109 | 18,077 | 200 | 424 | 53 |
| | 4 | 16,109 | 19,178 | 200 | 437 | 54 |
| | 5 | 16,109 | 20,314 | 200 | 451 | 56 |
| | 6 | 16,109 | 14,512 | 200 | 380 | 47 |
| | 7 | 16,109 | 13,874 | 200 | 372 | 46 |
| | 8 | 16,109 | 17,212 | 200 | 413 | 52 |
| Switzerland | 1 | 8,681 | 14,327 | 393 | 1,042 | 62 |
| | 2 | 8,681 | 4,940 | 393 | 617 | 36 |
| | 3 | 8,681 | 7,925 | 393 | 752 | 48 |
| | 4 | 8,681 | 10,239 | 393 | 857 | 54 |
| | 5 | 8,681 | 9,649 | 393 | 830 | 53 |
| | 6 | 8,681 | 7,116 | 393 | 716 | 45 |

| | | Deaths | | Mortality rate | | |
|---|---|---|---|---|---|---|
| Countries, areas and territories | Sensitivity analysis | Reported | Averted | Reported | Expected | Percentage change |
| | 7 | 8,681 | 6,731 | 393 | 698 | 44 |
| | 8 | 8,681 | 8,254 | 393 | 767 | 49 |
| United Kingdom (England) | 1 | 174,800 | 811,388 | 1,271 | 7,173 | 82 |
| | 2 | 174,800 | 208,058 | 1,271 | 2,785 | 54 |
| | 3 | 174,800 | 375,196 | 1,271 | 4,000 | 68 |
| | 4 | 174,800 | 428,954 | 1,271 | 4,391 | 71 |
| | 5 | 174,800 | 441,085 | 1,271 | 4,479 | 72 |
| | 6 | 174,800 | 300,958 | 1,271 | 3,460 | 63 |
| | 7 | 174,800 | 274,297 | 1,271 | 3,266 | 61 |
| | 8 | 174,800 | 357,990 | 1,271 | 3,875 | 67 |
| United Kingdom (Scotland) | 1 | 20,186 | 100,946 | 1,407 | 8,444 | 83 |
| | 2 | 20,186 | 23,556 | 1,407 | 3,049 | 54 |
| | 3 | 20,186 | 45,473 | 1,407 | 4,577 | 69 |
| | 4 | 20,186 | 49,938 | 1,407 | 4,888 | 71 |
| | 5 | 20,186 | 52,391 | 1,407 | 5,059 | 72 |
| | 6 | 20,186 | 35,443 | 1,407 | 3,878 | 64 |
| | 7 | 20,186 | 33,281 | 1,407 | 3,727 | 62 |
| | 8 | 20,186 | 42,796 | 1,407 | 4,391 | 68 |
| Ukraine | 1 | 164,230 | 32,016 | 1,794 | 2,144 | 16 |
| | 2 | 164,230 | 22,694 | 1,794 | 2,042 | 12 |
| | 3 | 164,230 | 25,788 | 1,794 | 2,076 | 14 |
| | 4 | 164,230 | 31,474 | 1,794 | 2,138 | 16 |
| | 5 | 164,230 | 28,698 | 1,794 | 2,108 | 15 |
| | 6 | 164,230 | 26,476 | 1,794 | 2,084 | 14 |

| | | Deaths | | Mortality rate | | |
|---|---|---|---|---|---|---|
| Countries, areas and territories | Sensitivity analysis | Reported | Averted | Reported | Expected | Percentage change |
| | 7 | 164,230 | 25,034 | 1,794 | 2,068 | 13 |
| | 8 | 164,230 | 27,340 | 1,794 | 2,093 | 14 |
| Kosovo[1] | 1 | 1,699 | 553 | 346 | 458 | 24 |
| | 2 | 1,699 | 307 | 346 | 408 | 15 |
| | 3 | 1,699 | 397 | 346 | 426 | 19 |
| | 4 | 1,699 | 518 | 346 | 451 | 23 |
| | 5 | 1,699 | 473 | 346 | 442 | 22 |
| | 6 | 1,699 | 400 | 346 | 427 | 19 |
| | 7 | 1,699 | 394 | 346 | 426 | 19 |
| | 8 | 1,699 | 432 | 346 | 434 | 20 |
| Total | 1 | 1,292,347 | 2,996,258 | 365 | 1,210 | 70 |
| | 2 | 1,292,347 | 879,143 | 365 | 613 | 40 |
| | 3 | 1,292,347 | 1,539,854 | 365 | 799 | 54 |
| | 4 | 1,292,347 | 1,786,791 | 365 | 869 | 58 |
| | 5 | 1,292,347 | 1,808,406 | 365 | 875 | 58 |
| | 6 | 1,292,347 | 1,274,631 | 365 | 724 | 50 |
| | 7 | 1,292,347 | 1,203,471 | 365 | 704 | 48 |
| | 8 | 1,292,347 | 1,509,348 | 365 | 791 | 54 |

[1] All references to Kosovo in this document should be understood to be in the context of the United Nations Security Council resolution 1244 (1999).

Scenario descriptions: 1: High VE values; 2: Low VE values; 3: Long lag time; 4: Short lag time; 5: Long vaccine waning time; 6: Short vaccine waning time; 7: High prior immunity; 8: Low prior immunity

*Supplementary table 7: initial Vaccine Effectiveness (VE) against mortality values identified in the literature according to vaccine dose and dominant variant.*

| Authors | Year | Country | Sudy population | Comparison population | Vaccines used | Variant | VE values |
|---|---|---|---|---|---|---|---|
| Amodio *et al* | 2022 | Italy (Sicily) | Adults | Unvaccinated | Pfizer–BioNTech; AstraZeneca; Moderna; Janssen | Alpha, Delta | VE2 (Alpha): 93% |
| Andersson *et al* | pre-print | Denmark, Finland, Norway and Sweden | 50+ with 2 boosters | 50+ with 1 booster | Pfizer -BioNTech (Comirnaty BA.1 and BA4.5) | Omicron | VE4 (Omicron): 78%<br>VE4 (Omicron): 80% |
| Andrews *et al* | 2022 | England | 16 years and older (prior vaccination not compulsory) | Symptomatic but test negative on PCR | Pfizer–BioNTech; AstraZeneca; Moderna | Alpha, Delta | VE2 (Delta): 95%<br>VE2 (Delta): 94%<br>VE2 (Delta): 99%<br>VE2 (Delta): 97% |
| Arbel *et al* | 2022 | Israel | 60+ with 2 boosters | 60+ with 1 booster | Pfizer–BioNTech | Omicron | VE4 (Omicron): 78% |
| Berec *et al* | 2022 | Czechia | 12+ years | Unvaccinated | Pfizer–BioNTech; AstraZeneca; Moderna; Janssen | Alpha, Delta | VE2 (Alpha): 92%<br>VE2 (Alpha): 96%<br>VE2 (Alpha): 93%<br>VE2 (Alpha): 68%<br>VE3 (Delta): 97% |
| Glatman-Freedman *et al* | 2021 | Israel | 16+ population & no prior positive PCR | Unvaccinated | Pfizer–BioNTech | Alpha | VE1 (Alpha): 72%<br>VE2 (Alpha): 92% |
| Goldberg *et al* | 2022 | Israel | 16 years and older (prior vaccination not compulsory) | Unvaccinated | Pfizer–BioNTech | Alpha | VE1 (Alpha): 70%<br>VE2 (Alpha): 95% |
| Hulme *et al* | pre-print | UK | 18+ with complete series | Unvaccinated | Pfizer–BioNTech; AstraZeneca | Delta | VE3 (Delta): 89% |

| | | | | | | | |
|---|---|---|---|---|---|---|---|
| Kaura *et al* | 2022 | UK (London) | 16+ years, no previous infection, no Moderna vaccine | Unvaccinated | Pfizer–BioNTech; AstraZeneca | Alpha | VE1 (Alpha): 86%<br>VE1 (Alpha): 86% |
| Kislaya *et al* | 2022 | Portugal | 60+ years | Unvaccinated | Pfizer–BioNTech; AstraZeneca; Moderna; Janssen | Omicron | VE3 (Omicron): 59%<br>VE4 (Omicron): 65% |
| Liu *et al* | pre-print | Australia | 65+ & one dose | Unvaccinated | Pfizer–BioNTech; AstraZeneca; Moderna | Omicron | VE2 (Omicron): 73%<br>VE3 (Omicron): 93%<br>VE3 (Omicron): 75%<br>VE4 (Omicron): 93%<br>VE4 (Omicron): 84% |
| Lytras *et al* | 2022 | Greece | General population | Unvaccinated | Pfizer–BioNTech; AstraZeneca; Moderna; Janssen | Alpha, Delta, Omicron | VE1 (Alpha): 65%<br>VE2 (Alpha): 90%<br>VE2 (Delta): 84%<br>VE3 (Delta): 98% |
| Machado *et al* | 2022 | Portugal | 65+ | Unvaccinated | Pfizer–BioNTech; AstraZeneca; Moderna; Janssen | Alpha | VE2 (Alpha): 81%<br>VE2 (Alpha): 95%<br>VE2 (Alpha): 95% |
| Magen *et al* | 2022 | Israel | 60+ years and four doses | 60+ years and three doses | Pfizer–BioNTech | Omicron | VE4 (Omicron): 74% |
| Monge *et al* | 2022 | Spain | 50-59 & complete series | | Pfizer–BioNTech; AstraZeneca; Moderna; Janssen | Alpha | VE2 (Alpha): 89%<br>VE2 (Alpha): 94%<br>VE2 (Alpha): 97% |

| | | | | | | | |
|---|---|---|---|---|---|---|---|
| Park *et al* | 2023 | South Korea | 60+ & vaccination with Pfizer–BioNTech; AstraZeneca; Moderna | Unvaccinated | Pfizer–BioNTech; AstraZeneca; Moderna | Omicron | VE2 (Omicron): 65%<br>VE2 (Omicron): 70%<br>VE3 (Omicron): 95%<br>VE3 (Omicron): 94%<br>VE3 (Omicron): 89%<br>VE4 (Omicron): 96%<br>VE4 (Omicron): 95%<br>VE4 (Omicron): 91% |
| Poukka *et al* | pre-print | Finland | 65+ with complete series and 1+ booster | 65+ with complete series and no booster | Pfizer -BioNTech (Comirnaty BA.1 and BA4.5) | Omicron | VE3 (Omicron): 66% |
| Russo *et al* | 2022 | Italy | Adults 19+, not nursing home residents | Unvaccinated | Unreported | Omicron | VE3 (Omicron): 67% |
| Sheikh *et al* | 2021 | Scotland | Adults with at least one dose | Adults and unvaccinated | Pfizer–BioNTech; AstraZeneca | Delta | VE2 (Delta): 91% |
| Voko *et al* | 2022 | Hungary | Adults aged 18 to 100 | Unvaccinated | Pfizer–BioNTech; Sinopharm; Sputnik-V; AstraZeneca; Moderna; Janssen | Delta | VE1 (Delta) <65: 92% + 92% + 100% + 96% + 87% + 57%<br>VE2 (Delta) <65: 88% + 85% + 100%+ 100%<br>VE2 (Delta) <65: 100% + 73% + 86%<br>VE1 (Delta) >65: 86% + 98% + 100% + 98% + 84% + 55%<br>VE2 (Delta) >65: 92% + 95% + 92% + 97%<br>VE2 (Delta) >65: 83% + 100% + 26% + 82% + 94% |

| | | | | | | | |
|---|---|---|---|---|---|---|---|
| Youg-Xu *et al* | 2022 | USA | Veterans (largely 65+ years) | Unvaccinated | Pfizer–BioNTech; Moderna | Delta, Omicron | VE2 (Delta): 92%<br>VE2 (Omicron): 77%<br>VE3 (Delta): 96%<br>VE3 (Omicron): 94% |
